# Environmental, Social, and Health Burdens in relation to Sleep-Disordered Breathing among Patients of Community-Based Health Centers in the United States

**DOI:** 10.1101/2025.10.01.25337104

**Authors:** Wensu Zhou, Symielle A. Gaston, Rupsha Singh, Kathryn S. Konrad, Ian D. Buller, W. Braxton Jackson, Dayna T. Neo, Robert Voss, Anna Steeves-Reece, Paul D. Juarez, Chandra L. Jackson

## Abstract

**Importance:** Sleep-disordered breathing (SDB) is preventable but underdiagnosed, with disparities among sociodemographic groups with limited material and social resources, partly driven by community-level environmental and social conditions along with healthcare-related factors.

**Objectives:** We sought to investigate associations between multifactorial community-level environmental, social, and health burdens and SDB prevalence. We also determined effect modification by age, sex, race, and ethnicity.

**Design, Setting, and Participants:** Cross-sectional analysis of electronic health records (EHR) data collected in 2022 from 1,957,775 adult patients in the OCHIN network (>2,000 community health centers across 40 U.S. states).

**Exposures:** Patients’ 2022 residential addresses were linked to a census tract-level environmental, social, and health burden (ESHB) index created by the Centers for Disease Control and Prevention and the Agency for Toxic Substances Disease Registry. The ESHB comprises 36 indicators that capture and rank community-level social (e.g., socioeconomic status), environmental (e.g., air pollution), and health (e.g., hypertension) vulnerability. Higher percentile ranks of ESHB (range: 0-1) indicated higher vulnerability.

**Main Outcomes and Measures:** SDB was identified via diagnostic and procedural codes, and subtypes were categorized as obstructive (OSA), central (CSA), other/unspecified (OUSA), multiple apneas, and procedure-based cases. Log-binomial regression estimated prevalence ratios (PRs) and 95% confidence intervals (CIs), adjusting for age, sex, race, and ethnicity. We assessed effect modification by testing cross-product terms.

**Results:** Among 1,957,775 patients (median age was 43.0 years [IQR: 30.0–58.0]; 40.6% men), SDB prevalence was 5.5%, with CSA at 0.03%, OSA at 3.8%, OUSA at 1.4%, multiple sleep apneas at 0.03%, and procedure-based cases at 0.21%. Each 0.1-unit increase in ESHB percentile rank was associated with higher prevalence of SDB (PR=1.01 [1.01–1.01]), OUSA (PR=1.01 [1.01–1.02]), and procedure-based cases (PR=1.05 [1.03–1.06]). The ESHB-SDB association was elevated among adults aged 18-49 years, women, American Indian/Alaska Native and White, and non-Hispanic. ESHB was not associated with CSA.

**Conclusions and Relevance:** Higher community-level environmental, social, and health vulnerabilities were associated with higher SDB prevalence (although future prospective studies are warranted). Our findings underscore the need to address community-level factors with potential tailoring of interventions across sociodemographic groups.

**KEY POINTS:** *Question:* Are community-level environmental, social, and health burdens associated with the prevalence of sleep-disordered breathing (SDB) among medically underserved patients in United States (U.S.) community health centers? If so, to what extent?

*Findings:* This cross-sectional study included electronic health records data from 1,957,775 patients of 2,000 community-based health centers in 40 states linked to the 2022 census tract-level environmental, social, and health burden (ESHB) index. Higher ESHB was associated with a higher prevalence of SDB and its subtypes (e.g., obstructive sleep apnea [OSA] and central sleep apnea [CSA]). Elevated associations were observed among adults aged 18-49 years, women, American Indian/Alaska Native and White, and non-Hispanic.

*Meaning:* Multifactorial community adversity was associated with SDB prevalence. Our findings underscore the need for future intervention studies to address environmental, social, and health burdens at the community level to reduce SDB prevalence in medically underserved populations. Prospective studies are needed.

## INTRODUCTION

Sleep-disordered breathing (SDB) is a prevalent chronic condition characterized by episodes of apnea or hypopnea, classified into obstructive, central, or complex types. ^1,2^ It is linked to other chronic conditions such as cardiovascular disease. ^3,4^ SDB affects an estimated 34% of men and 17% of women aged 30 to 70 years in the United States (U.S.), ^1^ yet many cases remain undiagnosed. Although approximately 12% of U.S. adults have obstructive sleep apnea (OSA), nearly 80% of cases are undiagnosed. ^5^ Identifying contributing factors may help lead to the prevention of SDB.

SDB is also a multifactorial condition that varies by individual-level characteristics (e.g., age, sex, race, ethnicity, and socioeconomic status [SES]) as well as community-level physical/environmental (e.g., air pollution), social (including socioeconomic), and health-related burdens (e.g., hypertension). ^6-8^ For example, among U.S. adults, estimated SDB prevalence was higher among older vs. younger (50-70 vs. 30-49 years), and in men than in women. ^1^ Multiple U.S. adults’ studies have shown higher burdens of SDB and its subtypes (e.g., OSA) among Black, Hispanic, and American Indian/Alaska Native (AI/AN) adults compared with White adults. ^6,9,10^ By SES, undiagnosed OSA was more common among middle-aged and older U.S. adults with lower versus higher education (e.g., high school or less) and income (e.g., under $19,000/year). ^11^ At the community level, multiple factors can contribute to SDB. Poor air quality can worsen SDB by promoting inflammation and reducing airway patency. ^12,13^ Limited access to healthy food contributes to higher levels of overweight/obesity, a key risk factor for SDB. ^7^ Unsafe neighborhoods and limited health-supportive infrastructure can reduce physical activity; inactivity promotes fluid buildup that shifts during sleep, narrows the airway, and increases SDB risk. ^14-18^ Neighborhood SES factors, like residential segregation, poverty, and poor housing, are also associated with increased stress, sleep fragmentation, and greater upper airway collapsibility, a key mechanism in OSA. ^14,19-21^ Limited health infrastructure can also contribute to the prevalence of chronic conditions like asthma, which often co-occurs with and can exacerbate SDB. ^22,23^ Moreover, these neighborhood and community characteristics influencing SDB burdens vary by sociodemographic group; for example, predominantly low-income neighborhoods often have higher air pollution and limited access to health-supportive resources.^24^

Neighborhood/community conditions and SDB relationships may also vary across sociodemographic groups. In fact, prior studies suggest linkages between both neighborhood/community environmental and social adversity with SDB that were stronger among younger vs. older adults, possibly due to more exposure to outdoor environments among younger individuals. ^25^ Stronger magnitudes of association between air pollution and OSA have been observed in women, likely related to physiological differences in airway obstruction combined with sex disparities in medical evaluation and diagnosis, as women often present with atypical symptoms that are frequently dismissed. ^7,26,27^ Stronger relationships between poor neighborhood walkability and OSA severity were observed among Black and Hispanic individuals compared to White individuals, ^28^ possibly reflecting compounded exposures such as unsafe environments and high air pollution. ^29,30^ In sum, prior findings may be explained by differences in exposure to numerous community-level burdens and resources that vary across sociodemographic groups. Thus, it is important to investigate effect modification by age, sex, race, and ethnicity in relationships between a broad set of community-level factors and SDB.

However, prior studies generally investigated community-level physical, social, and health-related burdens individually or analyzed multiple factors separately. ^14,27,31,32^ To address this literature gap, we leveraged the Environmental, Social, and Health Burden (ESHB) index, a composite of 36 indicators capturing environmental, social, and health-related challenges, including environmental quality and conditions; social capital and resources; and preexisting chronic disease burden across census tracts in the U.S. ^33,34^ The ESHB has been previously associated with several SDB risk factors such as asthma, obesity, poor mental health, and potential downstream outcomes including cardiovascular disease. ^35,36^ However, the direct contribution of ESHB to SDB has not been studied, particularly among populations who may be more vulnerable to these burdens. Therefore, we sought to use the ESHB to investigate associations between multifaceted community-level exposures and SDB across various sociodemographic groups. We hypothesized that: 1) individuals in census tracts with a higher (vs. lower) ESHB have a higher prevalence of SDB and its subtypes, and 2) associations are stronger among younger vs. older adults; women vs. men; Asian, AI/AN, Black/African American (BAA), Native Hawaiian/Pacific Islander (NHPI), and Multiracial individuals vs. White adults; and Hispanic vs. Non-Hispanic adults. As a sub aim, we also investigated associations between the three individual subdimensions encompassing the ESHB with SDB (Figure 1).

**Figure 1.**
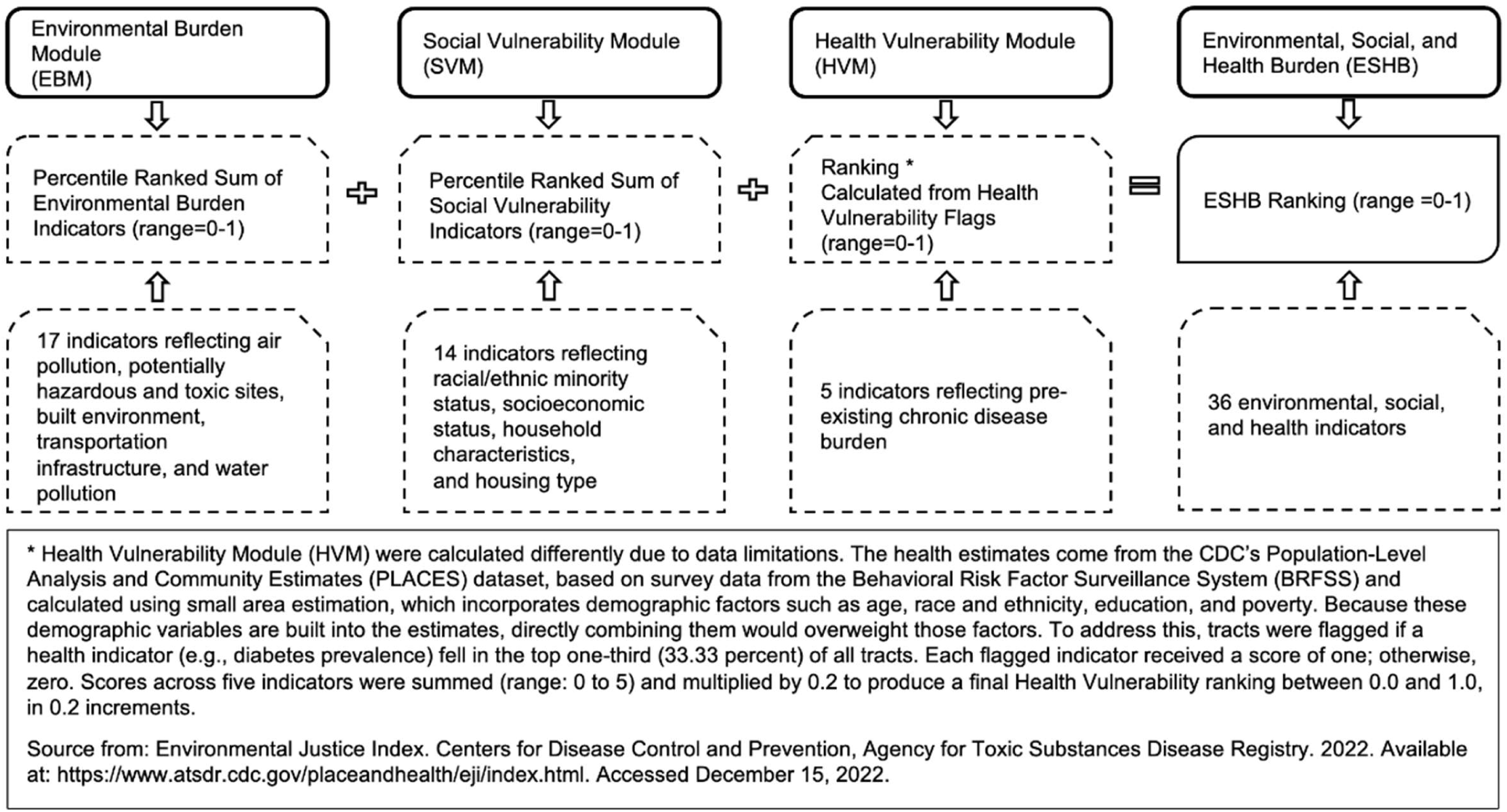
Definition of Environmental Burden Module (EBM), Social Vulnerability Module (SVM), Health Vulnerability Module (HVM), and Environmental, Social, and Health Burden (ESHB)

## METHODS

### Data source and study population

Electronic health record (EHR) data were collected from patients in the ‘Accelerating Data Value Across a National Community Health Center Network’ (ADVANCE), led by OCHIN (detailed in supplemental methods text). ^37,38^ Data from this sample of publicly insured or underinsured patients who are historically underrepresented in research offer a meaningful representation of populations facing health disparities due to their low income and racial and ethnic diversity. ^37,39,40^

This cross-sectional study used 2022 EHR data. Inclusion criteria for this study were (a) adults (aged ≥18 years) who (b) had at least one telehealth or ambulatory encounter between January 1, 2022, and December 31, 2022, (c) had a geolocatable residential address, and (d) had available data on the exposure, outcome, and covariates. The sample selection process is shown in eFigure 1. Compared to included patients (eTable 1), those excluded primarily due to missing ESHB data were more likely to be age ≥50 years, men, White, and non-Hispanic individuals, with similar body mass index (BMI) distribution, lower diabetes prevalence, but higher prevalence of hypertension.

### Exposure assessment: Environmental, Social, and Health Burden (ESHB)

Developed by the Centers for Disease Control and Prevention (CDC) and the Agency for Toxic Substances and Disease Registry (ATSDR), the ESHB index measures cumulative community-level burden at the census tract level by combining 36 environmental, social, and health factors. It consists of three submodules: the Environmental Burden Module [EBM], the Social Vulnerability Module [SVM], and the Health Vulnerability Module [HVM] (further detailed in supplemental text and Figure 1), ^34^ and was linked to patients’ geocoded residential addresses during their first clinical visit in 2022. Higher values (0-1) of ESHB and its three modules indicate higher burdens/vulnerability.

### Outcome assessment: Sleep-disordered breathing (SDB)

Diagnoses of SDB during a 2022 visit (occurring between January 1, 2022, and December 31, 2022) were obtained from patients’ EHR. SDB was identified using ICD-9-CM/ICD-10-CM and/or CPT/HCPCS codes. SDB was further categorized into five subtypes: Obstructive Sleep Apnea (OSA), Central Sleep Apnea (CSA), other/unspecified sleep apnea (OUSA), multiple sleep apneas (patients with more than one subtype diagnosis), and procedure-based cases (eFigure 2 and eTable 2-4).

### Potential Confounders

Sociodemographic characteristics of patients, including age groups, sex, race, and ethnicity, were considered potential confounders according to a Directed Acyclic Graph (DAG; eFigure 3). Categories and other variables considered ^41-44^ but not identified as confounders are further described in Tables 1 and 2.

**Table 1.** Characteristics of study patients, overall and by sleep-disordered breathing (SDB) status (n=1,957,775)

|  | Any SDB | CSA | OSA | OUSA | Multiple sleep apneas | Procedure-based cases | No SDB | Overall |
| --- | --- | --- | --- | --- | --- | --- | --- | --- |
| **N** | 106,857 (5.5) | 562 (0.03) | 73,919 (3.78) | 27,631 (1.41) | 642 (0.03) | 4,103 (0.21) | 1,850,918 (94.5) | 1,957,775 (100.0) |
| **Age category** |  |  |  |  |  |  |  |  |
| 18-34 years | 10,991 (10.3) <sup>a</sup> | 57 (10.1) <sup>a</sup> | 6,529 (8.8) <sup>a</sup> | 3,847 (13.9) <sup>a</sup> | 30 (4.7) <sup>a</sup> | 528 (12.9) <sup>a</sup> | 646,362 (34.9) | 657,353 (33.6) |
| 35-49 years | 26,870 (25.1) | 112 (19.9) | 18,102 (24.5) | 7,740 (28.0) | 118 (18.4) | 798 (19.4) | 507,742 (27.4) | 534,612 (27.3) |
| ≥50 years | 68,996 (64.6) | 393 (69.9) | 49,288 (66.7) | 16,044 (58.1) | 494 (76.9) | 2,777 (67.7) | 696,814 (37.6) | 765,810 (39.1) |
| **Sex** |  |  |  |  |  |  |  |  |
| Women | 53,739 (50.3) <sup>a</sup> | 248 (44.1) <sup>a</sup> | 36,905 (49.9) <sup>a</sup> | 13,639 (49.4) <sup>a</sup> | 266 (41.4) <sup>a</sup> | 2,681 (65.3) <sup>a</sup> | 1,108,118 (59.9) | 1,161,857 (59.3) |
| Men | 53,058 (49.7) | 314 (55.9) | 36,975 (50.0) | 13,974 (50.6) | 375 (58.4) | 1,420 (34.6) | 741,202 (40.0) | 794,260 (40.6) |
| Unknown/Missing | 60 (0.1) | <11 | 39 (0.1) | 18 (0.1) | <11 | <11 | 1,598 (0.1) | 1,658 (0.1) |
| **Race** |  |  |  |  |  |  |  |  |
| AI/AN | 1,509 (1.4) <sup>a</sup> | <11 <sup>a</sup> | 1,071 (1.4) <sup>a</sup> | 356 (1.3) <sup>a</sup> | <11 <sup>a</sup> | 65 (1.6) <sup>a</sup> | 22,017 (1.2) | 23,526 (1.2) |
| Asian | 3,547 (3.3) | <11 | 2,367 (3.2) | 899 (3.3) | 13 (2.0) | 259 (6.3) | 118,126 (6.4) | 121,673 (6.2) |
| BAA | 18,962 (17.7) | 65 (11.6) | 12,820 (17.3) | 5,330 (19.3) | 54 (8.4) | 693 (16.9) | 316,254 (17.1) | 335,216 (17.1) |
| NHPI | 375 (0.4) | <11 | 248 (0.3) | 106 (0.4) | <11 | 21 (0.5) | 7,656 (0.4) | 8,031 (0.4) |
| White | 70,621 (66.1) | 428 (76.2) | 49,397 (66.8) | 17,709 (64.1) | 518 (80.7) | 2,569 (62.6) | 1,066,210 (57.6) | 1,136,831 (58.1) |
| Multiple | 1,309 (1.2) | <11 | 841 (1.1) | 377 (1.4) | 11 (1.7) | 74 (1.8) | 18,421 (1.0) | 19,730 (1.0) |
| Unknown/Missing | 10,534 (9.9) | 46 (8.2) | 7,175 (9.7) | 2,854 (10.3) | 37 (5.8) | 422 (10.3) | 302,234 (16.3) | 312,768 (16.0) |
| **Ethnicity** |  |  |  |  |  |  |  |  |
| Hispanic | 21,302 (19.9) <sup>a</sup> | 74 (13.2) <sup>a</sup> | 14,356 (19.4) <sup>a</sup> | 5,948 (21.5) <sup>a</sup> | 78 (12.1) <sup>a</sup> | 846 (20.6) <sup>a</sup> | 632,780 (34.2) | 654,082 (33.4) |
| Non-Hispanic | 77,323 (72.4) | 445 (79.2) | 53,891 (72.9) | 19,513 (70.6) | 515 (80.2) | 2,959 (72.1) | 1,049,082 (56.7) | 1,126,405 (57.5) |
| Unknown/Missing | 8,232 (7.7) | 43 (7.7) | 5,672 (7.7) | 2,170 (7.9) | 49 (7.6) | 298 (7.3) | 169,056 (9.1) | 177,288 (9.1) |
| **ESHB** | 0.59 | 0.57 | 0.59 | 0.60 | 0.56 | 0.63 | 0.59 | 0.59 |
|  | [0.34, 0.79]<sup>b</sup> | [0.31, 0.78] | [0.33, 0.79]<sup>b</sup> | [0.34, 0.80] | [0.30, 0.74]<sup>b</sup> | [0.40, 0.75]<sup>b</sup> | [0.33, 0.80] | [0.33, 0.80] |
| Missing | 8,261 (7.7) | 49 (8.7) | 5,838 (7.9) | 2,135 (7.7) | 57 (8.9) | 182 (4.4) | 161,990 (8.8) | 170,251 (8.7) |
| **EBM** | 0.43 | 0.40 | 0.42 | 0.47 | 0.30 | 0.38 | 0.52 | 0.52 |
|  | [0.20, 0.71]<sup>b</sup> | [0.15, 0.68]<sup>b</sup> | [0.20, 0.71]<sup>b</sup> | [0.22, 0.73]<sup>b</sup> | [0.13, 0.55]<sup>b</sup> | [0.20, 0.68]<sup>b</sup> | [0.26, 0.75] | [0.25, 0.75] |
| Missing | 8,261 (7.7) | 49 (8.7) | 5,838 (7.9) | 2,135 (7.7) | 57 (8.9) | 182 (4.4) | 161,990 (8.8) | 170,251 (8.7) |
| **SVM** | 0.67 | 0.63 | 0.67 | 0.67 | 0.67 | 0.71 | 0.69 | 0.68 |
|  | [0.44, 0.85]<sup>b</sup> | [0.41, 0.83]<sup>b</sup> | [0.44, 0.85]<sup>b</sup> | [0.43, 0.85]<sup>b</sup> | [0.45, 0.81] | [0.51, 0.87]<sup>b</sup> | [0.43, 0.87] | [0.43, 0.86] |
| Missing | 8,261 (7.7) | 49 (8.7) | 5,838 (7.9) | 2,135 (7.7) | 57 (8.9) | 182 (4.4) | 161,990 (8.8) | 170,251 (8.7) |
| **HVM** | 0.40 | 0.40 | 0.40 | 0.40 | 0.40 | 0.40 | 0.20 | 0.20 |
|  | [0.00, 0.60]<sup>b</sup> | [0.00, 0.60]<sup>b</sup> | [0.00, 0.60]<sup>b</sup> | [0.00, 0.60]<sup>b</sup> | [0.20, 0.60]<sup>b</sup> | [0.20, 0.60]<sup>b</sup> | [0.00, 0.60] | [0.00, 0.60] |
| Missing | 8,261 (7.7) | 49 (8.7) | 5,838 (7.9) | 2,135 (7.7) | 57 (8.9) | 182 (4.4) | 161,990 (8.8) | 170,251 (8.7) |

Abbreviations: SDB=Sleep disordered breathing, CSA=Central Sleep Apnea, OSA=Obstructive Sleep Apnea, OUSA=Unspecified Sleep Apnea, AI/AN=American Indian/Alaskan Native, NHPI=Native Hawaiian/Pacific Islander, ESHB=Environmental, Social, and Health Burden, EBM=Environmental Burden Module, SVM=Social Vulnerability Module, HVM=Health Vulnerability Module.
Data are presented as n (%) or median [inter-quartile range (IQR)].
<sup>a</sup> Significant p-values are from chi-square tests between Overall SDB (or its subtypes) and No SDB.
<sup>b</sup> Significant p-values are from Mann-Whitney tests between Overall SDB (or its subtypes) and No SDB.

**Table 2.** Lifestyle and clinical characteristics of study patients, overall and by sleep-disordered breathing (SDB) status (n=1,957,775)

|  | Any SDB | CSA | OSA | OUSA | Multiple sleep apneas | Procedure-based cases | No SDB | Overall |
| --- | --- | --- | --- | --- | --- | --- | --- | --- |
| **N** | 10,6857 (5.5) | 562 (0.03) | 73,919 (3.78) | 27,631 (1.41) | 642 (0.03) | 4,103 (0.21) | 1,850,918 (94.5) | 1,957,775 (100) |
| **Body Mass Index** |  |  |  |  |  |  |  |  |
| Recommended (<25 kg/m²) | 6,203 (5.8) <sup>a</sup> | 53 (9.4) <sup>a</sup> | 3,449 (4.7) <sup>a</sup> | 1,978 (7.2) <sup>a</sup> | 45 (7.0) <sup>a</sup> | 678 (16.5) <sup>a</sup> | 344,252 (18.6) | 350,455 (17.9) |
| Overweight (25-30 kg/m²) | 16,162 (15.1) | 123 (21.9) | 10,338 (14.0) | 4,556 (16.5) | 122 (19.0) | 1,023 (24.9) | 426,567 (23.0) | 442,729 (22.6) |
| Obesity (>30 kg/m²) | 63,643 (59.6) | 267 (47.5) | 45,865 (62.0) | 15,413 (55.8) | 372 (57.9) | 1,726 (42.1) | 548,785 (29.6) | 612,428 (31.3) |
| Unknown/Missing | 20,849 (19.5) | 119 (21.2) | 14,267 (19.3) | 5,684 (20.6) | 103 (16.0) | 676 (16.5) | 531,314 (28.7) | 552,163 (28.2) |
| **Physical activity** |  |  |  |  |  |  |  |  |
| <10 minutes | 3,529 (3.3) <sup>a</sup> | 15 (2.7) <sup>a</sup> | 2,520 (3.4) <sup>a</sup> | 891 (3.2) <sup>a</sup> | 29 (4.5) <sup>a</sup> | 74 (1.8) <sup>a</sup> | 33,496 (1.8) | 37,025 (1.9) |
| 10-150 minutes | 2,473 (2.3) | 13 (2.3) | 1,703 (2.3) | 662 (2.4) | 20 (3.1) | 75 (1.8) | 24,812 (1.3) | 27,285 (1.4) |
| >150 minutes | 1,774 (1.7) | 17 (3.0) | 1,180 (1.6) | 523 (1.9) | <11 | 44 (1.1) | 25,567 (1.4) | 27,341 (1.4) |
| Missing | 99,081 (92.7) | 517 (92.0) | 68,516 (92.7) | 25,555 (92.5) | 583 (90.8) | 3,910 (95.3) | 1,767,043 (95.5) | 1,866,124 (95.3) |
| **Smoking status** |  |  |  |  |  |  |  |  |
| Current smoker | 18,054 (16.9) <sup>a</sup> | 98 (17.4) <sup>a</sup> | 11,790 (15.9) <sup>a</sup> | 5,309 (19.2) <sup>a</sup> | 113 (17.6) <sup>a</sup> | 744 (18.1) <sup>a</sup> | 252,909 (13.7) | 270,963 (13.8) |
| Former smoker | 25,386 (23.8) | 149 (26.5) | 18,317 (24.8) | 5,690 (20.6) | 206 (32.1) | 1,024 (25.0) | 200,020 (10.8) | 225,406 (11.5) |
| Never smoker | 49,369 (46.2) | 232 (41.3) | 34,501 (46.7) | 12,449 (45.1) | 249 (38.8) | 1,938 (47.2) | 941,574 (50.9) | 990,943 (50.6) |
| Unknown/missing | 14,048 (13.1) | 83 (14.8) | 9,311 (12.6) | 4,183 (15.1) | 74 (11.5) | 397 (9.7) | 456,415 (24.7) | 470,463 (24.0) |
| **Alcohol/substance use disorders** |  |  |  |  |  |  |  |  |
| No alcohol/substance use noted | 78,605 (73.6) <sup>a</sup> | 374 (66.5) <sup>a</sup> | 54,630 (73.9) <sup>a</sup> | 20,494 (74.2) <sup>a</sup> | 376 (58.6) <sup>a</sup> | 2,731 (66.6) <sup>a</sup> | 1,594,139 (86.1) | 1,672,744 (85.4) |
| Alcohol/substance use | 28,252 (26.4) | 188 (33.5) | 19,289 (26.1) | 7,137 (25.8) | 266 (41.4) | 1,372 (33.4) | 256,779 (13.9) | 285,031 (14.6) |
| **Hypertension status** |  |  |  |  |  |  |  |  |
| No hypertension | 19,689 (18.4) <sup>a</sup> | 114 (20.3) <sup>a</sup> | 12,653 (17.1) <sup>a</sup> | 5,760 (20.8) <sup>a</sup> | 119 (18.5) <sup>a</sup> | 1,043 (25.4) <sup>a</sup> | 692,886 (37.4) | 712,575 (36.4) |
| Hypertension | 84,216 (78.8) | 424 (75.4) | 59,337 (80.3) | 20,930 (75.7) | 516 (80.4) | 3,009 (73.3) | 948,393 (51.2) | 1,032,609 (52.7) |
| Missing | 2,952 (2.8) | 24 (4.3) | 1,929 (2.6) | 941 (3.4) | <11 | 51 (1.2) | 209,639 (11.3) | 212,591 (10.9) |
| **Diabetes status** |  |  |  |  |  |  |  |  |
| No diabetes | 60,891 (57.0)<sup>a</sup> | 364 (64.8)<sup>a</sup> | 40,352 (54.6)<sup>a</sup> | 17,339 (62.8)<sup>a</sup> | 339 (52.8)<sup>a</sup> | 2,497 (60.9)<sup>a</sup> | 1,545,817 (83.5) | 1,606,708 (82.1) |
| Diabetes | 45,966 (43.0) | 198 (35.2) | 33,567 (45.4) | 10,292 (37.2) | 303 (47.2) | 1,606 (39.1) | 305,101 (16.5) | 351,067 (17.9) |

Abbreviations: SDB=Sleep disordered breathing, CSA=Central Sleep Apnea, OSA=Obstructive Sleep Apnea, OUSA=Other/unspecified Sleep Apnea,
BMI=Body Mass Index.
Data are presented as n (%).
<sup>a</sup> Significant p-values are from chi-square tests between Overall SDB (or its subtypes) and No SDB.

### Potential effect modifiers

Age category, sex, race, and ethnicity were potential effect modifiers.

### Statistical analysis

We summarized (1) SDB prevalence and (2) sociodemographic, environmental, behavioral, and clinical characteristics in the overall population and by SDB status. Chi-square tests or Mann-Whitney U tests were used to compare individuals with SDB and its subtypes to individuals without SDB.

Intra-cluster correlations were checked and were negligible; therefore, we used simpler individual-level versus multilevel modeling. We used log-binomial regression to estimate prevalence ratios (PR) and 95% confidence intervals (CI) for associations between a 0.1-unit increase in ESHB, EBM, SVM, and HVM percentiles ranks with SDB and its subtypes. Three models for each were estimated: Model 1 was unadjusted; Model 2 was adjusted for age category; and Model 3 was fully adjusted for age category, sex, race, and ethnicity. Models were then stratified by age category, sex, race, and ethnicity. Two-way interaction terms between each modifier and ESHB, EBM, SVM, and HVM were added separately to fully adjusted models. To assess effect modification on the multiplicative scale, likelihood ratio tests were used to compare models with and without interaction terms.

In sensitivity analyses, we adjusted for multiple comparisons using Bonferroni-corrected p-values. Separately, we further excluded patients with a history of alcohol or substance use, as studies suggest a positive association between substance abuse and sleep disorders. ^45,46,47^ All statistical analyses were conducted using R version 4.4.3.

## RESULTS

### Patient characteristics

Among a total of 1,957,775 patients, the median age was 43.0 years (interquartile range [IQR]: 30.0-58.0; Table 1). Patients aged ≥50 years comprised 39.1% of the sample. Most were women (59.3%) and identified as White (58.1%), followed by BAA (17.1%), Asian (6.2%), AI/AN (1.2%), multiracial (1.0%), and NHPI (0.4%). Over half of patients (57.5%) identified as non-Hispanic. The documented prevalence of any SDB was 5.5%, with CSA at 0.03%, OSA at 3.8%, OUSA at 1.4%, multiple sleep apneas at 0.03%, and procedure-based cases at 0.21% The median value for ESHB was 0.59 (IQR: 0.33-0.80). ESHB was highest among procedure-based SDB cases, followed by OUSA cases, but was similar across patients with other SDB subtypes and patients without SDB. Other patient characteristics are shown in Table 2 and further described in supplemental material.

### Environmental, social, and health burden and prevalence of SDB

Each 0.1-unit increase in ESHB percentile rank was associated with a 1% higher prevalence of SDB (PR=1.01 [1.01–1.01]), OUSA (PR=1.01 [1.01–1.02]), and 5% higher prevalence of procedure-based cases (PR=1.05 [1.03–1.06]; Table 3). EBM was associated with a lower prevalence of any SDB, OSA, OUSA, multiple sleep apneas, and procedure-based cases. SVM and HVM were associated with a higher prevalence of SDB, OSA, multiple apneas, and procedure-based cases, with HVM additionally linked to higher OUSA prevalence. ESHB and its submodules were not associated with CSA. Details and results for associations with the three individual submodules (eTable 5 to eTable 7) are further described in the supplemental text.

**Table 3.**
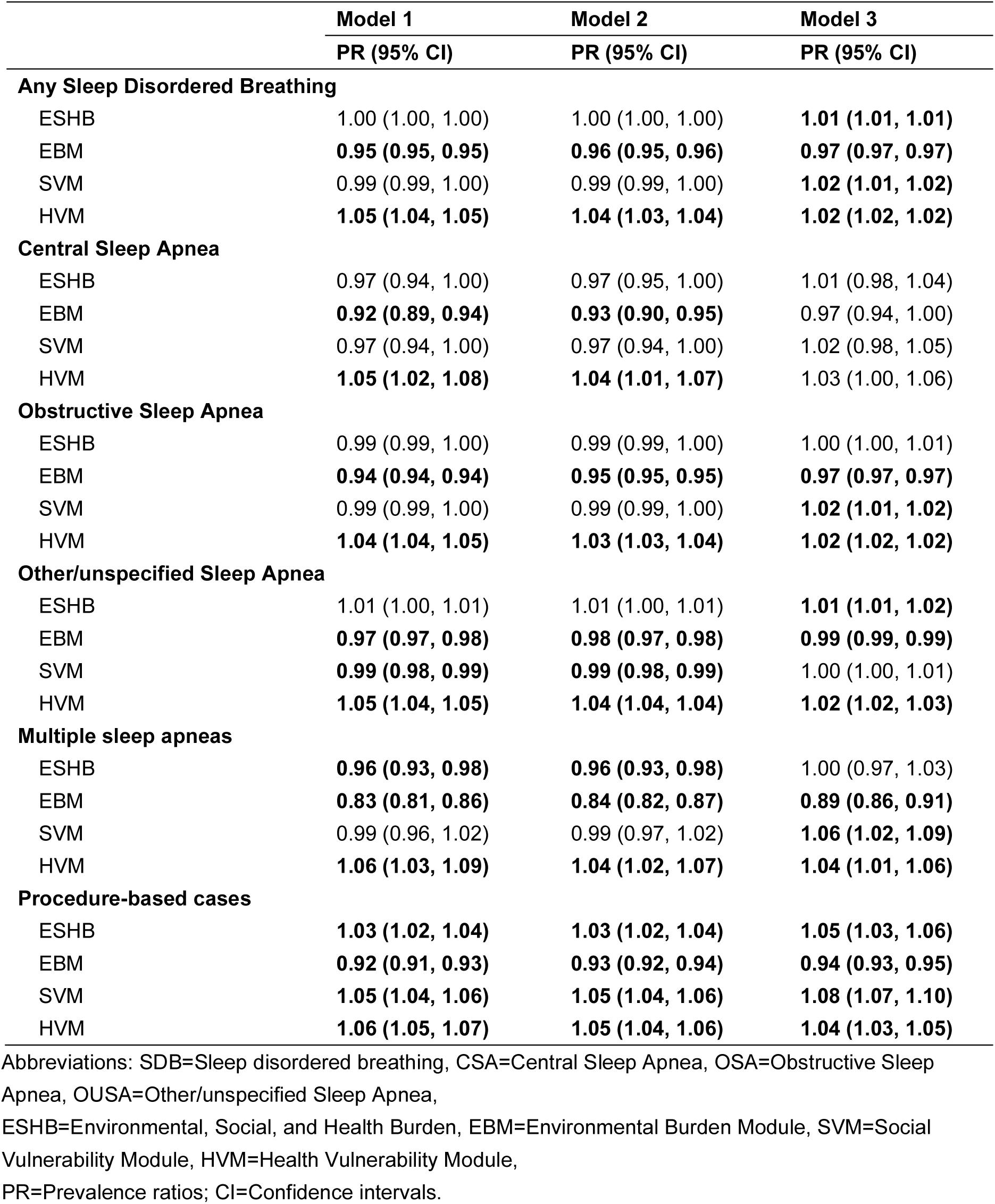
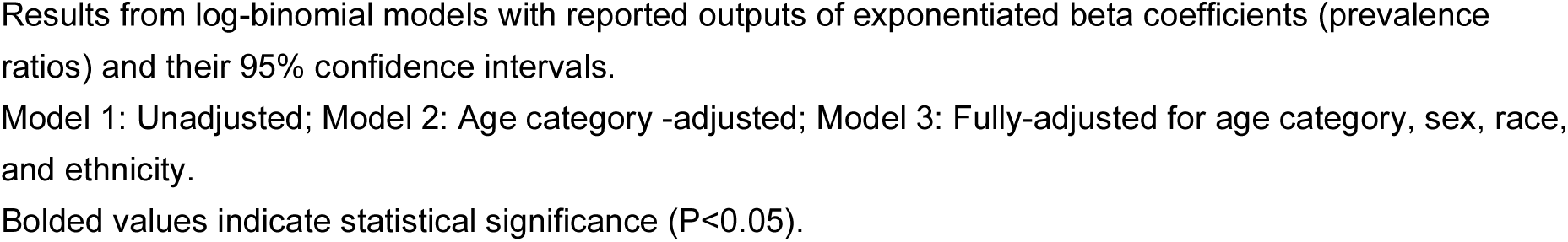
Cross-sectional associations of per 0.1-unit increase in Environmental, Social, and Health Burden (ESHB), Environmental Burden Module (EBM), Social Vulnerability Module (SVM), and Health Vulnerability Module (HVM) ranks with sleep-disordered breathing (SDB), central sleep apnea (CSA), obstructive sleep apnea (OSA), other/unspecified sleep apnea (OUSA), multiple sleep apneas, and procedure-based cases

|  | **Model 1** | **Model 2** | **Model 3** |
| --- | --- | --- | --- |
|  | **PR (95% CI)** | **PR (95% CI)** | **PR (95% CI)** |
| **Any Sleep Disordered Breathing** |  |  |  |
| ESHB | 1.00 (1.00, 1.00) | 1.00 (1.00, 1.00) | **1.01 (1.01, 1.01)** |
| EBM | **0.95 (0.95, 0.95)** | **0.96 (0.95, 0.96)** | **0.97 (0.97, 0.97)** |
| SVM | 0.99 (0.99, 1.00) | 0.99 (0.99, 1.00) | **1.02 (1.01, 1.02)** |
| HVM | **1.05 (1.04, 1.05)** | **1.04 (1.03, 1.04)** | **1.02 (1.02, 1.02)** |
| **Central Sleep Apnea** |  |  |  |
| ESHB | 0.97 (0.94, 1.00) | 0.97 (0.95, 1.00) | 1.01 (0.98, 1.04) |
| EBM | **0.92 (0.89, 0.94)** | **0.93 (0.90, 0.95)** | 0.97 (0.94, 1.00) |
| SVM | 0.97 (0.94, 1.00) | 0.97 (0.94, 1.00) | 1.02 (0.98, 1.05) |
| HVM | **1.05 (1.02, 1.08)** | **1.04 (1.01, 1.07)** | 1.03 (1.00, 1.06) |
| **Obstructive Sleep Apnea** |  |  |  |
| ESHB | 0.99 (0.99, 1.00) | 0.99 (0.99, 1.00) | 1.00 (1.00, 1.01) |
| EBM | **0.94 (0.94, 0.94)** | **0.95 (0.95, 0.95)** | **0.97 (0.97, 0.97)** |
| SVM | 0.99 (0.99, 1.00) | 0.99 (0.99, 1.00) | **1.02 (1.01, 1.02)** |
| HVM | **1.04 (1.04, 1.05)** | **1.03 (1.03, 1.04)** | **1.02 (1.02, 1.02)** |
| **Other/unspecified Sleep Apnea** |  |  |  |
| ESHB | 1.01 (1.00, 1.01) | 1.01 (1.00, 1.01) | **1.01 (1.01, 1.02)** |
| EBM | **0.97 (0.97, 0.98)** | **0.98 (0.97, 0.98)** | **0.99 (0.99, 0.99)** |
| SVM | **0.99 (0.98, 0.99)** | **0.99 (0.98, 0.99)** | 1.00 (1.00, 1.01) |
| HVM | **1.05 (1.04, 1.05)** | **1.04 (1.04, 1.04)** | **1.02 (1.02, 1.03)** |
| **Multiple sleep apneas** |  |  |  |
| ESHB | **0.96 (0.93, 0.98)** | **0.96 (0.93, 0.98)** | 1.00 (0.97, 1.03) |
| EBM | **0.83 (0.81, 0.86)** | **0.84 (0.82, 0.87)** | **0.89 (0.86, 0.91)** |
| SVM | 0.99 (0.96, 1.02) | 0.99 (0.97, 1.02) | **1.06 (1.02, 1.09)** |
| HVM | **1.06 (1.03, 1.09)** | **1.04 (1.02, 1.07)** | **1.04 (1.01, 1.06)** |
| **Procedure-based cases** |  |  |  |
| ESHB | **1.03 (1.02, 1.04)** | **1.03 (1.02, 1.04)** | **1.05 (1.03, 1.06)** |
| EBM | **0.92 (0.91, 0.93)** | **0.93 (0.92, 0.94)** | **0.94 (0.93, 0.95)** |
| SVM | **1.05 (1.04, 1.06)** | **1.05 (1.04, 1.06)** | **1.08 (1.07, 1.10)** |
| HVM | **1.06 (1.05, 1.07)** | **1.05 (1.04, 1.06)** | **1.04 (1.03, 1.05)** |

Abbreviations: SDB=Sleep disordered breathing, CSA=Central Sleep Apnea, OSA=Obstructive Sleep Apnea, OUSA=Other/unspecified Sleep Apnea, ESHB=Environmental, Social, and Health Burden, EBM=Environmental Burden Module, SVM=Social Vulnerability Module, HVM=Health Vulnerability Module, PR=Prevalence ratios; CI=Confidence intervals.
Results from log-binomial models with reported outputs of exponentiated beta coefficients (prevalence ratios) and their 95% confidence intervals.
Model 1: Unadjusted; Model 2: Age category -adjusted; Model 3: Fully-adjusted for age category, sex, race, and ethnicity.
Bolded values indicate statistical significance ( $P < 0.05$ ).

### Effect modification of associations between ESHB and SDB

#### By age

Age modified the associations between ESHB with SDB and OSA (both P_interaction_<0.01); PRs were elevated among the two youngest age groups (e.g., SDB: PR_18-34 years_=1.02 [1.01–1.03], PR_35-49 years_=1.02 [1.01–1.02]; Table 4), but no association among the oldest age group (e.g., SDB: PR≥_50 years_=1.00 [1.00–1.00]; Table 4).

**Table 4.**
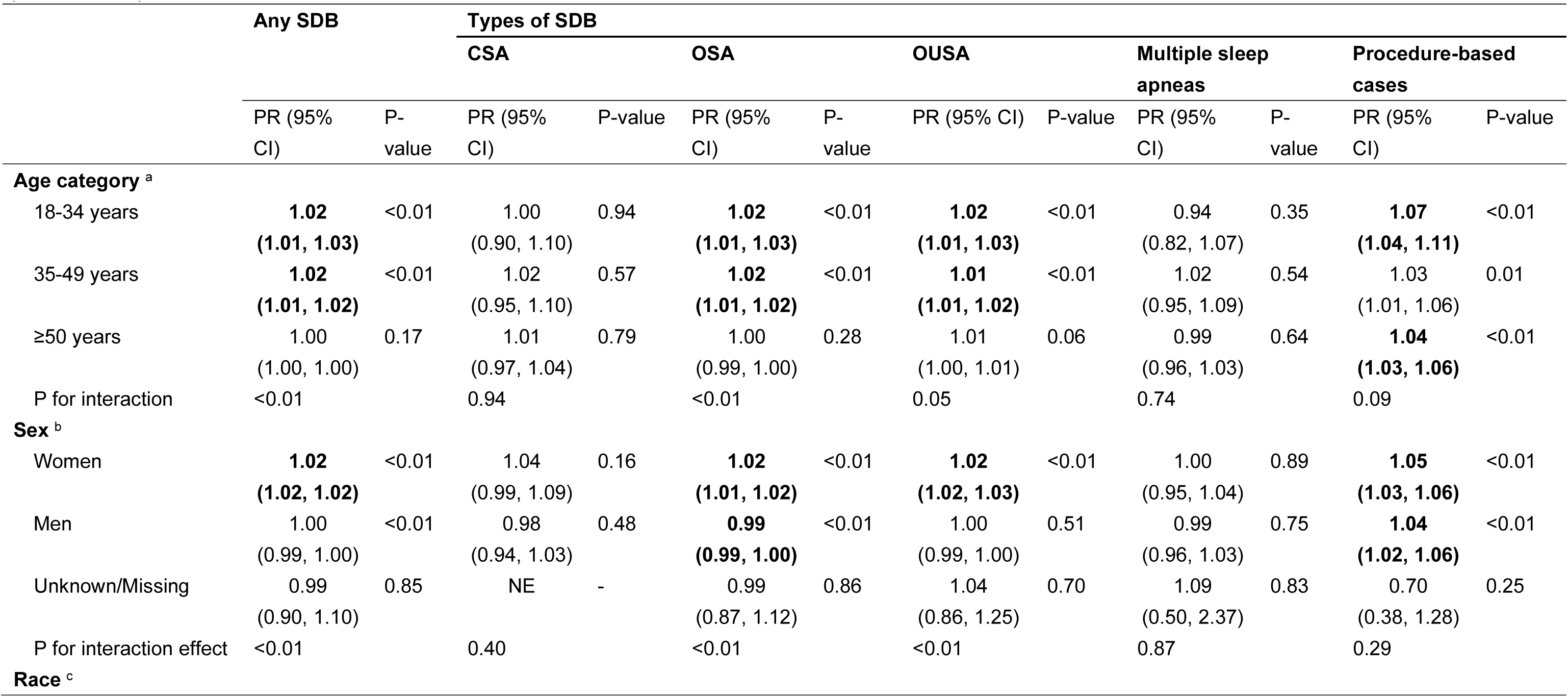
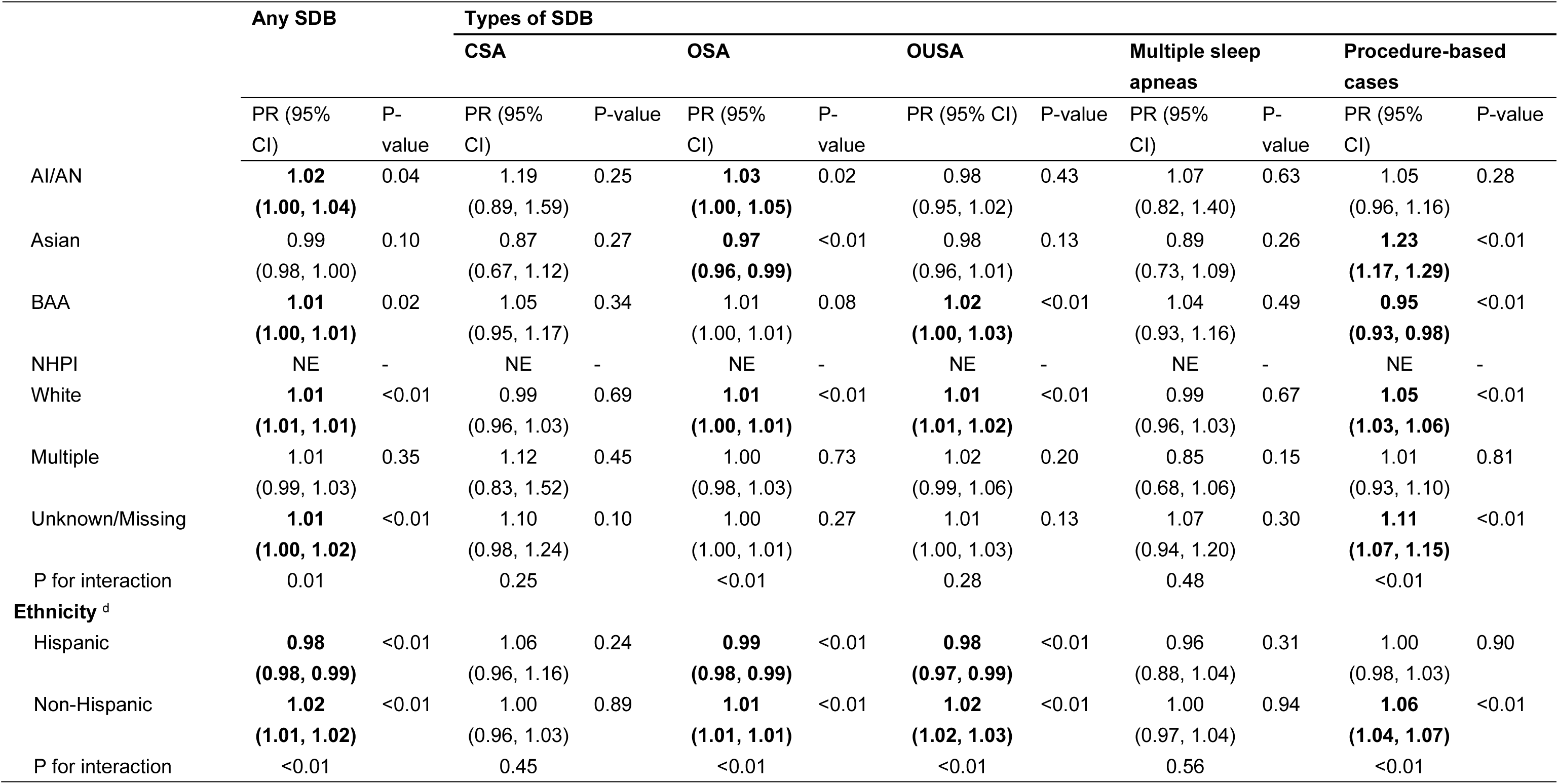
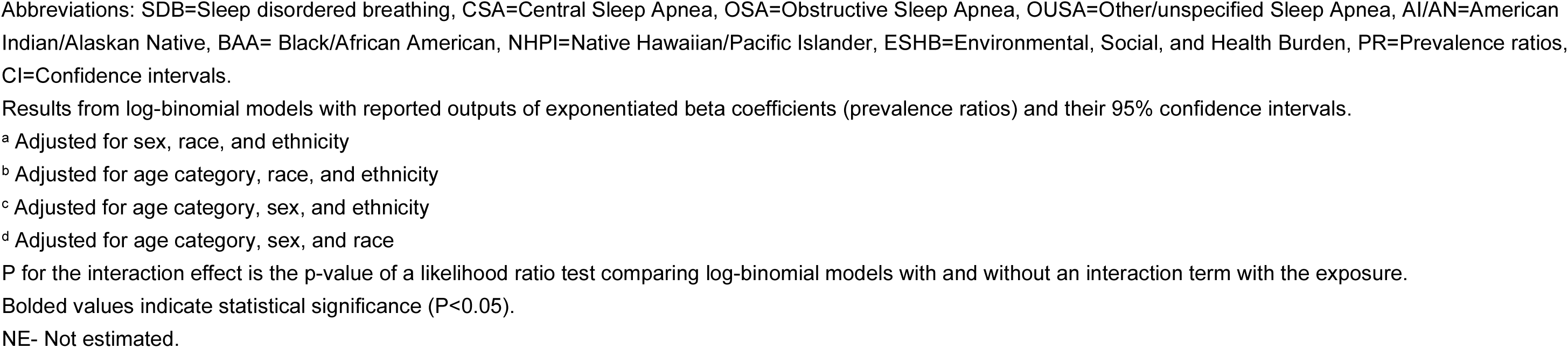
Subgroup cross-sectional associations of per 0.1-unit increase in Environmental, Social, and Health Burden (ESHB) ranks with sleep-disordered breathing (SDB), central sleep apnea (CSA), obstructive sleep apnea (OSA), and other/unspecified sleep apnea (OUSA), multiple sleep apneas, and procedure-based cases

|  | Any SDB |  | Types of SDB |  |  |  |  |  |  |  |  |  |
| --- | --- | --- | --- | --- | --- | --- | --- | --- | --- | --- | --- | --- |
|  |  |  | CSA |  | OSA |  | OUSA |  | Multiple sleep apneas |  | Procedure-based cases |  |
|  | PR (95% CI) | P-value | PR (95% CI) | P-value | PR (95% CI) | P-value | PR (95% CI) | P-value | PR (95% CI) | P-value | PR (95% CI) | P-value |
| **Age category <sup>a</sup>** |  |  |  |  |  |  |  |  |  |  |  |  |
| 18-34 years | **1.02**<br>**(1.01, 1.03)** | <0.01 | 1.00<br>(0.90, 1.10) | 0.94 | **1.02**<br>**(1.01, 1.03)** | <0.01 | **1.02**<br>**(1.01, 1.03)** | <0.01 | 0.94<br>(0.82, 1.07) | 0.35 | **1.07**<br>**(1.04, 1.11)** | <0.01 |
| 35-49 years | **1.02**<br>**(1.01, 1.02)** | <0.01 | 1.02<br>(0.95, 1.10) | 0.57 | **1.02**<br>**(1.01, 1.02)** | <0.01 | **1.01**<br>**(1.01, 1.02)** | <0.01 | 1.02<br>(0.95, 1.09) | 0.54 | 1.03<br>(1.01, 1.06) | 0.01 |
| ≥50 years | 1.00<br>(1.00, 1.00) | 0.17 | 1.01<br>(0.97, 1.04) | 0.79 | 1.00<br>(0.99, 1.00) | 0.28 | 1.01<br>(1.00, 1.01) | 0.06 | 0.99<br>(0.96, 1.03) | 0.64 | **1.04**<br>**(1.03, 1.06)** | <0.01 |
| P for interaction | <0.01 |  | 0.94 |  | <0.01 |  | 0.05 |  | 0.74 |  | 0.09 |  |
| **Sex <sup>b</sup>** |  |  |  |  |  |  |  |  |  |  |  |  |
| Women | **1.02**<br>**(1.02, 1.02)** | <0.01 | 1.04<br>(0.99, 1.09) | 0.16 | **1.02**<br>**(1.01, 1.02)** | <0.01 | **1.02**<br>**(1.02, 1.03)** | <0.01 | 1.00<br>(0.95, 1.04) | 0.89 | **1.05**<br>**(1.03, 1.06)** | <0.01 |
| Men | 1.00<br>(0.99, 1.00) | <0.01 | 0.98<br>(0.94, 1.03) | 0.48 | **0.99**<br>**(0.99, 1.00)** | <0.01 | 1.00<br>(0.99, 1.00) | 0.51 | 0.99<br>(0.96, 1.03) | 0.75 | **1.04**<br>**(1.02, 1.06)** | <0.01 |
| Unknown/Missing | 0.99<br>(0.90, 1.10) | 0.85 | NE | - | 0.99<br>(0.87, 1.12) | 0.86 | 1.04<br>(0.86, 1.25) | 0.70 | 1.09<br>(0.50, 2.37) | 0.83 | 0.70<br>(0.38, 1.28) | 0.25 |
| P for interaction effect | <0.01 |  | 0.40 |  | <0.01 |  | <0.01 |  | 0.87 |  | 0.29 |  |
| **Race <sup>c</sup>** |  |  |  |  |  |  |  |  |  |  |  |  |
|  |  |  | CSA |  | OSA |  | OUSA |  | Multiple sleep apneas |  | Procedure-based cases |  |
|  | PR (95% CI) | P-value | PR (95% CI) | P-value | PR (95% CI) | P-value | PR (95% CI) | P-value | PR (95% CI) | P-value | PR (95% CI) | P-value |
| AI/AN | **1.02**<br>**(1.00, 1.04)** | 0.04 | 1.19<br>(0.89, 1.59) | 0.25 | **1.03**<br>**(1.00, 1.05)** | 0.02 | 0.98<br>(0.95, 1.02) | 0.43 | 1.07<br>(0.82, 1.40) | 0.63 | 1.05<br>(0.96, 1.16) | 0.28 |
| Asian | 0.99<br>(0.98, 1.00) | 0.10 | 0.87<br>(0.67, 1.12) | 0.27 | **0.97**<br>**(0.96, 0.99)** | <0.01 | 0.98<br>(0.96, 1.01) | 0.13 | 0.89<br>(0.73, 1.09) | 0.26 | **1.23**<br>**(1.17, 1.29)** | <0.01 |
| BAA | **1.01**<br>**(1.00, 1.01)** | 0.02 | 1.05<br>(0.95, 1.17) | 0.34 | 1.01<br>(1.00, 1.01) | 0.08 | **1.02**<br>**(1.00, 1.03)** | <0.01 | 1.04<br>(0.93, 1.16) | 0.49 | **0.95**<br>**(0.93, 0.98)** | <0.01 |
| NHPI | NE | - | NE | - | NE | - | NE | - | NE | - | NE | - |
| White | **1.01**<br>**(1.01, 1.01)** | <0.01 | 0.99<br>(0.96, 1.03) | 0.69 | **1.01**<br>**(1.00, 1.01)** | <0.01 | **1.01**<br>**(1.01, 1.02)** | <0.01 | 0.99<br>(0.96, 1.03) | 0.67 | **1.05**<br>**(1.03, 1.06)** | <0.01 |
| Multiple | 1.01<br>(0.99, 1.03) | 0.35 | 1.12<br>(0.83, 1.52) | 0.45 | 1.00<br>(0.98, 1.03) | 0.73 | 1.02<br>(0.99, 1.06) | 0.20 | 0.85<br>(0.68, 1.06) | 0.15 | 1.01<br>(0.93, 1.10) | 0.81 |
| Unknown/Missing | **1.01**<br>**(1.00, 1.02)** | <0.01 | 1.10<br>(0.98, 1.24) | 0.10 | 1.00<br>(1.00, 1.01) | 0.27 | 1.01<br>(1.00, 1.03) | 0.13 | 1.07<br>(0.94, 1.20) | 0.30 | **1.11**<br>**(1.07, 1.15)** | <0.01 |
| P for interaction | 0.01 |  | 0.25 |  | <0.01 |  | 0.28 |  | 0.48 |  | <0.01 |  |
| **Ethnicity<sup>d</sup>** |  |  |  |  |  |  |  |  |  |  |  |  |
| Hispanic | **0.98**<br>**(0.98, 0.99)** | <0.01 | 1.06<br>(0.96, 1.16) | 0.24 | **0.99**<br>**(0.98, 0.99)** | <0.01 | **0.98**<br>**(0.97, 0.99)** | <0.01 | 0.96<br>(0.88, 1.04) | 0.31 | 1.00<br>(0.98, 1.03) | 0.90 |
| Non-Hispanic | **1.02**<br>**(1.01, 1.02)** | <0.01 | 1.00<br>(0.96, 1.03) | 0.89 | **1.01**<br>**(1.01, 1.01)** | <0.01 | **1.02**<br>**(1.02, 1.03)** | <0.01 | 1.00<br>(0.97, 1.04) | 0.94 | **1.06**<br>**(1.04, 1.07)** | <0.01 |
| P for interaction | <0.01 |  | 0.45 |  | <0.01 |  | <0.01 |  | 0.56 |  | <0.01 |  |

Abbreviations: SDB=Sleep disordered breathing, CSA=Central Sleep Apnea, OSA=Obstructive Sleep Apnea, OUSA=Other/unspecified Sleep Apnea, AI/AN=American Indian/Alaskan Native, BAA= Black/African American, NHPI=Native Hawaiian/Pacific Islander, ESHB=Environmental, Social, and Health Burden, PR=Prevalence ratios, CI=Confidence intervals.
Results from log-binomial models with reported outputs of exponentiated beta coefficients (prevalence ratios) and their 95% confidence intervals.
<sup>a</sup> Adjusted for sex, race, and ethnicity
<sup>b</sup> Adjusted for age category, race, and ethnicity
<sup>c</sup> Adjusted for age category, sex, and ethnicity
<sup>d</sup> Adjusted for age category, sex, and race
P for the interaction effect is the p-value of a likelihood ratio test comparing log-binomial models with and without an interaction term with the exposure.
Bolded values indicate statistical significance ( $P < 0.05$ ).
NE- Not estimated.

#### By sex

We observed positive associations between ESHB and prevalence of SDB, OSA, and OUSA among women but not men (all P_interaction_<0.01; e.g., SDB: PR_women_=1.02 [1.02–1.02] and PR_men_=1.00 [0.99–1.00]; OSA: PR_women_=1.02 [1.01–1.02] and PR_men_=0.99 [0.99–1.00]; OUSA: PR_women_=1.02 [1.02–1.03] and PR_men_=1.00 [0.99–1.00]; Table 4).

#### By race

There was evidence of effect modification by race in associations between ESHB and SDB, OSA, and procedure-based cases (all P_interaction_<0.01; Table 4); however, CIs largely overlapped across racial groups, with a clear effect modification observed only for procedure-based cases. For instance, although PRs among AI/AN and White patients were elevated, the 95% CIs of the ESHB-SDB and ESHB-OSA associations among AI/AN patients included one (e.g., SDB: PR_AI/AN_=1.02 [1.00–1.04] and PR_White_=1.01 [1.01–1.01]; OSA: PR_AI/AN_=1.03 [1.00–1.05] and PR_White_=1.01 [1.00–1.01]; Table 4). There was an elevated association between ESHB and prevalence of procedure-based cases among Asian patients (PR_Asian_=1.23 [1.17–1.29]), higher than White patients (PR_White_=1.05 [1.03–1.06]), whereas there was a negative association among BAA patients (PR_BAA_=0.95 [0.93–0.98]; Table 4).

#### By ethnicity

There was an inverse association between ESHB and SDB (PR_Hispanic_=0.98 [0.98–0.99]), OSA (PR_Hispanic_=0.99 [0.98–0.99]), and OUSA (PR_Hispanic_=0.98 [0.97–0.99]) among Hispanic patients; however, ESHB was positively associated with SDB (PR_non-Hispanic_=1.02 [1.01–1.02]), OSA (PR_non-Hispanic_=1.01 [1.01–1.02]), and OUSA (PR_non-Hispanic_=1.02 [1.02–1.03]) among non-Hispanic patients (all P_interaction_<0.01; Table 4). ESHB was not associated with procedure-based SDB among Hispanic patients (PR_Hispanic_=1.00 [0.98–1.03]) but was positively associated among non-Hispanic patients (PR_non-Hispanic_=1.06 [1.04–1.07]; P_interaction_<0.01; Table 4).

### Sensitivity analyses

After accounting for multiple comparisons with Bonferroni correction, the estimates of associations between ESHB and its three submodules with SDB and its subtypes did not substantially change (eTable 8-11). Excluding patients with a history of substance use did not substantially change the results (eTable 12).

## DISCUSSION

In this study, which investigates multifactorial community-level risk factors in relation to SDB, patients of community health centers across the United States, living in communities experiencing higher environmental, social, and health-related burdens or vulnerabilities, had a higher prevalence of SDB and non-specific SDB subtypes. This main finding supported our overarching hypothesis. Furthermore, the ESHB-SDB association was elevated among adults aged 18-49 years, women, AI/AN and White (despite overlapping CIs), and non-Hispanic, whereas for procedure-based cases the association was elevated among Asian adults. Separate analyses of the three modules revealed several important nuances. For instance, although patients with higher environmental burdens had lower prevalence of SDB and its subtypes, living in communities with higher social vulnerability (e.g., greater unemployment and poverty) and higher health vulnerability, reflecting higher community-level prevalence of chronic diseases (e.g., asthma), was consistently associated with a higher prevalence of SDB and its non-specific subtypes.

To our knowledge, this is the first study to investigate ESHB in relation to SDB, limiting direct comparisons to prior literature; however, several studies of ESHB and risk factors for SDB support our findings. One study from the CDC’s 2022 Population-Level Analysis and Community Estimates (PLACES; providing chronic disease and health-related data across all U.S. counties) found that adults in the highest vs. lowest ESHB quartile had higher prevalence of SDB risk factors such as obesity. ^36^ In a study of 71,677 U.S. neighborhoods using the 2023 CDC PLACES dataset, adults in the highest vs. lowest quartiles of ESHB had higher asthma prevalence; ^35^ this finding is particularly relevant given the established link between asthma and SDB. ^48^

Multiple mechanisms likely underlie the association between ESHB and SDB. ESHB combines harmful community-level exposures in the physical, social (including socioeconomic), and health environments. Although environmental burden was not linked to SDB in the expected direction, plausible mechanisms remain. For instance, air pollution is believed to contribute to SDB risk by increasing respiratory disease burden, triggering systemic inflammation, and raising susceptibility to infections. ^49^ Factors in the built environment, such as limited residential neighborhood walkability, are linked to higher obesity, increasing SDB risk through visceral fat accumulation. ^28,32^ Social, particularly socioeconomic, deprivation likely impacts SDB through mechanisms such as limited access to resources. Disinvestment in low-SES neighborhoods can also lead to poor infrastructure and increased exposure to environmental hazards. ^50^ Residential segregation may further limit access to education and income opportunities, increasing susceptibility to SDB through psychosocial stress, which elevates proinflammatory cytokines that impair upper airway dilator activity. ^51,52^ Pre-existing health conditions may disproportionately burden medically underserved communities, influencing SDB prevalence. For instance, asthma may worsen airway obstruction and contribute to SDB. ^53^ Other chronic conditions that are often not well-managed, such as diabetes and hypertension (highly prevalent in patients with SDB outcomes), may exacerbate SDB severity through metabolic dysregulation and vascular stress. ^54^

Previous studies on this topic lacked subgroup analyses, limiting direct comparison to our study; however, the distribution of ESHB across age, sex, race, and ethnicity in the U.S. general population partially explains our observations. ^35,36^ Prior findings suggest that subgroups disproportionately bear community-level burdens that may have increased vulnerability to environmental influences on SDB. Specifically, data from 71,677 U.S. census tracts (∼320 million adults) showed that higher ESHB quartiles comprised larger proportions of adults aged 18-44 years and women. ^35^ In that study, the proportion of AI/AN adults increased from the first to third quartiles of ESHB but slightly declined in the highest, while White individuals decreased as ESHB quartiles increased. ^35^ There was also a higher proportion of Asian individuals in the fourth versus the second and third ESHB quartiles. ^35^ In our study, the stronger associations between ESHB and specific SDB outcomes (i.e., SDB and procedure-based cases) among younger (aged 18-49 years) and AI/AN and Asian adults may be partially explained by the higher likelihood of living in census tracts with higher ESHB among the groups showing stronger associations. Regarding the opposing associations observed among non-Hispanic and Hispanic patients (i.e., PRs >1 and <1) in our sample, the non-Hispanic group included heterogeneous racial and ethnic groups with varying exposures and baseline SDB risk, which impacts the interpretability of the results. Additionally, the data comprise a low-SES population, which limits generalizability to socioeconomically diverse populations in which patterns of associations may differ from the patterns we observed.

Regarding the unexpected association between higher environmental burdens and lower SDB prevalence, in our sample, median EBM scores were lower among those with vs. without SDB. This may reflect the characteristics of our medically underserved, non-nationally representative sample, where this group is mainly exposed to adverse environmental burden with limited variability, making it difficult to detect associations. Additionally, unmeasured neighborhood characteristics could explain these findings. For example, greenspace that has been found to differ by race and ethnicity was unmeasured but may be an important factor influencing associations with SDB. ^14,55,56^ Moreover, certain EBM indicators, like the proportion of census tract area near hazardous/toxic sites (see Figure 1), may not accurately reflect individuals’ direct exposure to these hazards. Some EBM components (e.g., water pollution) may have limited relevance to SDB pathophysiology. ^14^ Environmental burden is also influenced by social vulnerability. ^24^ It is also possible that social vulnerability may play a dominant role, potentially masking environmental influences on SDB.

As mentioned, this is the first study, to our knowledge, to investigate multifactorial community-level determinants of SDB, using data from over one million patients representing medically underserved populations across the U.S. Another strength is that the ESHB captures the complexity of community-level multifactorial physical, social, and health burden risk factors for SDB beyond individual factors. We distinguished OSA, CSA, and multiple apnea types from non-specific subtypes using EHR data, and the use of individual-level outcomes and covariates reduced confounding and improved precision.

### Limitations

The cross-sectional study design limits causal inferences. Although using census tract-level (vs. individual-level) ESHB data may introduce bias, data remain valuable for capturing multiple exposures. Future studies could enhance the precision of exposure assessment by incorporating individual-level exposure data, such as from wearable devices. ^4,57,58^ The study only captured cases that sought and received care within the OCHIN network, limiting generalizability. Nonetheless, communities from across the United States were included. Prospective studies with broader populations are needed to fully elucidate the influences of multifactorial environmental factors on SDB.

## CONCLUSIONS

Higher overall community-level environmental, social, and health burden was associated with a higher prevalence of SDB and non-specific SDB subtypes (i.e., OUSA and procedure-based cases) among a medically underserved population. ESHB-SDB associations were stronger in adults aged 18-49 vs. ≥50 years, women vs. men, AI/AN vs. White, and non-Hispanic vs. Hispanic adults. While further prospective research is needed, our findings underscore the importance of addressing community-level burdens in medically underserved populations while considering the prioritization of resources to reduce the burden of SDB, a prevalent sleep disorder with potentially severe health consequences.

## Data Availability

All data produced in the present study are available upon reasonable request to the authors

## Author Contributions

**Authors:** WS. Zhou, SA. Gaston, R. Singh, K. Konrad, I. Buller, WB Jackson, D. Neo, R. Voss, A. Steeves-Reece, PD. Juarez, CL. Jackson

Study concept: CL. Jackson.

Study design: CL. Jackson, SA. Gaston, R. Singh.

Acquisition of data: I. Buller, WB Jackson, D. Neo, R. Voss, A. Steeves-Reece, PD. Juarez, CL. Jackson

Statistical Analysis: I. Buller, K. Konrad, D. Neo

Interpretation of data: WS. Zhou, SA. Gaston, R. Singh, I. Buller, WB Jackson, K. Konrad, D. Neo, R. Voss, A. Steeves-Reece, PD. Juarez, CL. Jackson

Drafting of the manuscript: WS. Zhou.

Critical revision of the manuscript for important intellectual content: WS. Zhou, SA. Gaston, R. Singh, I. Buller, WB Jackson, K. Konrad, D. Neo, R. Voss, A. Steeves-Reece, PD. Juarez, CL. Jackson

Administrative, technical, and material support: WB Jackson, R. Voss, A. Steeves-Reece, PD. Juarez, CL. Jackson

Obtaining funding: CL. Jackson.

Study supervision: CL. Jackson.

Final Approval: WS. Zhou, SA. Gaston, R. Singh, K. Konrad, I. Buller, F. LaPorte, D. Neo, R. Voss, A. Steeves-Reece, PD. Juarez, CL. Jackson

## Disclaimer

The contributions of the NIH authors were made as part of their official duties as NIH federal employees, are in compliance with agency policy requirements, and are considered Works of the United States Government. However, the findings and conclusions presented in this paper are those of the authors and do not necessarily reflect the views of the NIH or the U.S. Department of Health and Human Services.

## Data Sharing Statement

Data available upon reasonable request.

## Acknowledgements

The research reported in this work was powered by PCORnet® and conducted with the Accelerating Data Value Across a National Community Health Center Network (ADVANCE) Clinical Research Network. PCORnet was developed with funding from the Patient-Centered Outcomes Research Institute®. The research reported in this work was powered by PCORnet®. PCORnet has been developed with funding from the Patient-Centered Outcomes Research Institute® (PCORI®) and conducted with the Accelerating Data Value Across a National Community Health Center Network (ADVANCE) Clinical Research Network (CRN). ADVANCE is a Clinical Research Network in PCORnet® led by OCHIN in partnership with Health Choice Network, Fenway Health, University of Washington, and Oregon Health & Science University. ADVANCE’s participation in PCORnet® is funded through the PCORI Award RI-OCHIN-01-MC.

## Supplemental Materials

**eFigure 1.**
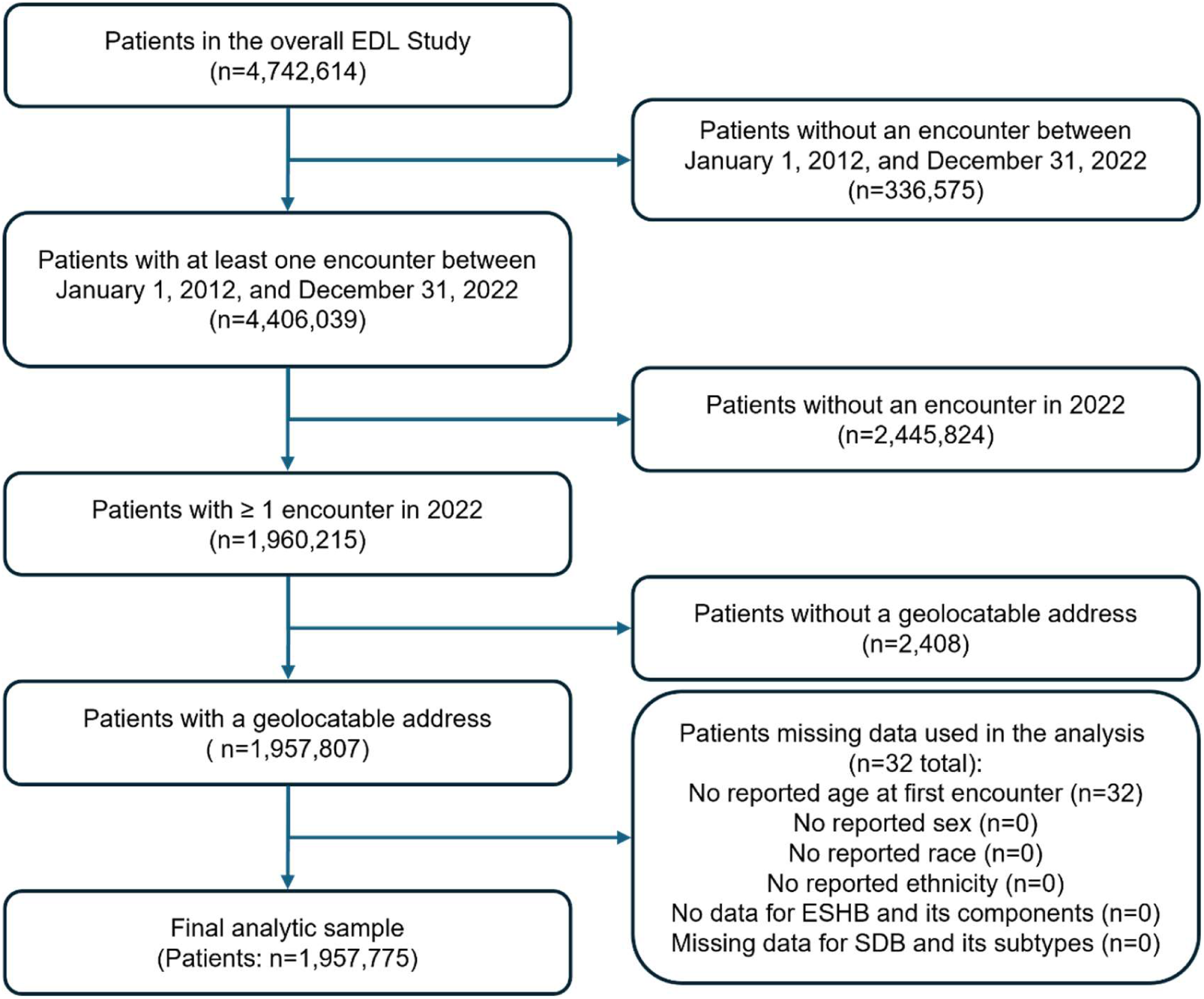
A flowchart of the study inclusions/exclusion criteria of the association between Environmental, Social, and Health Burden (ESHB) and sleep-disordered breathing (SDB)

**eFigure 2.**
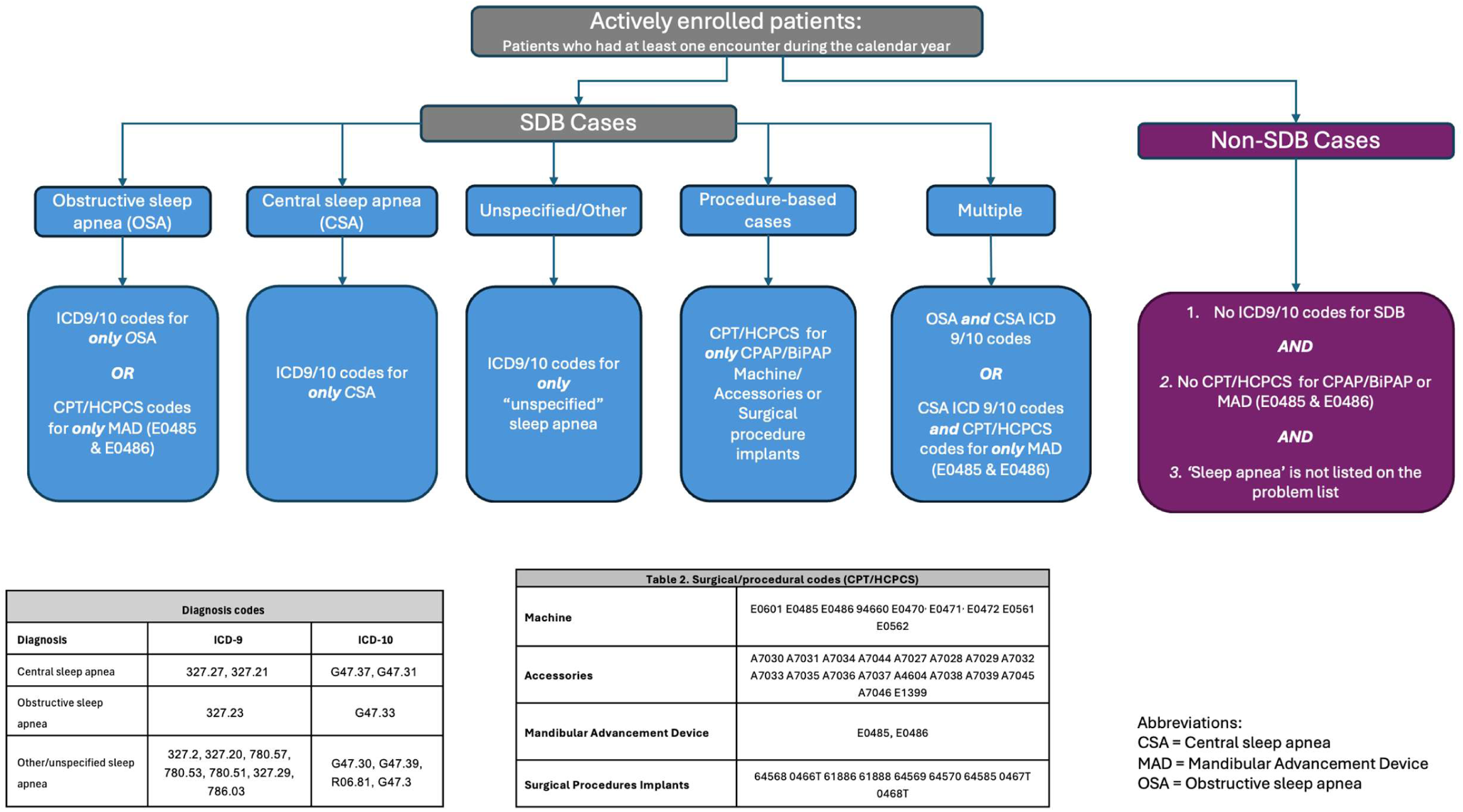
Flowchart of sample selection

**eFigure 3.**
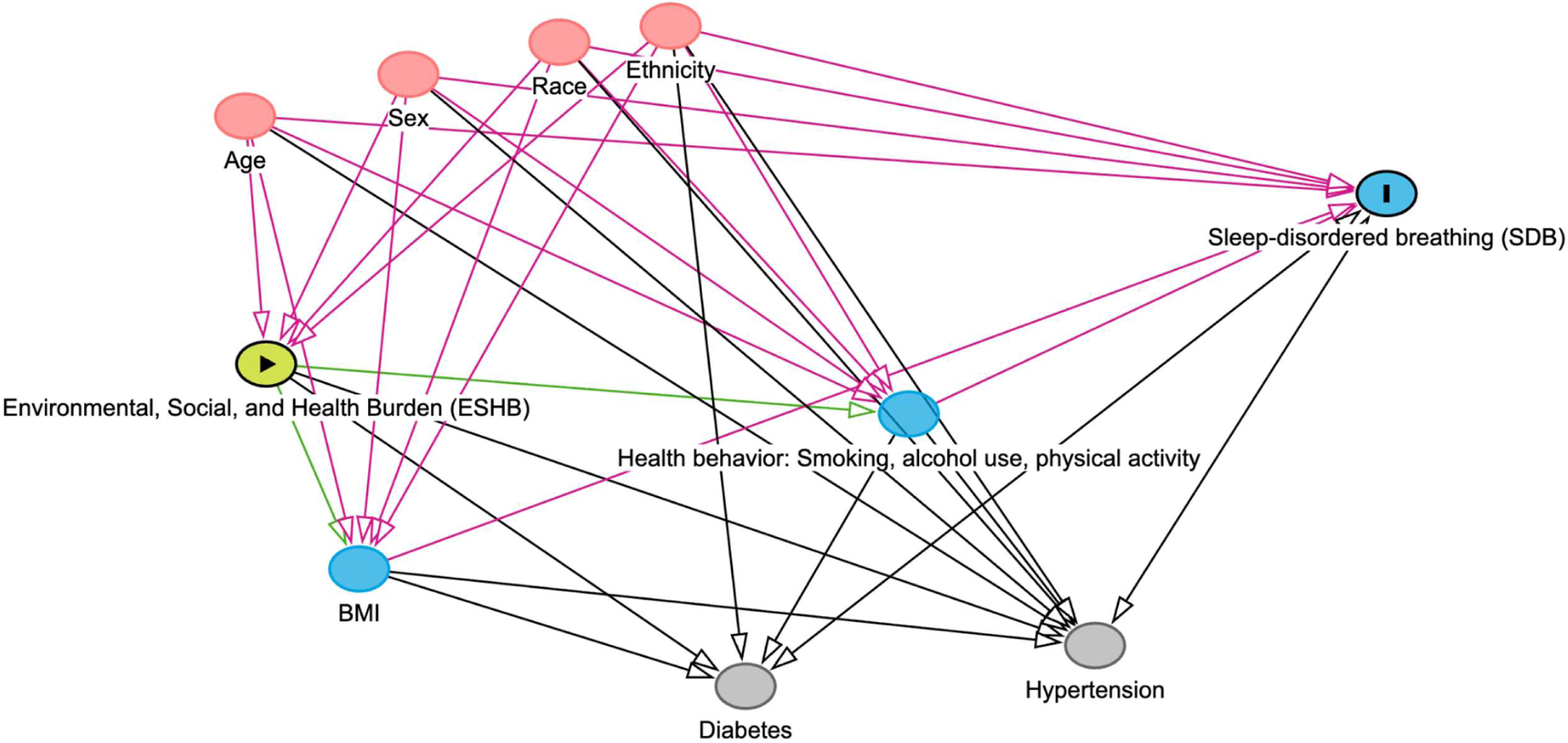
Directed Acyclic Graph (DAG) for the association between Environmental, Social, and Health Burden (ESHB) and sleep-disordered breathing (SDB)

**eTable 1.**
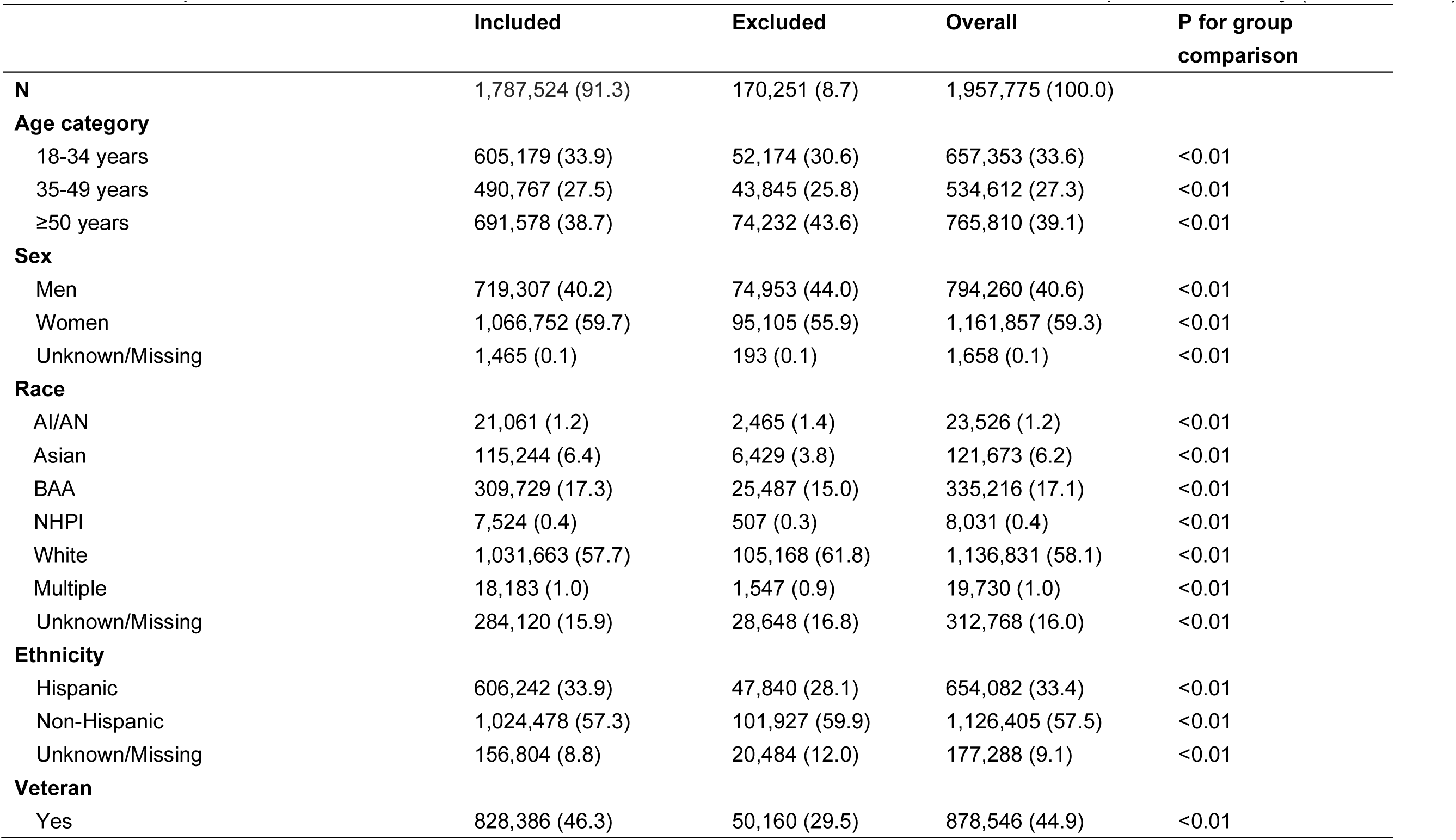
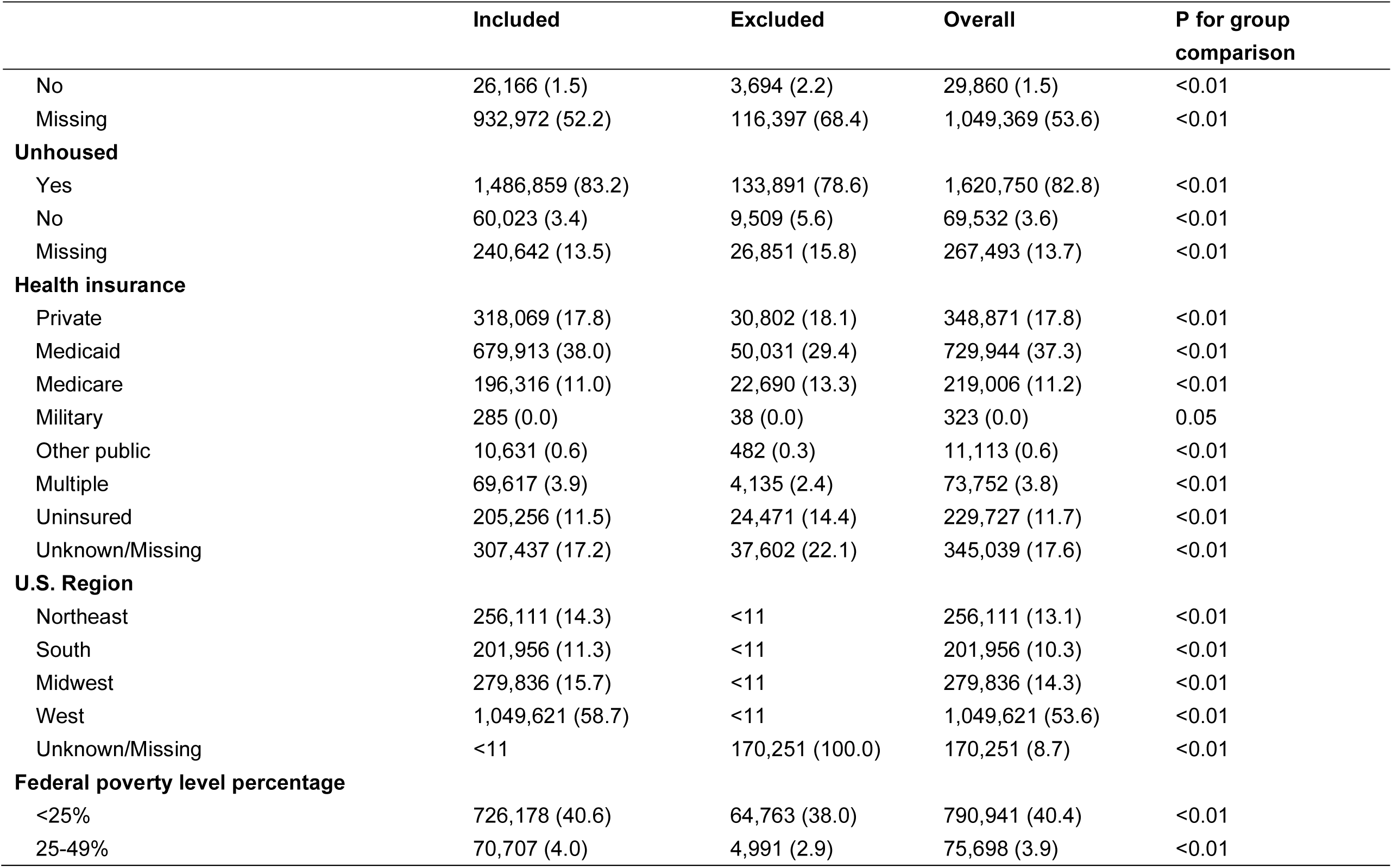
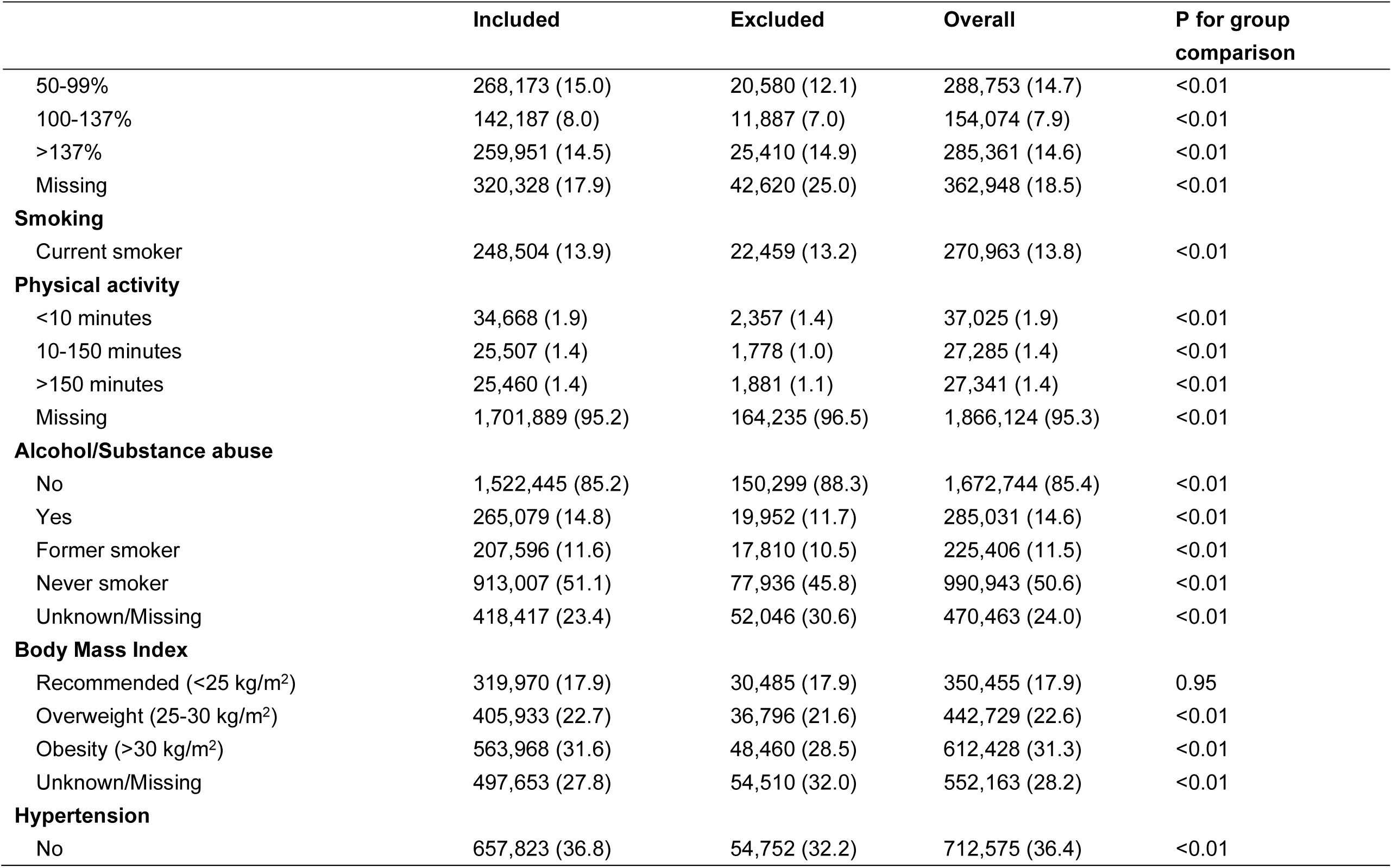
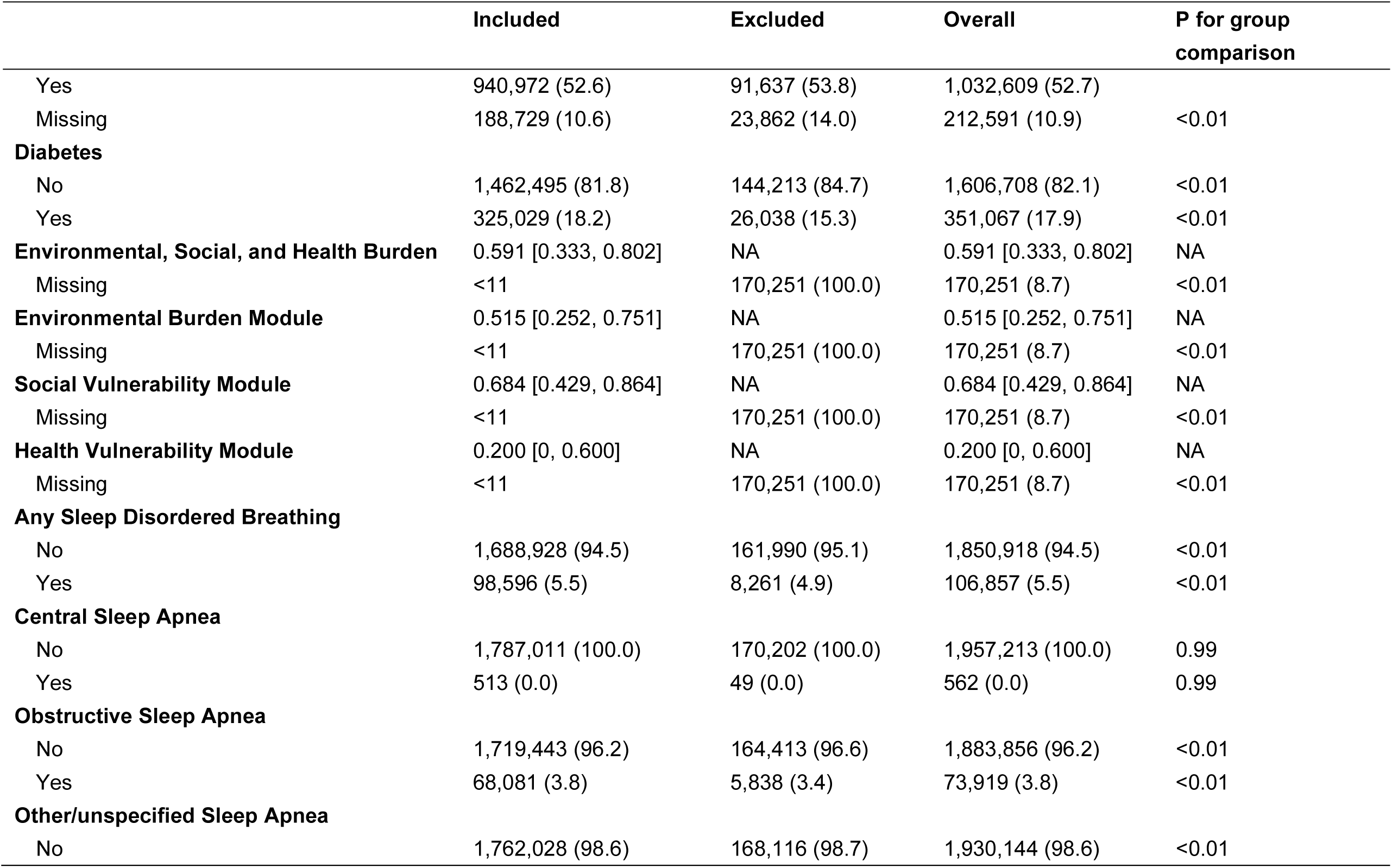
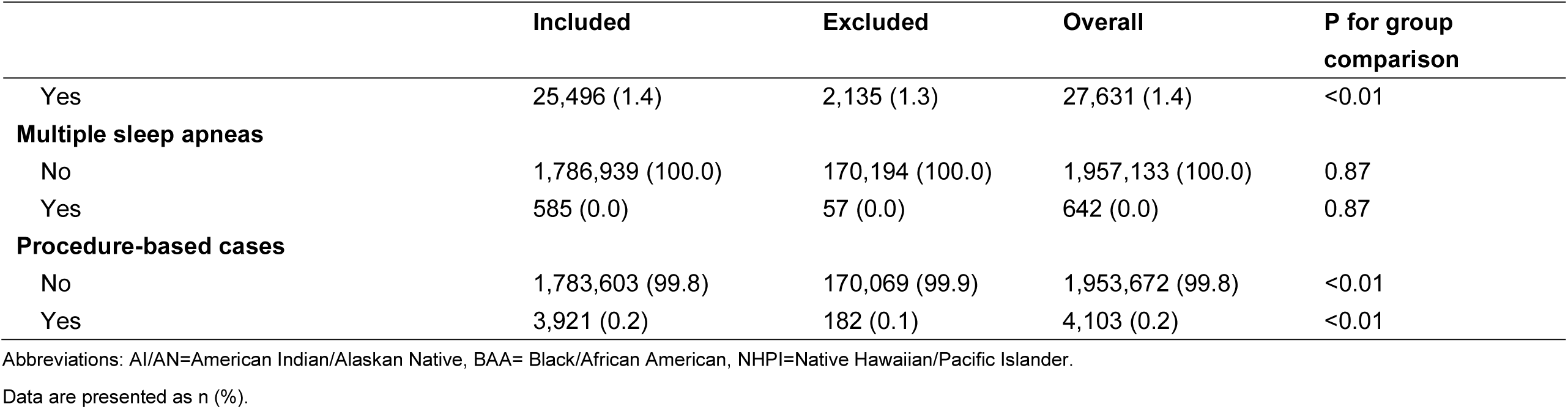
Comparison of socioeconomic characteristics between the included and excluded samples in the study (n=1,957,775)

## Supplemental Text

### METHODS

#### Data sources

Patient data were obtained from an electronic health record (EHR) data from the Across a National Community Health Center Network (ADVANCE), led by OCHIN (https://www.pcori.org/research-results/2015/accelerating-data-value-across-national-community-health-center-network-advance). ^1^ ADVANCE includes more than 144 independent health systems and over five million patients seen at safety-net and Federally Qualified Health Center (FQHC) clinics, making it a nationally representative Clinical Data Research Network (CDRN). ^1^ The OCHIN is a national nonprofit health technology organization that includes more than 34,500 providers at over 2,000 community-based health centers across 40 U.S. states. Among patients included in OCHIN, 55% were at or below the federal poverty level, 80% were uninsured or relied on public health insurance, and 16.5% resided in rural communities. ^2^ These patients are publicly insured or underinsured and have historically been underrepresented in research due to limited access to medical resources stemming from poverty and a general distrust of health care systems. ^1^ The population’s relatively low income and racial and ethnic diversity provide a meaningful representation of individuals experiencing health disparities. ^3,4^

#### Exposure assessment: Environmental, Social, and Health Burden (ESHB)

The U.S. Department of Health and Human Services’ Office of Environmental Justice and the Centers for Disease Control and Prevention (CDC) and Agency for Toxic Substances and Disease Registry (ATSDR) developed a metric at the census tract level that integrates environmental, social, and health indicators. ^5^ These indicators are from various sources, including multiple federal agencies, such as the U.S. Census Bureau, the U.S. Environmental Protection Agency, the U.S. Mine Safety and Health Administration, and the CDC. The ESHB ranks census tracts on 10 different domains (racial/ethnic minority status, socioeconomic status, household characteristics, housing type, air pollution, potentially hazardous and toxic sites, built environment, transportation infrastructure, water pollution, pre-existing chronic disease burden) categorized under three main burdens/vulnerabilities, which represent environmental burden (i.e., 17 indicators reflecting air pollution, potentially hazardous and toxic sites, built environment, transportation infrastructure, and water pollution), social vulnerability (i.e., 14 indicators reflecting racial/ethnic minority status, socioeconomic status, household characteristics, and housing type), and health vulnerability (i.e., five indicators reflecting pre-existing chronic disease burden). ^5^ Accordingly, there are three submodules: the Environmental Burden Module [EBM], the Social Vulnerability Module [SVM], and the Health Vulnerability Module [HVM]. An overall ESHB score was computed by summing the ranked scores of three modules. The sum of the three module rankings (range: 0-3) was converted to a percentile rank (range: 0-1) across all U.S. census tracts, where higher values indicate higher vulnerability.

#### Potential Confounders

Sociodemographic characteristics of patients, including age category in 2022 (categorized as 18-34, 35-49, 50 years and older), sex (man, woman), race (American Indian/Alaskan Native [AI/AN], Asian, Black/African American [BAA], Native Hawaiian/Pacific Islander [NHPI], White, multiple, and unknown), and ethnicity (Hispanic, Non-Hispanic, unknown). Health behavior and clinical characteristics including body mass index (BMI; diagnosed at the closest visit prior to or during 2022), smoking status (the closest non-missing data to first encounter in 2022), alcohol and substance use status (ever reported or diagnosed prior to or during 2022), physical activity (measured at the closest visit prior to or during 2022), and comorbidities were obtained. Obesity (BMI ≥30 kg/m^2^) was identified using ICD-9/10-CM codes (E66.9). Smoking status was a self-reported variable using data from the general information of patients. Alcohol and substance use disorders were identified using ICD-9/10 codes, a Drug Abuse Screening Test (DAST-10) score ≥3, or an Alcohol Use Disorders Identification Test - Consumption (AUDIT-C) score ≥3 for women or ≥4 for men. ^6,7^ Patients meeting any of these criteria were classified as positive (“yes”) for alcohol or substance abuse, while alcohol or substance abuse was defined as “no” among patients with no indicated ICD codes and who met criteria as ‘negative’ for all tests (DAST-10 <3, AUDIT-C <3 for women or <4 for men). Patient-reported physical activity status was categorized into three groups based on the Physical Activity Guidelines for adults and older adults, which recommend at least 150 minutes to 300 minutes a week of moderate-intensity, or 75 minutes to 150 minutes a week of vigorous-intensity aerobic physical activity, or an equivalent combination of moderate- and vigorous-intensity aerobic activity. ^8^ The cut points were used because intensity of physical activity was not captured in the EHR. Thus, the three categories were: physically inactive (<10 minutes), insufficiently active (10-150 minutes/week), and physically active (weekly physical activity >150 minutes/week). Comorbidities included hypertension (at first encounter, including prior to 2022) and diabetes (ever diagnosed prior to or during 2022). Hypertension was defined as ≥1 Systolic BP reading ≥130 mmHg, diastolic BP reading ≥80 mmHg, hypertension medication, or ICD9/10 diagnosis. Types 1 and 2 diabetes were defined as ever diagnosed with diabetes (by the time of the 2022 assessment) and identified using the Centers for Medicare and Medicaid Services Chronic Conditions Data Warehouse (CMS CCDW) codes. ^9^

#### Sensitivity analysis

As a sensitivity analysis, we examined Bonferroni-corrected p-values and confidence intervals to adjust for multiple comparisons. ^10^ Bonferroni multiple testing corrections for the stratified models are reported in the supplemental materials. We further excluded patients with a history of alcohol or substance use, as studies suggest a positive association between substance abuse and sleep disorders. ^11,12^ This analysis helped ensure the robustness of our findings by confirming whether excluding these individuals would alter the main results.

### RESULTS

#### Patient characteristics

For ESHB submodules, EBM ranks were lower in patients with SDB (including all SDB subtypes); however, SVM was highest among procedure-based cases, and HVM was consistently higher among patients with SDB (including its subtypes).

#### Environmental, social, and health burden and prevalence of SDB

Each 0.1-unit increase in EBM percentile rank was associated with a lower prevalence of any SDB (PR=0.97 [0.97–0.97]), OSA (PR=0.97 [0.97–0.97]), and OUSA (PR=0.99 [0.99–0.99]), multiple sleep apneas (PR=0.89 [0.86–0.91]), and procedure-based cases (PR=0.94 [0.93–0.95]). Each 0.1-unit increase in SVM percentile rank was associated with a higher prevalence of any SDB (PR=1.02 [1.01–1.02]), OSA (PR=1.02 [1.01–1.02]), multiple sleep apneas (PR=1.06 [1.02–1.09]), and procedure-based cases (PR=1.08 [1.07–1.10]). Each 0.1-unit increase in HVM percentile rank was associated with a higher documented prevalence of any SDB (PR=1.02 [1.02–1.02]), OSA (PR=1.02 [1.02–1.02]), OUSA (PR=1.02 [1.02–1.03]), multiple sleep apneas (PR=1.04 [1.01–1.06]), and procedure-based cases (PR=1.04 [1.03–1.05]). There were no associations with CSA.

#### Effect modification of associations between EBM, SVM, and HVM in relation to SDB

##### By age

EBM was more strongly associated with a lower prevalence of any SDB and OSA among the oldest age group (PR_18-34 years=_0.98 [0.97–0.99], PR_35-49 years_=0.97 [0.97–0.98], PR≥_50 years_=0.97 [0.97–0.97], P_interaction_<0.01; eTable 5). SVM and HVM were more strongly associated with any SDB and specific subtypes (i.e., OSA and OUSA) among the youngest age group (e.g., SVM–SDB: PR_18-34 years_=1.03 [1.02–1.04], PR_35-49 years_=1.02 [1.02–1.03], PR_≥50 years_=1.01 [1.01–1.02]; HVM-SDB: PR_18-34 years_=1.04 [1.03–1.04], PR_35-49 years_=1.03 [1.03–1.04], PR_≥50 years_=1.02 [1.01–1.02]; eTable 6-7).

##### By sex

There were stronger protective associations between EBM and SDB, OSA, and OUSA among men (e.g., SDB: PR_men_=0.96 [0.96–0.97], PR_women_=0.98 [0.98–0.98]; eTable 5). Consistent with ESHB, SVM and HVM were more strongly related to SDB, OSA, and OUSA in women than men (eTable 6-7).

##### By race

EBM had stronger protective associations with SDB and OSA among AI/AN individuals (SDB: PR_AI/AN_=0.94 [0.93–0.96] and OSA: PR_AI/AN_=0.93 [0.91–0.95]) and with multiple sleep apneas among multiracial individuals (PR_Multiracial_=0.72 [0.55–0.94]) than White patients (SDB: PR_White_=0.97 [0.97–0.97], OSA: PR_White_=0.96 [0.96–0.97], Multiple: PR_White_=0.94 [0.92–0.96]). Among BAA, EBM was also associated with a lower prevalence of procedure-based cases (PR=0.86 [0.83–0.88]; eTable 5). Overall, the patterns of association for SVM and HVM with SDB or OSA were consistent with those observed for the ESHB-SDB association (eTable 6-7), with one subtle difference: for the SVM and SDB relationship, PRs were comparable between AI/AN and White participants but lower among Asian participants (SDB: PR_Asian_=0.99 [0.98–1.01] and OSA: PR_Asian_=0.98 [0.96–0.99]; eTable 6). For procedure-based case prevalences, the associations with SVM and HVM were higher among Asian compared to White patients (SVM: PR_Asian_=1.30 [1.23–1.37] and PR_White_=1.09 [1.07–1.11]; HVM: PR_Asian_=1.29 [1.23–1.34] and PR_White_=1.04 [1.02–1.05]; eTable 6-7), similar to observed associations with ESHB (eTable 5).

##### By ethnicity

There was an inverse association between ESHB and SDB (PR_Hispanic_=0.98 [0.98–0.99]), OSA (PR_Hispanic_=0.99 [0.98–0.99]), and OUSA (PR_Hispanic_=0.98 [0.97–0.99]) among Hispanic patients (eTable 4); EBM conferred a protective association against SDB and procedure-based cases in non-Hispanic patients, but the CIs overlapped with those observed among Hispanic patients (eTable 5). There were stronger associations between higher EBM and lower prevalence of OSA and OUSA among Hispanic patients (eTable 5). For SVM, there were inverse associations with SDB, OSA, OUSA, and procedure-based cases among Hispanic patients, whereas higher SVM was linked to higher prevalence of these outcomes among non-Hispanic patients (eTable 6). For HVM, more pronounced positive associations with SDB, OSA, OUSA, and procedure-based cases were observed in non-Hispanic individuals (eTable 7).

---

**eTable 2.** Detailed identification of sleep-disordered breathing (SDB) and its type.

| Variable | Variable specifications |
| --- | --- |
| SDB | Identified using (1) diagnosis codes at a study encounter, (2) procedural codes, (3) surgical codes, and (4) diagnosis codes from the medical history and conditions tables (named Problem list in the OCHIN data). |
|  | a. **Non-SDB case:** Patients without an SDB ICD 9/10 code (eTable 3) AND no CPAP/BiPAP procedure/surgical codes or Mandibular Advancement Device codes (E0485 and E0486; eTable 4) AND sleep apnea not indicated on the problem list (eTable 4).<br>b. **SDB case:** SDB ICD 9/10 code (eTable 3) OR CPAP/BiPAP procedure/surgical codes and Mandibular Advancement Device codes (E0485 and E0486; eTable 3) OR sleep apnea indicated as active on the problem list (eTable 4). |
| SDB Type<br>(only among<br>SDB cases) | c. **Central sleep apnea:** central sleep apnea ICD 9/10 codes (eTable 3) AND without the presence of obstructive sleep apnea diagnosis code<br>d. **Obstructive sleep apnea:** obstructive sleep apnea ICD 9/10 codes (eTable 3) OR Mandibular Advancement Device codes (E0485 and E0486; eTable 3) AND without the presence of a central sleep apnea diagnosis code<br>e. **Procedure-based cases** (*cases identified from surgical and procedure codes*): Patients identified as an SDB case with HCPCS/CPT Codes from surgical history or procedures tables for CPAP and BiPAP Machine and Accessories or Surgical Procedure Implants (eTable 3) and NOT Mandibular Advancement Device codes (eTable 3), and no central or obstructive sleep apnea diagnosis identified with ICD 9/10 codes<br>*\*\* Note: these patients have missing information as a construct of the EHR; this is not necessarily a distinct diagnosis group. Suggest labelling as cases identified from surgical and procedure codes, or cases missing diagnosis information, to help readers better understand the patients this group is capturing*<br>f. **Unspecified** (*suggest using “unspecified” as this group name, as “unspecified” is used for ICD codes included in this group (e.g., codes: G47.30, G47.3, 780.57)*): Patients with an unspecified SDB ICD 9/10 diagnosis code (eTable 3) only and no CSA or OSA ICD 9/10 code.<br>g. More than one SDB type identified (**multiple**): cases with OSA and CSA ICD 9/10 codes, or CSA ICD 9/10 and MAD HCPCS/CPT Codes. See eTable 3-4 below. |

**eTable 3.**
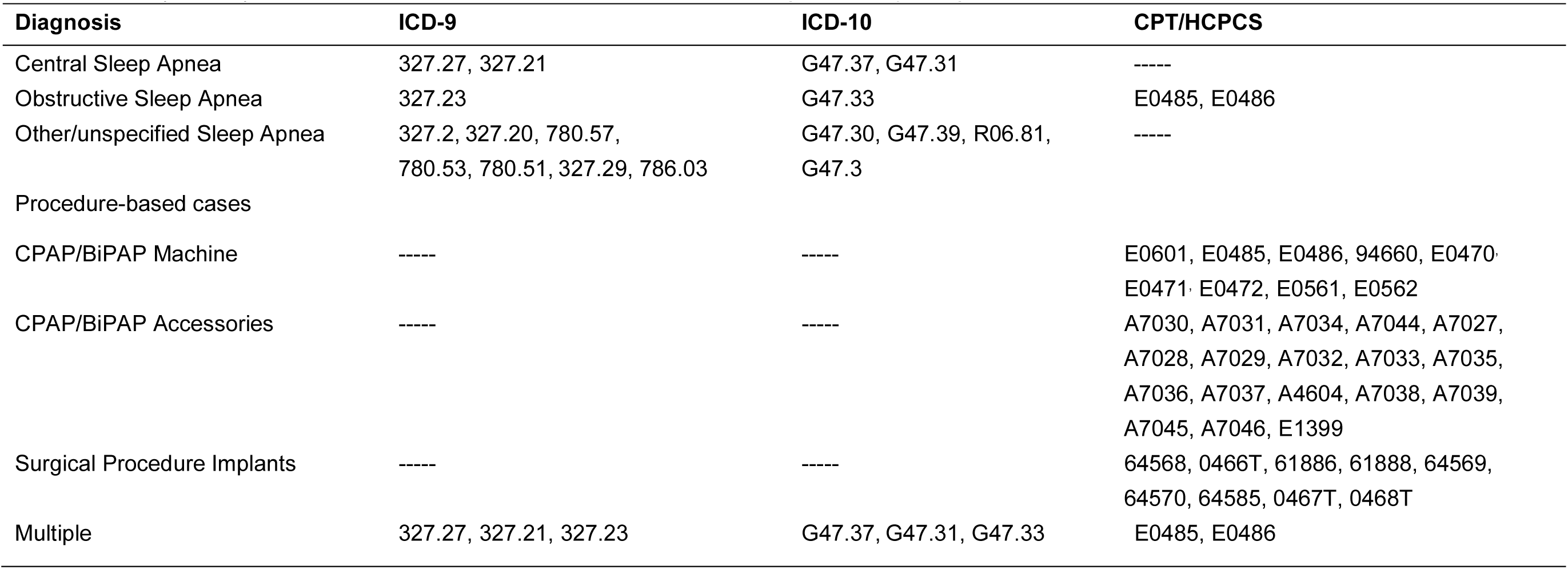
ICD-9/ICD-10 Codes and CPT/HCPCS for central sleep apnea (CSA), obstructive sleep apnea (OSA), and other/unspecified sleep apnea (OUSA), multiple sleep apneas, and sleep apnea diagnosed by surgical/procedural code

**eTable 4.**
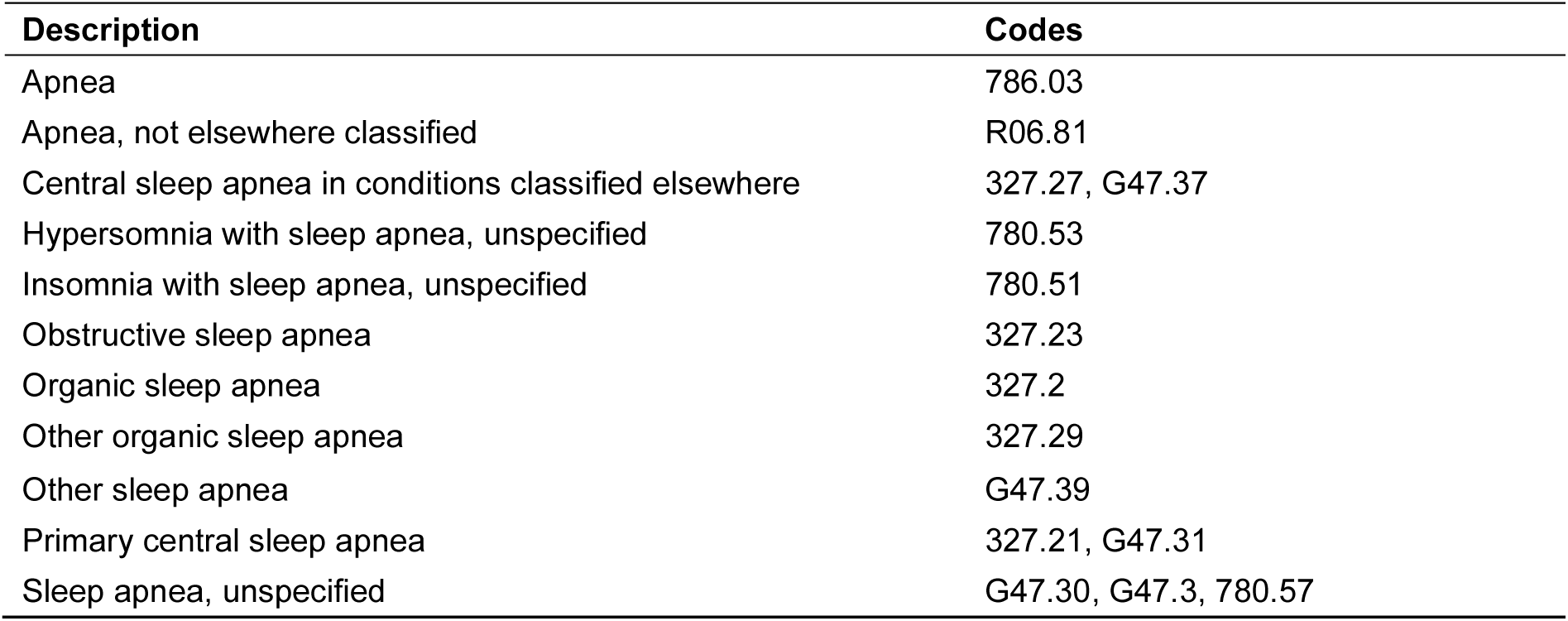
Sleep apnea conditions captured in the problem list

**eTable 5.**
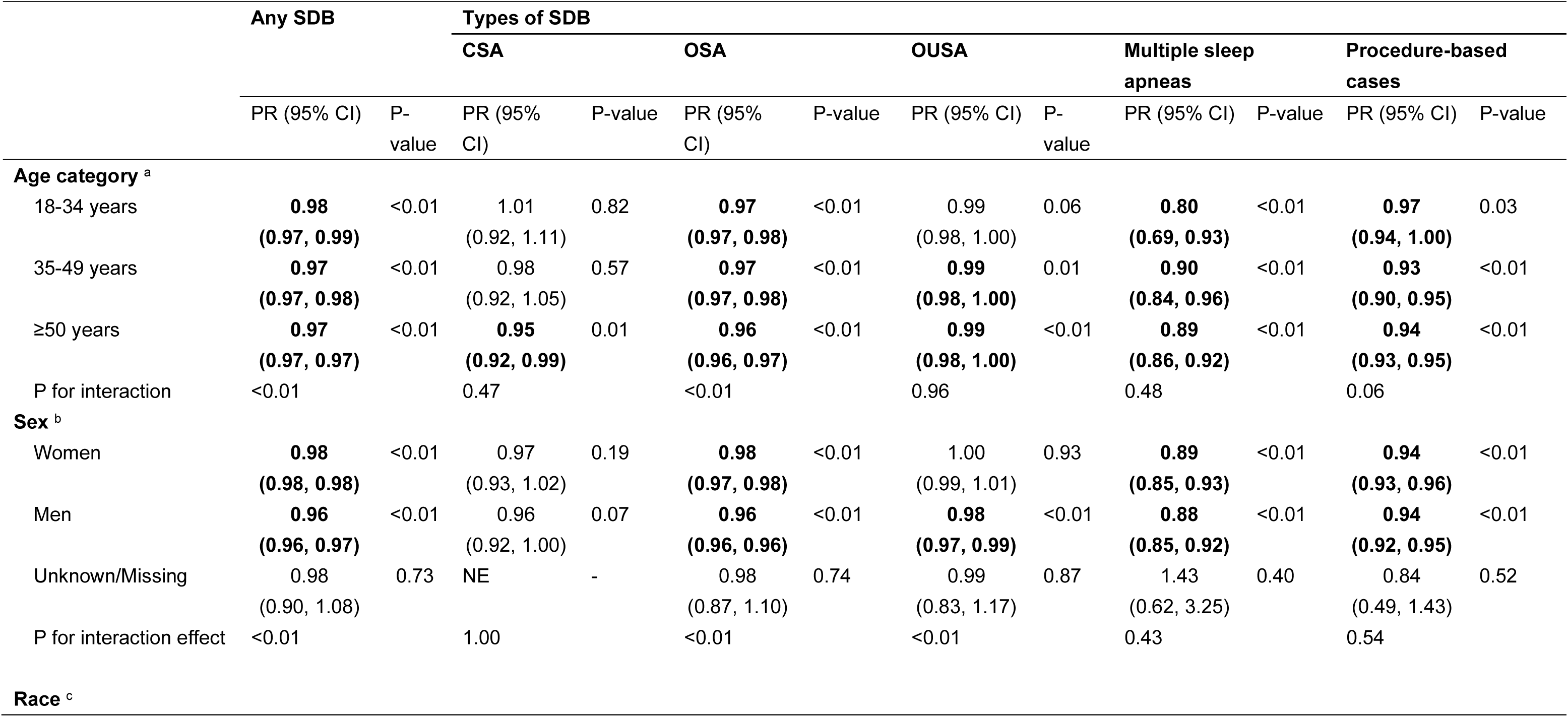
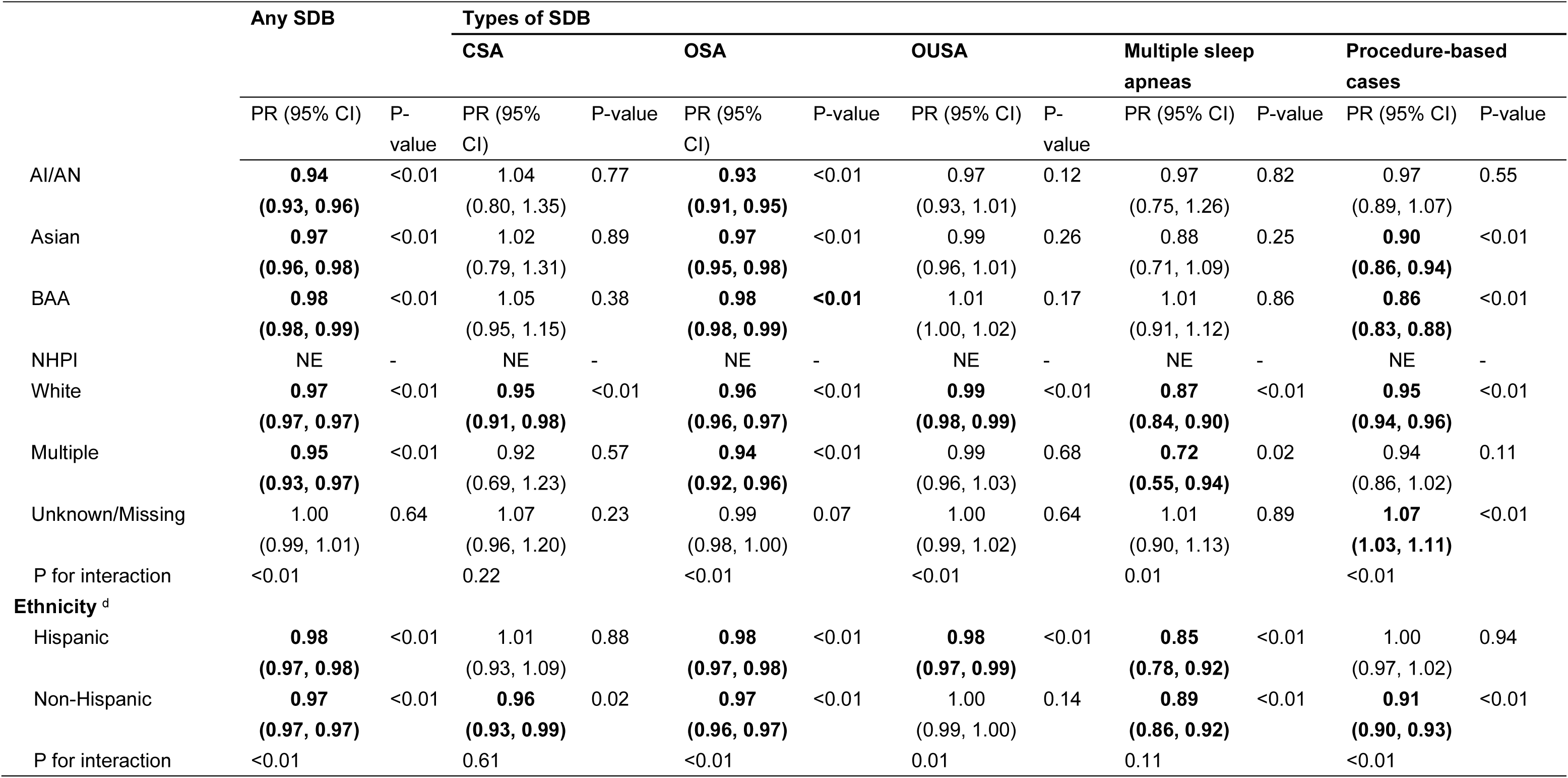
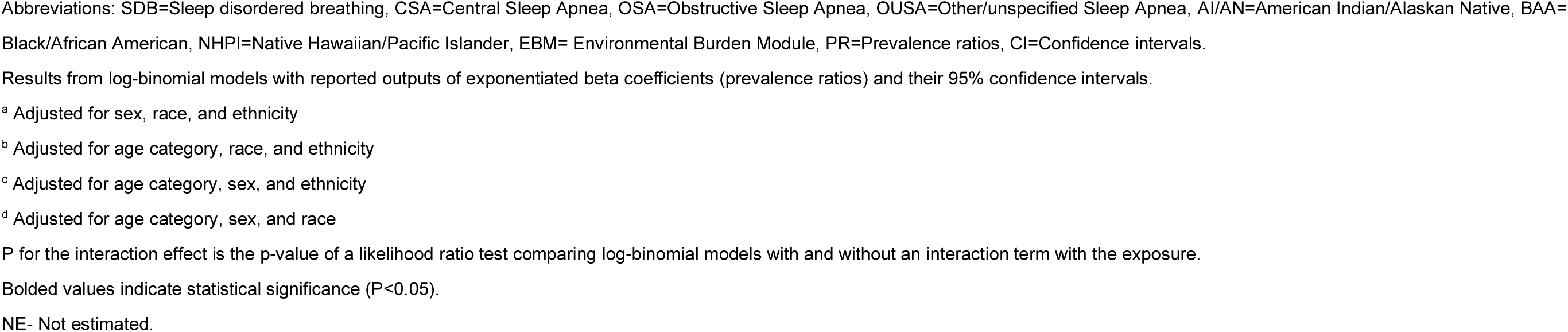
Subgroup cross-sectional associations of per 0.1-unit increase in Environmental Burden Module (EBM) ranks with sleep-disordered breathing (SDB), central sleep apnea (CSA), obstructive sleep apnea (OSA), and other/unspecified sleep apnea (OUSA), multiple sleep apneas, and procedure-based cases

|  | Any SDB |  | Types of SDB |  |  |  |  |  |  |  |  |  |
| --- | --- | --- | --- | --- | --- | --- | --- | --- | --- | --- | --- | --- |
|  |  |  | CSA |  | OSA |  | OUSA |  | Multiple sleep apneas |  | Procedure-based cases |  |
|  | PR (95% CI) | P-value | PR (95% CI) | P-value | PR (95% CI) | P-value | PR (95% CI) | P-value | PR (95% CI) | P-value | PR (95% CI) | P-value |
| **Age category <sup>a</sup>** |  |  |  |  |  |  |  |  |  |  |  |  |
| 18-34 years | **0.98**<br>**(0.97, 0.99)** | <0.01 | 1.01<br>(0.92, 1.11) | 0.82 | **0.97**<br>**(0.97, 0.98)** | <0.01 | 0.99<br>(0.98, 1.00) | 0.06 | **0.80**<br>**(0.69, 0.93)** | <0.01 | **0.97**<br>**(0.94, 1.00)** | 0.03 |
| 35-49 years | **0.97**<br>**(0.97, 0.98)** | <0.01 | 0.98<br>(0.92, 1.05) | 0.57 | **0.97**<br>**(0.97, 0.98)** | <0.01 | **0.99**<br>**(0.98, 1.00)** | 0.01 | **0.90**<br>**(0.84, 0.96)** | <0.01 | **0.93**<br>**(0.90, 0.95)** | <0.01 |
| ≥50 years | **0.97**<br>**(0.97, 0.97)** | <0.01 | **0.95**<br>**(0.92, 0.99)** | 0.01 | **0.96**<br>**(0.96, 0.97)** | <0.01 | **0.99**<br>**(0.98, 1.00)** | <0.01 | **0.89**<br>**(0.86, 0.92)** | <0.01 | **0.94**<br>**(0.93, 0.95)** | <0.01 |
| P for interaction | <0.01 |  | 0.47 |  | <0.01 |  | 0.96 |  | 0.48 |  | 0.06 |  |
| **Sex <sup>b</sup>** |  |  |  |  |  |  |  |  |  |  |  |  |
| Women | **0.98**<br>**(0.98, 0.98)** | <0.01 | 0.97<br>(0.93, 1.02) | 0.19 | **0.98**<br>**(0.97, 0.98)** | <0.01 | 1.00<br>(0.99, 1.01) | 0.93 | **0.89**<br>**(0.85, 0.93)** | <0.01 | **0.94**<br>**(0.93, 0.96)** | <0.01 |
| Men | **0.96**<br>**(0.96, 0.97)** | <0.01 | 0.96<br>(0.92, 1.00) | 0.07 | **0.96**<br>**(0.96, 0.96)** | <0.01 | **0.98**<br>**(0.97, 0.99)** | <0.01 | **0.88**<br>**(0.85, 0.92)** | <0.01 | **0.94**<br>**(0.92, 0.95)** | <0.01 |
| Unknown/Missing | 0.98<br>(0.90, 1.08) | 0.73 | NE | - | 0.98<br>(0.87, 1.10) | 0.74 | 0.99<br>(0.83, 1.17) | 0.87 | 1.43<br>(0.62, 3.25) | 0.40 | 0.84<br>(0.49, 1.43) | 0.52 |
| P for interaction effect | <0.01 |  | 1.00 |  | <0.01 |  | <0.01 |  | 0.43 |  | 0.54 |  |
| **Race <sup>c</sup>** |  |  |  |  |  |  |  |  |  |  |  |  |
|  |  |  | CSA |  | OSA |  | OUSA |  | Multiple sleep apneas |  | Procedure-based cases |  |
|  | PR (95% CI) | P-value | PR (95% CI) | P-value | PR (95% CI) | P-value | PR (95% CI) | P-value | PR (95% CI) | P-value | PR (95% CI) | P-value |
| AI/AN | **0.94**<br>**(0.93, 0.96)** | <0.01 | 1.04<br>(0.80, 1.35) | 0.77 | **0.93**<br>**(0.91, 0.95)** | <0.01 | 0.97<br>(0.93, 1.01) | 0.12 | 0.97<br>(0.75, 1.26) | 0.82 | 0.97<br>(0.89, 1.07) | 0.55 |
| Asian | **0.97**<br>**(0.96, 0.98)** | <0.01 | 1.02<br>(0.79, 1.31) | 0.89 | **0.97**<br>**(0.95, 0.98)** | <0.01 | 0.99<br>(0.96, 1.01) | 0.26 | 0.88<br>(0.71, 1.09) | 0.25 | **0.90**<br>**(0.86, 0.94)** | <0.01 |
| BAA | **0.98**<br>**(0.98, 0.99)** | <0.01 | 1.05<br>(0.95, 1.15) | 0.38 | **0.98**<br>**(0.98, 0.99)** | <0.01 | 1.01<br>(1.00, 1.02) | 0.17 | 1.01<br>(0.91, 1.12) | 0.86 | **0.86**<br>**(0.83, 0.88)** | <0.01 |
| NHPI | NE | - | NE | - | NE | - | NE | - | NE | - | NE | - |
| White | **0.97**<br>**(0.97, 0.97)** | <0.01 | **0.95**<br>**(0.91, 0.98)** | <0.01 | **0.96**<br>**(0.96, 0.97)** | <0.01 | **0.99**<br>**(0.98, 0.99)** | <0.01 | **0.87**<br>**(0.84, 0.90)** | <0.01 | **0.95**<br>**(0.94, 0.96)** | <0.01 |
| Multiple | **0.95**<br>**(0.93, 0.97)** | <0.01 | 0.92<br>(0.69, 1.23) | 0.57 | **0.94**<br>**(0.92, 0.96)** | <0.01 | 0.99<br>(0.96, 1.03) | 0.68 | **0.72**<br>**(0.55, 0.94)** | 0.02 | 0.94<br>(0.86, 1.02) | 0.11 |
| Unknown/Missing | 1.00<br>(0.99, 1.01) | 0.64 | 1.07<br>(0.96, 1.20) | 0.23 | 0.99<br>(0.98, 1.00) | 0.07 | 1.00<br>(0.99, 1.02) | 0.64 | 1.01<br>(0.90, 1.13) | 0.89 | **1.07**<br>**(1.03, 1.11)** | <0.01 |
| P for interaction | <0.01 |  | 0.22 |  | <0.01 |  | <0.01 |  | 0.01 |  | <0.01 |  |
| **Ethnicity<sup>d</sup>** |  |  |  |  |  |  |  |  |  |  |  |  |
| Hispanic | **0.98**<br>**(0.97, 0.98)** | <0.01 | 1.01<br>(0.93, 1.09) | 0.88 | **0.98**<br>**(0.97, 0.98)** | <0.01 | **0.98**<br>**(0.97, 0.99)** | <0.01 | **0.85**<br>**(0.78, 0.92)** | <0.01 | 1.00<br>(0.97, 1.02) | 0.94 |
| Non-Hispanic | **0.97**<br>**(0.97, 0.97)** | <0.01 | **0.96**<br>**(0.93, 0.99)** | 0.02 | **0.97**<br>**(0.96, 0.97)** | <0.01 | 1.00<br>(0.99, 1.00) | 0.14 | **0.89**<br>**(0.86, 0.92)** | <0.01 | **0.91**<br>**(0.90, 0.93)** | <0.01 |
| P for interaction | <0.01 |  | 0.61 |  | <0.01 |  | 0.01 |  | 0.11 |  | <0.01 |  |

Abbreviations: SDB=Sleep disordered breathing, CSA=Central Sleep Apnea, OSA=Obstructive Sleep Apnea, OUSA=Other/unspecified Sleep Apnea, AI/AN=American Indian/Alaskan Native, BAA=Black/African American, NHPI=Native Hawaiian/Pacific Islander, EBM= Environmental Burden Module, PR=Prevalence ratios, CI=Confidence intervals.
Results from log-binomial models with reported outputs of exponentiated beta coefficients (prevalence ratios) and their 95% confidence intervals.
<sup>a</sup> Adjusted for sex, race, and ethnicity
<sup>b</sup> Adjusted for age category, race, and ethnicity
<sup>c</sup> Adjusted for age category, sex, and ethnicity
<sup>d</sup> Adjusted for age category, sex, and race
P for the interaction effect is the p-value of a likelihood ratio test comparing log-binomial models with and without an interaction term with the exposure.
Bolded values indicate statistical significance ( $P < 0.05$ ).
NE- Not estimated.

**eTable 6.**
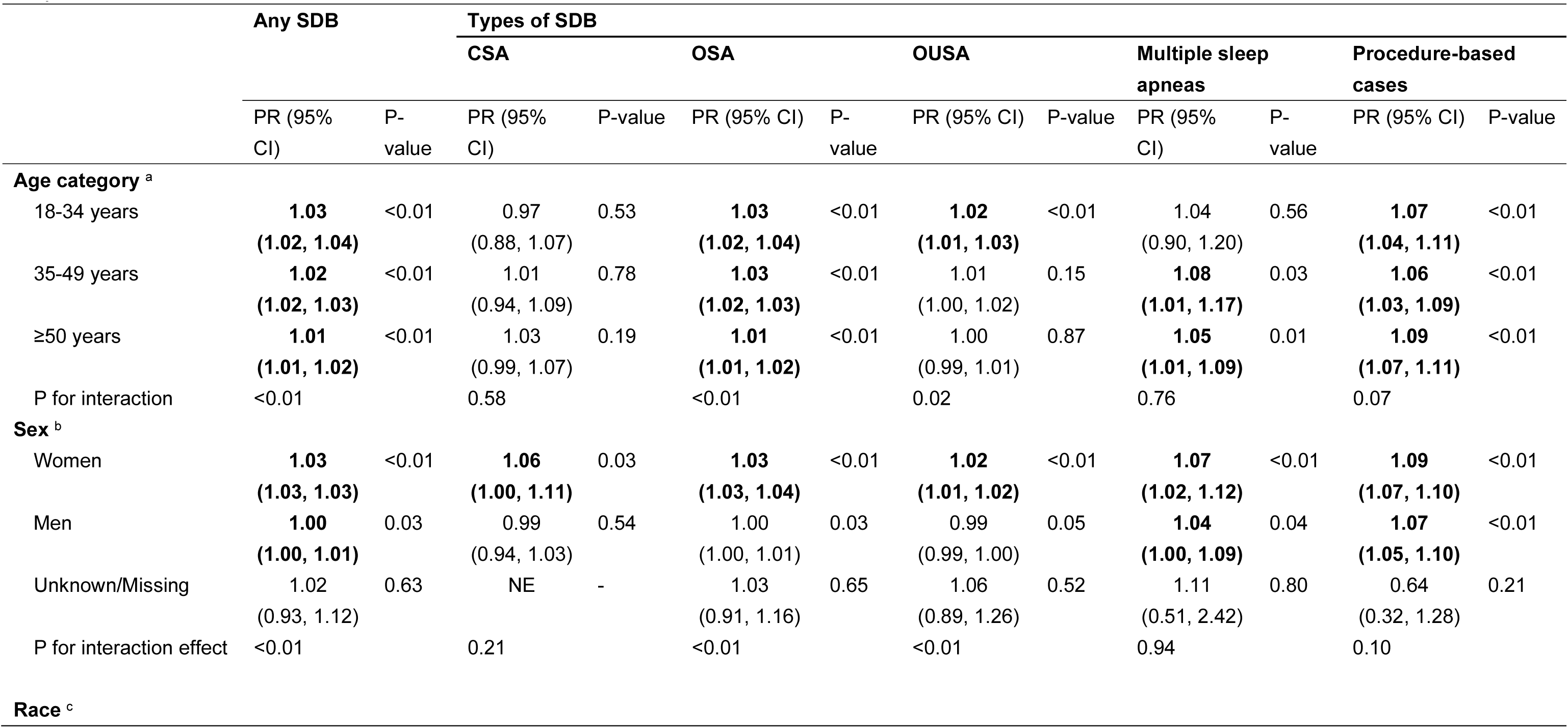
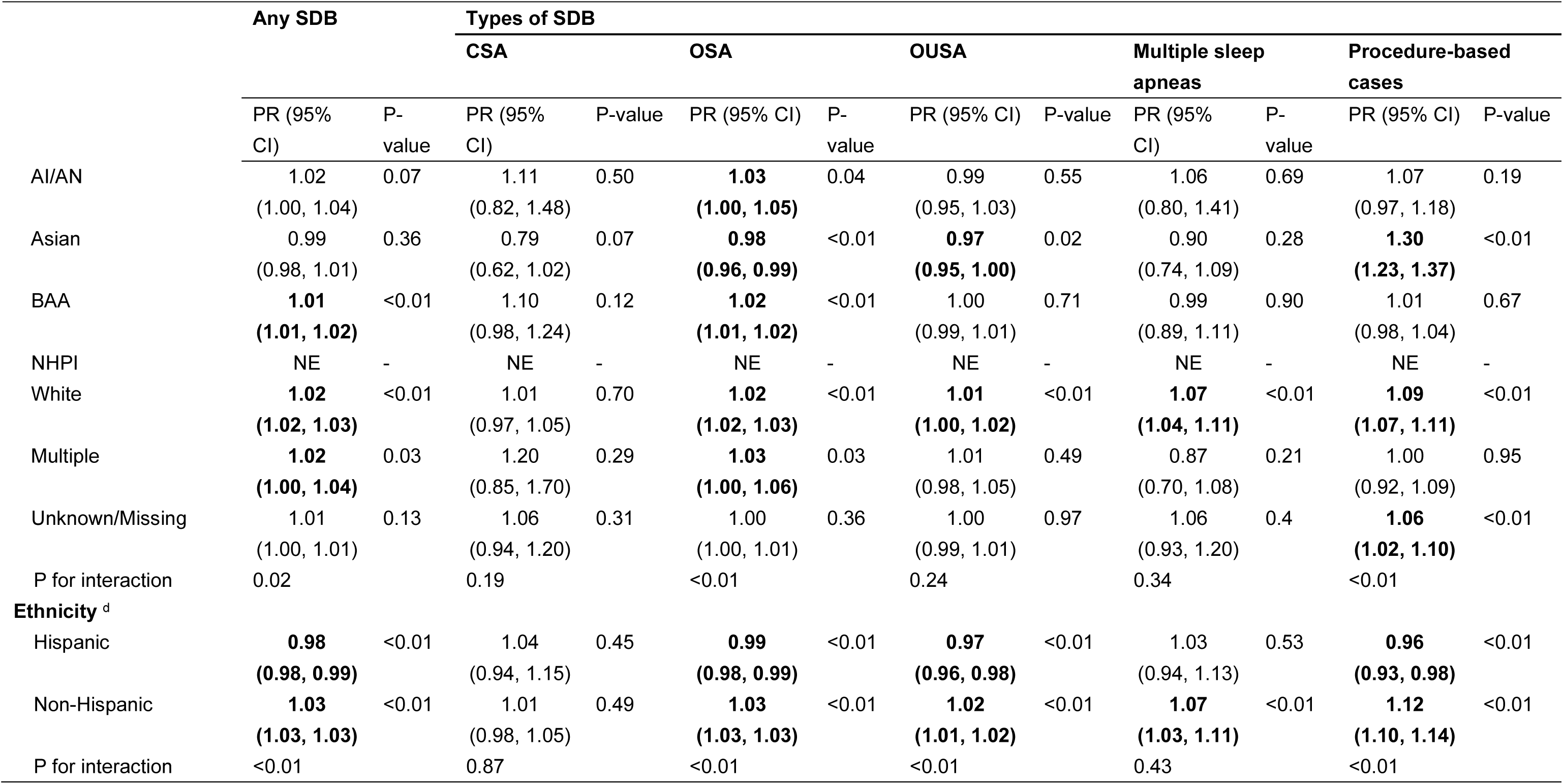
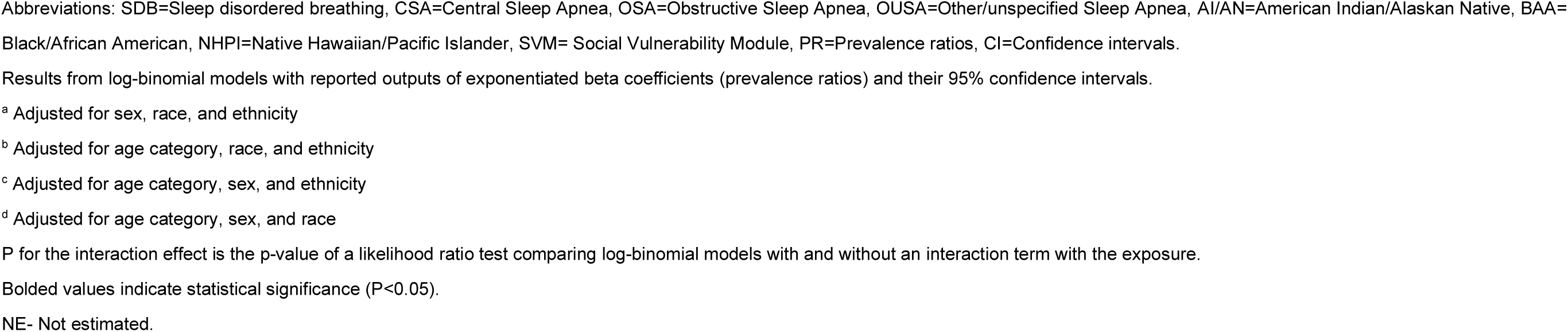
Subgroup cross-sectional associations of per 0.1-unit increase in Social Vulnerability Module (SVM) ranks with sleep-disordered breathing (SDB), central sleep apnea (CSA), obstructive sleep apnea (OSA), and other/unspecified sleep apnea (OUSA), multiple sleep apneas, and procedure-based cases

|  | Any SDB |  | Types of SDB |  |  |  |  |  |  |  |  |  |
| --- | --- | --- | --- | --- | --- | --- | --- | --- | --- | --- | --- | --- |
|  |  |  | CSA |  | OSA |  | OUSA |  | Multiple sleep apneas |  | Procedure-based cases |  |
|  | PR (95% CI) | P-value | PR (95% CI) | P-value | PR (95% CI) | P-value | PR (95% CI) | P-value | PR (95% CI) | P-value | PR (95% CI) | P-value |
| **Age category <sup>a</sup>** |  |  |  |  |  |  |  |  |  |  |  |  |
| 18-34 years | **1.03**<br>**(1.02, 1.04)** | <0.01 | 0.97<br>(0.88, 1.07) | 0.53 | **1.03**<br>**(1.02, 1.04)** | <0.01 | **1.02**<br>**(1.01, 1.03)** | <0.01 | 1.04<br>(0.90, 1.20) | 0.56 | **1.07**<br>**(1.04, 1.11)** | <0.01 |
| 35-49 years | **1.02**<br>**(1.02, 1.03)** | <0.01 | 1.01<br>(0.94, 1.09) | 0.78 | **1.03**<br>**(1.02, 1.03)** | <0.01 | 1.01<br>(1.00, 1.02) | 0.15 | **1.08**<br>**(1.01, 1.17)** | 0.03 | **1.06**<br>**(1.03, 1.09)** | <0.01 |
| ≥50 years | **1.01**<br>**(1.01, 1.02)** | <0.01 | 1.03<br>(0.99, 1.07) | 0.19 | **1.01**<br>**(1.01, 1.02)** | <0.01 | 1.00<br>(0.99, 1.01) | 0.87 | **1.05**<br>**(1.01, 1.09)** | 0.01 | **1.09**<br>**(1.07, 1.11)** | <0.01 |
| P for interaction | <0.01 |  | 0.58 |  | <0.01 |  | 0.02 |  | 0.76 |  | 0.07 |  |
| **Sex <sup>b</sup>** |  |  |  |  |  |  |  |  |  |  |  |  |
| Women | **1.03**<br>**(1.03, 1.03)** | <0.01 | **1.06**<br>**(1.00, 1.11)** | 0.03 | **1.03**<br>**(1.03, 1.04)** | <0.01 | **1.02**<br>**(1.01, 1.02)** | <0.01 | **1.07**<br>**(1.02, 1.12)** | <0.01 | **1.09**<br>**(1.07, 1.10)** | <0.01 |
| Men | **1.00**<br>**(1.00, 1.01)** | 0.03 | 0.99<br>(0.94, 1.03) | 0.54 | 1.00<br>(1.00, 1.01) | 0.03 | 0.99<br>(0.99, 1.00) | 0.05 | **1.04**<br>**(1.00, 1.09)** | 0.04 | **1.07**<br>**(1.05, 1.10)** | <0.01 |
| Unknown/Missing | 1.02<br>(0.93, 1.12) | 0.63 | NE | - | 1.03<br>(0.91, 1.16) | 0.65 | 1.06<br>(0.89, 1.26) | 0.52 | 1.11<br>(0.51, 2.42) | 0.80 | 0.64<br>(0.32, 1.28) | 0.21 |
| P for interaction effect | <0.01 |  | 0.21 |  | <0.01 |  | <0.01 |  | 0.94 |  | 0.10 |  |
| **Race <sup>c</sup>** |  |  |  |  |  |  |  |  |  |  |  |  |
|  |  |  | CSA |  | OSA |  | OUSA |  | Multiple sleep apneas |  | Procedure-based cases |  |
|  | PR (95% CI) | P-value | PR (95% CI) | P-value | PR (95% CI) | P-value | PR (95% CI) | P-value | PR (95% CI) | P-value | PR (95% CI) | P-value |
| AI/AN | 1.02<br>(1.00, 1.04) | 0.07 | 1.11<br>(0.82, 1.48) | 0.50 | **1.03**<br>**(1.00, 1.05)** | 0.04 | 0.99<br>(0.95, 1.03) | 0.55 | 1.06<br>(0.80, 1.41) | 0.69 | 1.07<br>(0.97, 1.18) | 0.19 |
| Asian | 0.99<br>(0.98, 1.01) | 0.36 | 0.79<br>(0.62, 1.02) | 0.07 | **0.98**<br>**(0.96, 0.99)** | <0.01 | **0.97**<br>**(0.95, 1.00)** | 0.02 | 0.90<br>(0.74, 1.09) | 0.28 | **1.30**<br>**(1.23, 1.37)** | <0.01 |
| BAA | **1.01**<br>**(1.01, 1.02)** | <0.01 | 1.10<br>(0.98, 1.24) | 0.12 | **1.02**<br>**(1.01, 1.02)** | <0.01 | 1.00<br>(0.99, 1.01) | 0.71 | 0.99<br>(0.89, 1.11) | 0.90 | 1.01<br>(0.98, 1.04) | 0.67 |
| NHPI | NE | - | NE | - | NE | - | NE | - | NE | - | NE | - |
| White | **1.02**<br>**(1.02, 1.03)** | <0.01 | 1.01<br>(0.97, 1.05) | 0.70 | **1.02**<br>**(1.02, 1.03)** | <0.01 | **1.01**<br>**(1.00, 1.02)** | <0.01 | **1.07**<br>**(1.04, 1.11)** | <0.01 | **1.09**<br>**(1.07, 1.11)** | <0.01 |
| Multiple | **1.02**<br>**(1.00, 1.04)** | 0.03 | 1.20<br>(0.85, 1.70) | 0.29 | **1.03**<br>**(1.00, 1.06)** | 0.03 | 1.01<br>(0.98, 1.05) | 0.49 | 0.87<br>(0.70, 1.08) | 0.21 | 1.00<br>(0.92, 1.09) | 0.95 |
| Unknown/Missing | 1.01<br>(1.00, 1.01) | 0.13 | 1.06<br>(0.94, 1.20) | 0.31 | 1.00<br>(1.00, 1.01) | 0.36 | 1.00<br>(0.99, 1.01) | 0.97 | 1.06<br>(0.93, 1.20) | 0.4 | **1.06**<br>**(1.02, 1.10)** | <0.01 |
| P for interaction | 0.02 |  | 0.19 |  | <0.01 |  | 0.24 |  | 0.34 |  | <0.01 |  |
| **Ethnicity <sup>d</sup>** |  |  |  |  |  |  |  |  |  |  |  |  |
| Hispanic | **0.98**<br>**(0.98, 0.99)** | <0.01 | 1.04<br>(0.94, 1.15) | 0.45 | **0.99**<br>**(0.98, 0.99)** | <0.01 | **0.97**<br>**(0.96, 0.98)** | <0.01 | 1.03<br>(0.94, 1.13) | 0.53 | **0.96**<br>**(0.93, 0.98)** | <0.01 |
| Non-Hispanic | **1.03**<br>**(1.03, 1.03)** | <0.01 | 1.01<br>(0.98, 1.05) | 0.49 | **1.03**<br>**(1.03, 1.03)** | <0.01 | **1.02**<br>**(1.01, 1.02)** | <0.01 | **1.07**<br>**(1.03, 1.11)** | <0.01 | **1.12**<br>**(1.10, 1.14)** | <0.01 |
| P for interaction | <0.01 |  | 0.87 |  | <0.01 |  | <0.01 |  | 0.43 |  | <0.01 |  |

Abbreviations: SDB=Sleep disordered breathing, CSA=Central Sleep Apnea, OSA=Obstructive Sleep Apnea, OUSA=Other/unspecified Sleep Apnea, AI/AN=American Indian/Alaskan Native, BAA=Black/African American, NHPI=Native Hawaiian/Pacific Islander, SVM= Social Vulnerability Module, PR=Prevalence ratios, CI=Confidence intervals.
Results from log-binomial models with reported outputs of exponentiated beta coefficients (prevalence ratios) and their 95% confidence intervals.
<sup>a</sup> Adjusted for sex, race, and ethnicity
<sup>b</sup> Adjusted for age category, race, and ethnicity
<sup>c</sup> Adjusted for age category, sex, and ethnicity
<sup>d</sup> Adjusted for age category, sex, and race
P for the interaction effect is the p-value of a likelihood ratio test comparing log-binomial models with and without an interaction term with the exposure.
Bolded values indicate statistical significance ( $P < 0.05$ ).
NE- Not estimated.

**eTable 7.**
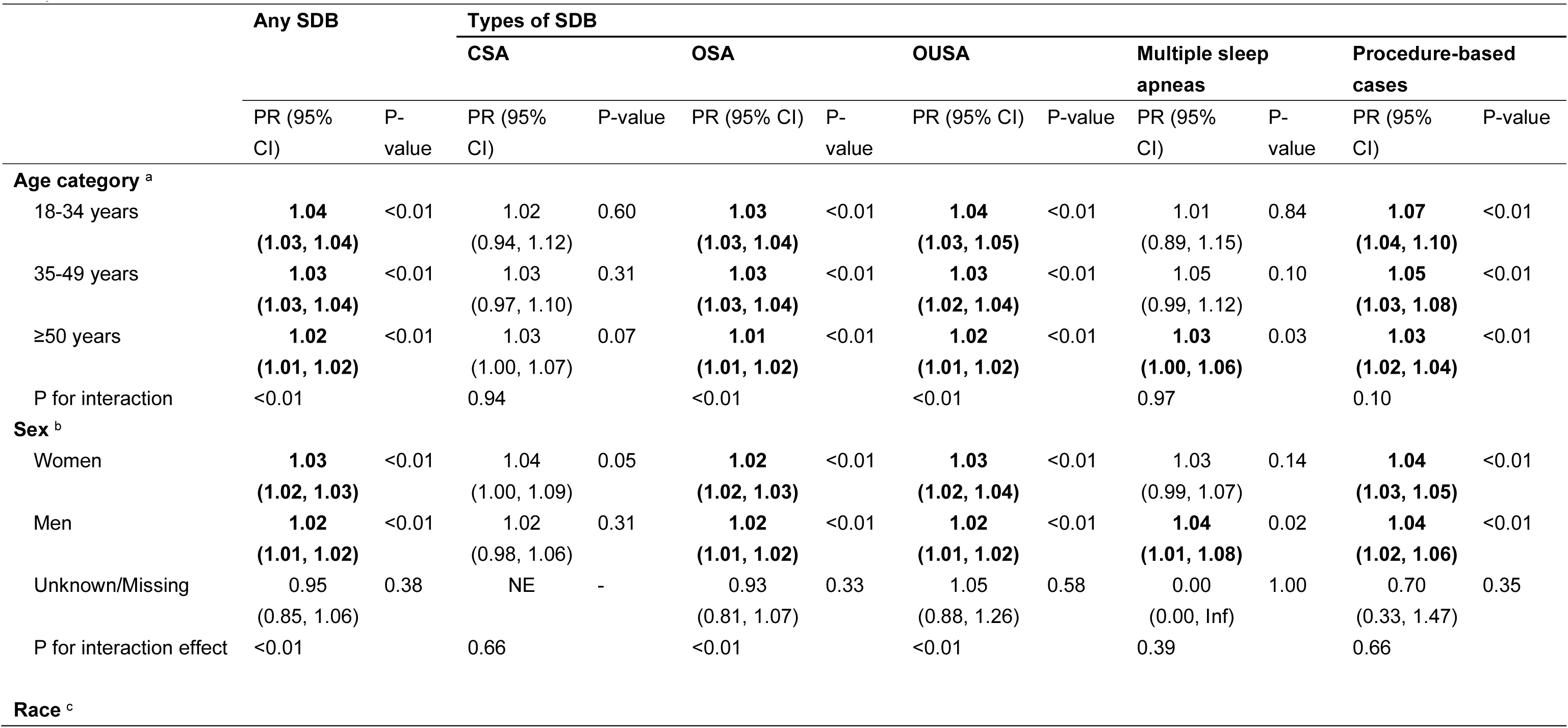
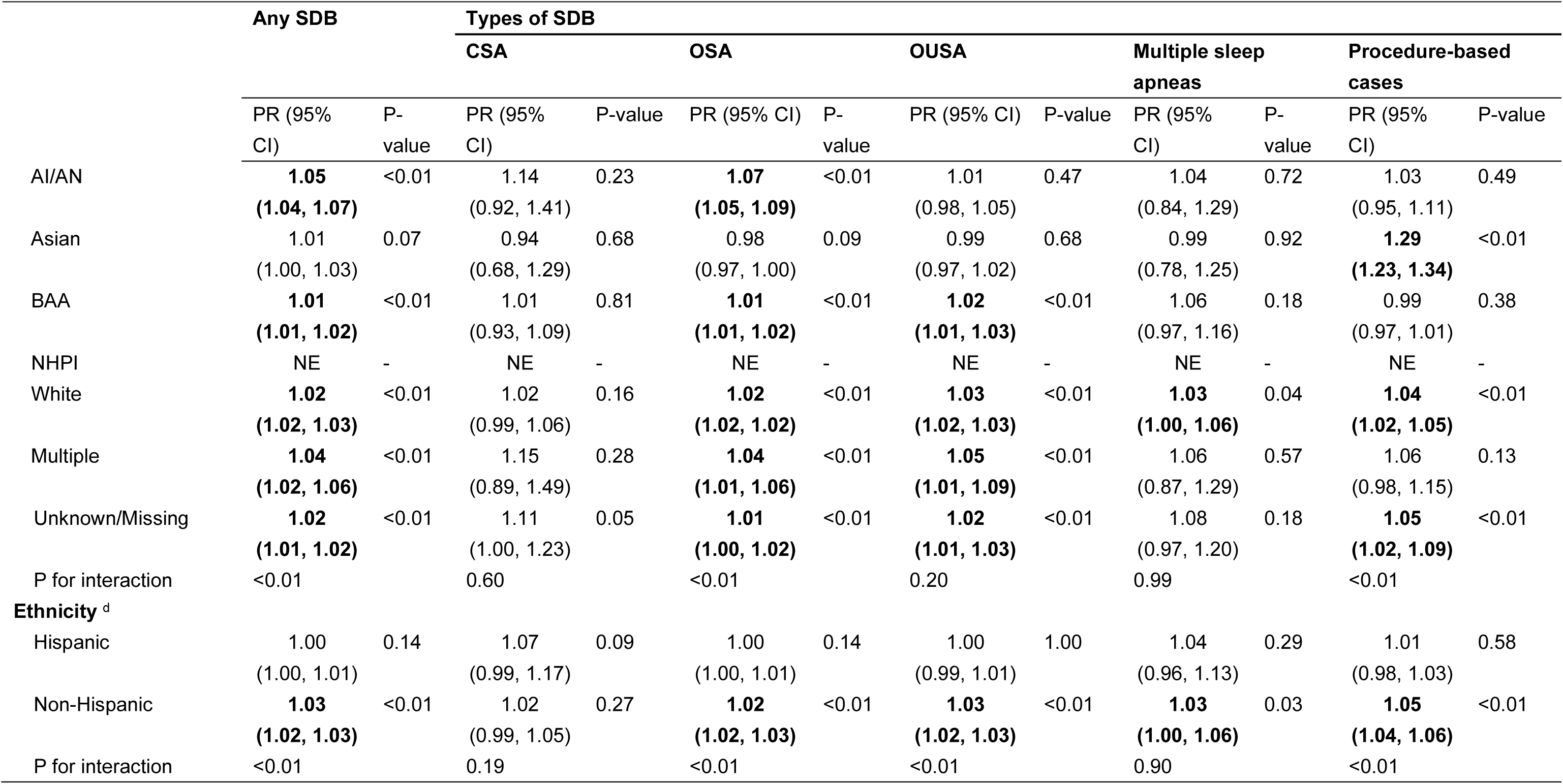
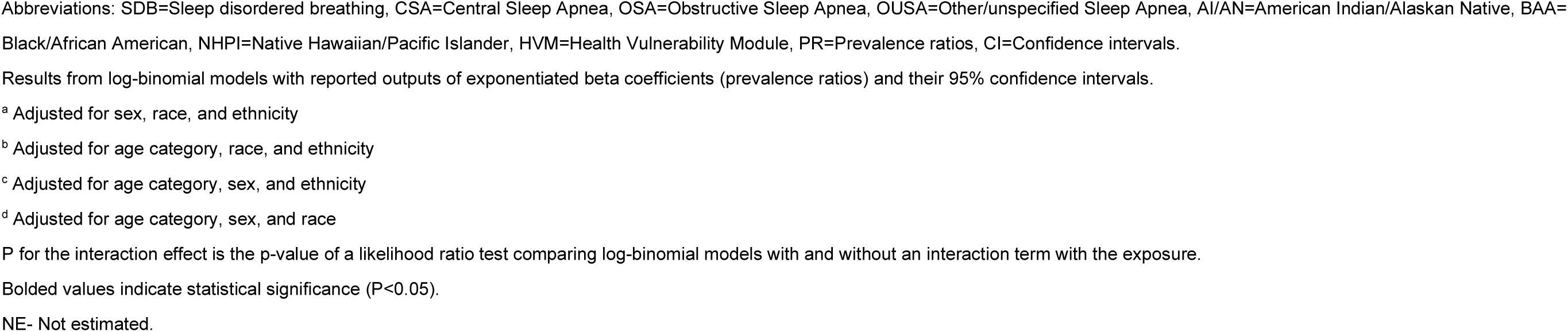
Subgroup cross-sectional associations of per 0.1-unit increase in Health Vulnerability Module (HVM) ranks with sleep-disordered breathing (SDB), central sleep apnea (CSA), obstructive sleep apnea (OSA), and other/unspecified sleep apnea (OUSA), multiple sleep apneas, and procedure-based cases

|  | Any SDB |  | Types of SDB |  |  |  |  |  |  |  |  |  |
| --- | --- | --- | --- | --- | --- | --- | --- | --- | --- | --- | --- | --- |
|  |  |  | CSA |  | OSA |  | OUSA |  | Multiple sleep apneas |  | Procedure-based cases |  |
|  | PR (95% CI) | P-value | PR (95% CI) | P-value | PR (95% CI) | P-value | PR (95% CI) | P-value | PR (95% CI) | P-value | PR (95% CI) | P-value |
| **Age category <sup>a</sup>** |  |  |  |  |  |  |  |  |  |  |  |  |
| 18-34 years | **1.04**<br>**(1.03, 1.04)** | <0.01 | 1.02<br>(0.94, 1.12) | 0.60 | **1.03**<br>**(1.03, 1.04)** | <0.01 | **1.04**<br>**(1.03, 1.05)** | <0.01 | 1.01<br>(0.89, 1.15) | 0.84 | **1.07**<br>**(1.04, 1.10)** | <0.01 |
| 35-49 years | **1.03**<br>**(1.03, 1.04)** | <0.01 | 1.03<br>(0.97, 1.10) | 0.31 | **1.03**<br>**(1.03, 1.04)** | <0.01 | **1.03**<br>**(1.02, 1.04)** | <0.01 | 1.05<br>(0.99, 1.12) | 0.10 | **1.05**<br>**(1.03, 1.08)** | <0.01 |
| ≥50 years | **1.02**<br>**(1.01, 1.02)** | <0.01 | 1.03<br>(1.00, 1.07) | 0.07 | **1.01**<br>**(1.01, 1.02)** | <0.01 | **1.02**<br>**(1.01, 1.02)** | <0.01 | **1.03**<br>**(1.00, 1.06)** | 0.03 | **1.03**<br>**(1.02, 1.04)** | <0.01 |
| P for interaction | <0.01 |  | 0.94 |  | <0.01 |  | <0.01 |  | 0.97 |  | 0.10 |  |
| **Sex <sup>b</sup>** |  |  |  |  |  |  |  |  |  |  |  |  |
| Women | **1.03**<br>**(1.02, 1.03)** | <0.01 | 1.04<br>(1.00, 1.09) | 0.05 | **1.02**<br>**(1.02, 1.03)** | <0.01 | **1.03**<br>**(1.02, 1.04)** | <0.01 | 1.03<br>(0.99, 1.07) | 0.14 | **1.04**<br>**(1.03, 1.05)** | <0.01 |
| Men | **1.02**<br>**(1.01, 1.02)** | <0.01 | 1.02<br>(0.98, 1.06) | 0.31 | **1.02**<br>**(1.01, 1.02)** | <0.01 | **1.02**<br>**(1.01, 1.02)** | <0.01 | **1.04**<br>**(1.01, 1.08)** | 0.02 | **1.04**<br>**(1.02, 1.06)** | <0.01 |
| Unknown/Missing | 0.95<br>(0.85, 1.06) | 0.38 | NE | - | 0.93<br>(0.81, 1.07) | 0.33 | 1.05<br>(0.88, 1.26) | 0.58 | 0.00<br>(0.00, Inf) | 1.00 | 0.70<br>(0.33, 1.47) | 0.35 |
| P for interaction effect | <0.01 |  | 0.66 |  | <0.01 |  | <0.01 |  | 0.39 |  | 0.66 |  |
| **Race <sup>c</sup>** |  |  |  |  |  |  |  |  |  |  |  |  |
|  |  |  | CSA |  | OSA |  | OUSA |  | Multiple sleep apneas |  | Procedure-based cases |  |
|  | PR (95% CI) | P-value | PR (95% CI) | P-value | PR (95% CI) | P-value | PR (95% CI) | P-value | PR (95% CI) | P-value | PR (95% CI) | P-value |
| AI/AN | **1.05**<br>**(1.04, 1.07)** | <0.01 | 1.14<br>(0.92, 1.41) | 0.23 | **1.07**<br>**(1.05, 1.09)** | <0.01 | 1.01<br>(0.98, 1.05) | 0.47 | 1.04<br>(0.84, 1.29) | 0.72 | 1.03<br>(0.95, 1.11) | 0.49 |
| Asian | 1.01<br>(1.00, 1.03) | 0.07 | 0.94<br>(0.68, 1.29) | 0.68 | 0.98<br>(0.97, 1.00) | 0.09 | 0.99<br>(0.97, 1.02) | 0.68 | 0.99<br>(0.78, 1.25) | 0.92 | **1.29**<br>**(1.23, 1.34)** | <0.01 |
| BAA | **1.01**<br>**(1.01, 1.02)** | <0.01 | 1.01<br>(0.93, 1.09) | 0.81 | **1.01**<br>**(1.01, 1.02)** | <0.01 | **1.02**<br>**(1.01, 1.03)** | <0.01 | 1.06<br>(0.97, 1.16) | 0.18 | 0.99<br>(0.97, 1.01) | 0.38 |
| NHPI | NE | - | NE | - | NE | - | NE | - | NE | - | NE | - |
| White | **1.02**<br>**(1.02, 1.03)** | <0.01 | 1.02<br>(0.99, 1.06) | 0.16 | **1.02**<br>**(1.02, 1.02)** | <0.01 | **1.03**<br>**(1.02, 1.03)** | <0.01 | **1.03**<br>**(1.00, 1.06)** | 0.04 | **1.04**<br>**(1.02, 1.05)** | <0.01 |
| Multiple | **1.04**<br>**(1.02, 1.06)** | <0.01 | 1.15<br>(0.89, 1.49) | 0.28 | **1.04**<br>**(1.01, 1.06)** | <0.01 | **1.05**<br>**(1.01, 1.09)** | <0.01 | 1.06<br>(0.87, 1.29) | 0.57 | 1.06<br>(0.98, 1.15) | 0.13 |
| Unknown/Missing | **1.02**<br>**(1.01, 1.02)** | <0.01 | 1.11<br>(1.00, 1.23) | 0.05 | **1.01**<br>**(1.00, 1.02)** | <0.01 | **1.02**<br>**(1.01, 1.03)** | <0.01 | 1.08<br>(0.97, 1.20) | 0.18 | **1.05**<br>**(1.02, 1.09)** | <0.01 |
| P for interaction | <0.01 |  | 0.60 |  | <0.01 |  | 0.20 |  | 0.99 |  | <0.01 |  |
| **Ethnicity<sup>d</sup>** |  |  |  |  |  |  |  |  |  |  |  |  |
| Hispanic | 1.00<br>(1.00, 1.01) | 0.14 | 1.07<br>(0.99, 1.17) | 0.09 | 1.00<br>(1.00, 1.01) | 0.14 | 1.00<br>(0.99, 1.01) | 1.00 | 1.04<br>(0.96, 1.13) | 0.29 | 1.01<br>(0.98, 1.03) | 0.58 |
| Non-Hispanic | **1.03**<br>**(1.02, 1.03)** | <0.01 | 1.02<br>(0.99, 1.05) | 0.27 | **1.02**<br>**(1.02, 1.03)** | <0.01 | **1.03**<br>**(1.02, 1.03)** | <0.01 | **1.03**<br>**(1.00, 1.06)** | 0.03 | **1.05**<br>**(1.04, 1.06)** | <0.01 |
| P for interaction | <0.01 |  | 0.19 |  | <0.01 |  | <0.01 |  | 0.90 |  | <0.01 |  |

Abbreviations: SDB=Sleep disordered breathing, CSA=Central Sleep Apnea, OSA=Obstructive Sleep Apnea, OUSA=Other/unspecified Sleep Apnea, AI/AN=American Indian/Alaskan Native, BAA=Black/African American, NHPI=Native Hawaiian/Pacific Islander, HVM=Health Vulnerability Module, PR=Prevalence ratios, CI=Confidence intervals.
Results from log-binomial models with reported outputs of exponentiated beta coefficients (prevalence ratios) and their 95% confidence intervals.
<sup>a</sup> Adjusted for sex, race, and ethnicity
<sup>b</sup> Adjusted for age category, race, and ethnicity
<sup>c</sup> Adjusted for age category, sex, and ethnicity
<sup>d</sup> Adjusted for age category, sex, and race
P for the interaction effect is the p-value of a likelihood ratio test comparing log-binomial models with and without an interaction term with the exposure.
Bolded values indicate statistical significance (P<0.05).
NE- Not estimated.

**eTable 8.** Sensitivity analysis for subgroup cross-sectional associations of per 0.1-unit increase in Environmental, Social, and Health Burden (ESHB) ranks with sleep-disordered breathing (SDB), central sleep apnea (CSA), obstructive sleep apnea (OSA), and other/unspecified sleep apnea (OUSA), multiple sleep apneas, and procedure-based cases

|  | Any SDB |  | Types of SDB |  |  |  |  |  |  |  |  |  |
| --- | --- | --- | --- | --- | --- | --- | --- | --- | --- | --- | --- | --- |
|  |  |  | CSA |  | OSA |  | OUSA |  | Multiple sleep apneas |  | Procedure-based cases |  |
|  | PR<br>(Bonferroni-corrected<br>95% CI) | P-value | PR<br>(Bonferroni-corrected<br>95% CI) | P-value | PR<br>(Bonferroni-corrected<br>95% CI) | P-value | PR<br>(Bonferroni-corrected<br>95% CI) | P-value | PR<br>(Bonferroni-corrected<br>95% CI) | P-value | PR<br>(Bonferroni-corrected<br>95% CI) | P-value |
| **Age category <sup>a</sup>** |  |  |  |  |  |  |  |  |  |  |  |  |
| 18-34 years | **1.02**<br>**(1.01, 1.03)** | <0.01 | 1.00<br>(0.88, 1.12) | 0.94 | **1.02**<br>**(1.01, 1.03)** | <0.01 | **1.02**<br>**(1.01, 1.04)** | <0.01 | 0.94<br>(0.80, 1.10) | 0.35 | **1.07**<br>**(1.03, 1.12)** | <0.01 |
| 35-49 years | **1.02**<br>**(1.01, 1.02)** | <0.01 | 1.02<br>(0.94, 1.11) | 0.57 | **1.02**<br>**(1.01, 1.02)** | <0.01 | **1.01**<br>**(1.00, 1.02)** | <0.01 | 1.02<br>(0.94, 1.11) | 0.54 | **1.03**<br>**(1.00, 1.07)** | 0.01 |
| ≥50 years | 1.00<br>(1.00, 1.01) | 0.17 | 1.01<br>(0.96, 1.05) | 0.79 | 1.00<br>(0.99, 1.00) | 0.28 | 1.01<br>(1.00, 1.01) | 0.06 | 0.99<br>(0.95, 1.03) | 0.64 | **1.04**<br>**(1.03, 1.06)** | <0.01 |
| P for interaction | <0.01 |  | 0.94 |  | <0.01 |  | 0.05 |  | 0.74 |  | 0.09 |  |
| **Sex <sup>b</sup>** |  |  |  |  |  |  |  |  |  |  |  |  |
| Women | **1.02**<br>**(1.02, 1.02)** | <0.01 | 1.04<br>(0.98, 1.10) | 0.16 | **1.02**<br>**(1.01, 1.02)** | <0.01 | **1.02**<br>**(1.02, 1.03)** | <0.01 | 1.00<br>(0.94, 1.05) | 0.89 | **1.05**<br>**(1.03, 1.07)** | <0.01 |
| Men | **1.00**<br>**(0.99, 1.00)** | <0.01 | 0.98<br>(0.93, 1.04) | 0.48 | **0.99**<br>**(0.99, 1.00)** | <0.01 | 1.00<br>(0.99, 1.01) | 0.51 | 0.99<br>(0.95, 1.04) | 0.75 | **1.04**<br>**(1.02, 1.07)** | <0.01 |
| Unknown/Missing | 0.99<br>(0.88, 1.12) | 0.85 | NE | - | 0.99<br>(0.85, 1.15) | 0.86 | 1.04<br>(0.83, 1.30) | 0.70 | 1.09<br>(0.42, 2.82) | 0.83 | 0.70<br>(0.33, 1.47) | 0.25 |
| P for interaction effect | <0.01 |  | 0.40 |  | <0.01 |  | <0.01 |  | 0.87 |  | 0.29 |  |
|  |  |  | CSA |  | OSA |  | OUSA |  | Multiple sleep apneas |  | Procedure-based cases |  |
|  | PR<br>(Bonferroni-corrected<br>95% CI) | P-value | PR<br>(Bonferroni-corrected<br>95% CI) | P-value | PR<br>(Bonferroni-corrected<br>95% CI) | P-value | PR<br>(Bonferroni-corrected<br>95% CI) | P-value | PR<br>(Bonferroni-corrected<br>95% CI) | P-value | PR<br>(Bonferroni-corrected<br>95% CI) | P-value |
| **Race <sup>c</sup>** |  |  |  |  |  |  |  |  |  |  |  |  |
| AI/AN | **1.02**<br>**(0.99, 1.05)** | 0.04 | 1.19<br>(0.79, 1.77) | 0.25 | **1.03**<br>**(1.00, 1.06)** | 0.02 | 0.98<br>(0.93, 1.04) | 0.43 | 1.07<br>(0.74, 1.54) | 0.63 | 1.05<br>(0.92, 1.20) | 0.28 |
| Asian | 0.99<br>(0.97, 1.01) | 0.10 | 0.87<br>(0.61, 1.23) | 0.27 | **0.97**<br>**(0.95, 0.99)** | <0.01 | 0.98<br>(0.95, 1.01) | 0.13 | 0.89<br>(0.67, 1.18) | 0.26 | **1.23**<br>**(1.15, 1.31)** | <0.01 |
| BAA | **1.01**<br>**(1.00, 1.01)** | 0.02 | 1.05<br>(0.91, 1.21) | 0.34 | 1.01<br>(1.00, 1.02) | 0.08 | **1.02**<br>**(1.00, 1.03)** | <0.01 | 1.04<br>(0.89, 1.21) | 0.49 | **0.95**<br>**(0.92, 0.99)** | <0.01 |
| NHPI | NE | - | NE | - | NE | - | NE | - | NE | - | NE | - |
| White | **1.01**<br>**(1.01, 1.01)** | <0.01 | 0.99<br>(0.94, 1.04) | 0.69 | **1.01**<br>**(1.00, 1.01)** | <0.01 | **1.01**<br>**(1.01, 1.02)** | <0.01 | 0.99<br>(0.95, 1.04) | 0.67 | **1.05**<br>**(1.03, 1.07)** | <0.01 |
| Multiple | 1.01<br>(0.98, 1.04) | 0.35 | 1.12<br>(0.74, 1.71) | 0.45 | 1.00<br>(0.97, 1.04) | 0.73 | 1.02<br>(0.97, 1.08) | 0.20 | 0.85<br>(0.63, 1.15) | 0.15 | 1.01<br>(0.90, 1.13) | 0.81 |
| Unknown/Missing | **1.01**<br>**(1.00, 1.02)** | <0.01 | 1.10<br>(0.94, 1.30) | 0.10 | 1.00<br>(0.99, 1.02) | 0.27 | 1.01<br>(0.99, 1.03) | 0.13 | 1.07<br>(0.90, 1.26) | 0.30 | **1.11**<br>**(1.05, 1.17)** | <0.01 |
| P for interaction | 0.01 |  | 0.25 |  | <0.01 |  | 0.28 |  | 0.48 |  | <0.01 |  |
| **Ethnicity <sup>d</sup>** |  |  |  |  |  |  |  |  |  |  |  |  |
| Hispanic | **0.98**<br>**(0.98, 0.99)** | <0.01 | 1.06<br>(0.95, 1.17) | 0.24 | **0.99**<br>**(0.98, 0.99)** | <0.01 | **0.98**<br>**(0.97, 0.99)** | <0.01 | 0.96<br>(0.87, 1.05) | 0.31 | 1.00<br>(0.97, 1.03) | 0.90 |
|  |  |  | CSA |  | OSA |  | OUSA |  | Multiple sleep apneas |  | Procedure-based cases |  |
|  | PR<br>(Bonferroni-corrected<br>95% CI) | P-value | PR<br>(Bonferroni-corrected<br>95% CI) | P-value | PR<br>(Bonferroni-corrected<br>95% CI) | P-value | PR<br>(Bonferroni-corrected<br>95% CI) | P-value | PR<br>(Bonferroni-corrected<br>95% CI) | P-value | PR<br>(Bonferroni-corrected<br>95% CI) | P-value |
| Non-Hispanic | **1.02**<br>**(1.01, 1.02)** | <0.01 | 1.00<br>(0.96, 1.04) | 0.89 | **1.01**<br>**(1.01, 1.02)** | <0.01 | **1.02**<br>**(1.02, 1.03)** | <0.01 | 1.00<br>(0.96, 1.04) | 0.94 | **1.06**<br>**(1.04, 1.07)** | <0.01 |
| P for interaction | <0.01 |  | 0.45 |  | <0.01 |  | <0.01 |  | 0.56 |  | <0.01 |  |

Abbreviations: SDB=Sleep disordered breathing, CSA=Central Sleep Apnea, OSA=Obstructive Sleep Apnea, OUSA=Other/unspecified Sleep Apnea, AI/AN=American Indian/Alaskan Native, BAA=
Black/African American, NHPI=Native Hawaiian/Pacific Islander, ESHB=Environmental, Social, and Health Burden, PR=Prevalence ratios, CI=Confidence intervals.
Results from log-binomial models with reported outputs of exponentiated beta coefficients (prevalence ratios) and their Bonferroni-corrected 95% confidence intervals.
<sup>a</sup> Adjusted for sex, race, and ethnicity (Bonferroni-corrected p-critical value = 0.017).
<sup>b</sup> Adjusted for age category, race, and ethnicity (Bonferroni-corrected p-critical value = 0.017).
<sup>c</sup> Adjusted for age category, sex, and ethnicity (Bonferroni-corrected p-critical value = 0.008).
<sup>d</sup> Adjusted for age category, sex, and race (Bonferroni-corrected p-critical value = 0.025).
P for the interaction effect are the p-value of a likelihood ratio test comparing log-binomial models with and without an interaction term.
Bolded values indicate statistical significance (P<0.05).
NE- Not estimated.

**eTable 9.** Sensitivity analysis for subgroup cross-sectional associations of per 0.1-unit increase in Environmental Burden Module (EBM) ranks with sleep-disordered breathing (SDB), central sleep apnea (CSA), obstructive sleep apnea (OSA), and other/unspecified sleep apnea (OUSA), multiple sleep apneas, and procedure-based cases

|  | Any SDB |  | Types of SDB |  |  |  |  |  |  |  |  |  |
| --- | --- | --- | --- | --- | --- | --- | --- | --- | --- | --- | --- | --- |
|  |  |  | CSA |  | OSA |  | OUSA |  | Multiple sleep apneas |  | Procedure-based cases |  |
|  | PR<br>(Bonferroni-corrected<br>95% CI) | P-value | PR<br>(Bonferroni-corrected<br>95% CI) | P-value | PR<br>(Bonferroni-corrected<br>95% CI) | P-value | PR<br>(Bonferroni-corrected<br>95% CI) | P-value | PR<br>(Bonferroni-corrected<br>95% CI) | P-value | PR<br>(Bonferroni-corrected<br>95% CI) | P-value |
| **Age category <sup>a</sup>** |  |  |  |  |  |  |  |  |  |  |  |  |
| 18-34 years | **0.98**<br>**(0.97, 0.99)** | <0.01 | 1.01<br>(0.90, 1.13) | 0.82 | **0.97**<br>**(0.96, 0.99)** | <0.01 | 0.99<br>(0.98, 1.00) | 0.06 | **0.80**<br>**(0.67, 0.96)** | <0.01 | **0.97**<br>**(0.93, 1.00)** | 0.03 |
| 35-49 years | **0.97**<br>**(0.97, 0.98)** | <0.01 | 0.98<br>(0.90, 1.07) | 0.57 | **0.97**<br>**(0.97, 0.98)** | <0.01 | **0.99**<br>**(0.98, 1.00)** | 0.01 | **0.90**<br>**(0.83, 0.98)** | <0.01 | **0.93**<br>**(0.90, 0.96)** | <0.01 |
| ≥50 years | **0.97**<br>**(0.97, 0.97)** | <0.01 | **0.95**<br>**(0.91, 1.00)** | 0.01 | **0.96**<br>**(0.96, 0.97)** | <0.01 | **0.99**<br>**(0.98, 1.00)** | <0.01 | **0.89**<br>**(0.85, 0.93)** | <0.01 | **0.94**<br>**(0.93, 0.96)** | <0.01 |
| P for interaction | <0.01 |  | 0.47 |  | <0.01 |  | 0.96 |  | 0.48 |  | 0.06 |  |
| **Sex <sup>b</sup>** |  |  |  |  |  |  |  |  |  |  |  |  |
| Women | **0.98**<br>**(0.98, 0.98)** | <0.01 | 0.97<br>(0.92, 1.03) | 0.19 | **0.98**<br>**(0.97, 0.98)** | <0.01 | 1.00<br>(0.99, 1.01) | 0.93 | **0.89**<br>**(0.84, 0.94)** | <0.01 | **0.94**<br>**(0.93, 0.96)** | <0.01 |
| Men | **0.96**<br>**(0.96, 0.97)** | <0.01 | 0.96<br>(0.91, 1.01) | 0.07 | **0.96**<br>**(0.95, 0.96)** | <0.01 | **0.98**<br>**(0.97, 0.99)** | <0.01 | **0.88**<br>**(0.84, 0.93)** | <0.01 | **0.94**<br>**(0.91, 0.96)** | <0.01 |
| Unknown/Missing | 0.98<br>(0.88, 1.10) | 0.73 | NE | - | 0.98<br>(0.85, 1.13) | 0.74 | 0.99<br>(0.80, 1.22) | 0.87 | 1.43<br>(0.52, 3.91) | 0.40 | 0.84<br>(0.44, 1.61) | 0.52 |
| P for interaction effect | <0.01 |  | 1.00 |  | <0.01 |  | <0.01 |  | 0.43 |  | 0.54 |  |
|  |  |  | CSA |  | OSA |  | OUSA |  | Multiple sleep apneas |  | Procedure-based cases |  |
|  | PR<br>(Bonferroni-corrected<br>95% CI) | P-value | PR<br>(Bonferroni-corrected<br>95% CI) | P-value | PR<br>(Bonferroni-corrected<br>95% CI) | P-value | PR<br>(Bonferroni-corrected<br>95% CI) | P-value | PR<br>(Bonferroni-corrected<br>95% CI) | P-value | PR<br>(Bonferroni-corrected<br>95% CI) | P-value |
| **Race <sup>c</sup>** |  |  |  |  |  |  |  |  |  |  |  |  |
| AI/AN | **0.94**<br>**(0.92, 0.97)** | <0.01 | 1.04<br>(0.73, 1.49) | 0.77 | **0.93**<br>**(0.90, 0.96)** | <0.01 | 0.97<br>(0.92, 1.02) | 0.12 | 0.97<br>(0.68, 1.39) | 0.82 | 0.97<br>(0.86, 1.10) | 0.55 |
| Asian | **0.97**<br>**(0.95, 0.98)** | <0.01 | 1.02<br>(0.72, 1.44) | 0.89 | **0.97**<br>**(0.95, 0.99)** | <0.01 | 0.99<br>(0.95, 1.02) | 0.26 | 0.88<br>(0.66, 1.18) | 0.25 | **0.90**<br>**(0.84, 0.96)** | <0.01 |
| BAA | **0.98**<br>**(0.98, 0.99)** | <0.01 | 1.05<br>(0.91, 1.20) | 0.38 | **0.98**<br>**(0.97, 0.99)** | <0.01 | 1.01<br>(0.99, 1.02) | 0.17 | 1.01<br>(0.87, 1.17) | 0.86 | **0.86**<br>**(0.82, 0.89)** | <0.01 |
| NHPI | NE | - | NE | - | NE | - | NE | - | NE | - | NE | - |
| White | **0.97**<br>**(0.97, 0.97)** | <0.01 | **0.95**<br>**(0.90, 0.99)** | <0.01 | **0.96**<br>**(0.96, 0.97)** | <0.01 | **0.99**<br>**(0.98, 0.99)** | <0.01 | **0.87**<br>**(0.83, 0.91)** | <0.01 | **0.95**<br>**(0.93, 0.97)** | <0.01 |
| Multiple | **0.95**<br>**(0.93, 0.98)** | <0.01 | 0.92<br>(0.61, 1.37) | 0.57 | **0.94**<br>**(0.91, 0.97)** | <0.01 | 0.99<br>(0.94, 1.04) | 0.68 | **0.72**<br>**(0.49, 1.04)** | 0.02 | 0.94<br>(0.84, 1.05) | 0.11 |
| Unknown/Missing | 1.00<br>(0.99, 1.01) | 0.64 | 1.07<br>(0.92, 1.25) | 0.23 | 0.99<br>(0.98, 1.00) | 0.07 | 1.00<br>(0.98, 1.02) | 0.64 | 1.01<br>(0.86, 1.18) | 0.89 | **1.07**<br>**(1.01, 1.12)** | <0.01 |
| P for interaction | <0.01 |  | 0.22 |  | <0.01 |  | <0.01 |  | 0.01 |  | <0.01 |  |
| **Ethnicity <sup>d</sup>** |  |  |  |  |  |  |  |  |  |  |  |  |
| Hispanic | **0.98**<br>**(0.97, 0.98)** | <0.01 | 1.01<br>(0.91, 1.11) | 0.88 | **0.98**<br>**(0.97, 0.98)** | <0.01 | **0.98**<br>**(0.97, 0.99)** | <0.01 | **0.85**<br>**(0.77, 0.93)** | <0.01 | 1.00<br>(0.97, 1.03) | 0.94 |
|  |  |  | CSA |  | OSA |  | OUSA |  | Multiple sleep apneas |  | Procedure-based cases |  |
|  | PR<br>(Bonferroni-corrected<br>95% CI) | P-value | PR<br>(Bonferroni-corrected<br>95% CI) | P-value | PR<br>(Bonferroni-corrected<br>95% CI) | P-value | PR<br>(Bonferroni-corrected<br>95% CI) | P-value | PR<br>(Bonferroni-corrected<br>95% CI) | P-value | PR<br>(Bonferroni-corrected<br>95% CI) | P-value |
| Non-Hispanic | **0.97**<br>**(0.97, 0.97)** | <0.01 | **0.96**<br>**(0.92, 1.00)** | 0.02 | **0.97**<br>**(0.96, 0.97)** | <0.01 | 1.00<br>(0.99, 1.00) | 0.14 | **0.89**<br>**(0.85, 0.92)** | <0.01 | **0.91**<br>**(0.90, 0.93)** | <0.01 |
| P for interaction | <0.01 |  | 0.61 |  | <0.01 |  | 0.01 |  | 0.11 |  | <0.01 |  |

Abbreviations: SDB=Sleep disordered breathing, CSA=Central Sleep Apnea, OSA=Obstructive Sleep Apnea, OUSA=Other/unspecified Sleep Apnea, AI/AN=American Indian/Alaskan Native, BAA=
Black/African American, NHPI=Native Hawaiian/Pacific Islander, EBM= Environmental Burden Module, PR=Prevalence ratios, CI=Confidence intervals.
Results from log-binomial models with reported outputs of exponentiated beta coefficients (prevalence ratios) and their Bonferroni-corrected 95% confidence intervals.
<sup>a</sup> Adjusted for sex, race, and ethnicity (Bonferroni-corrected p-critical value = 0.017).
<sup>b</sup> Adjusted for age category, race, and ethnicity (Bonferroni-corrected p-critical value = 0.017).
<sup>c</sup> Adjusted for age category, sex, and ethnicity (Bonferroni-corrected p-critical value = 0.008).
<sup>d</sup> Adjusted for age category, sex, and race (Bonferroni-corrected p-critical value = 0.025).
P for the interaction effect is the p-value of a likelihood ratio test comparing log-binomial models with and without an interaction term.
Bolded values indicate statistical significance (P<0.05).
NE- Not estimated.

**eTable 10.**
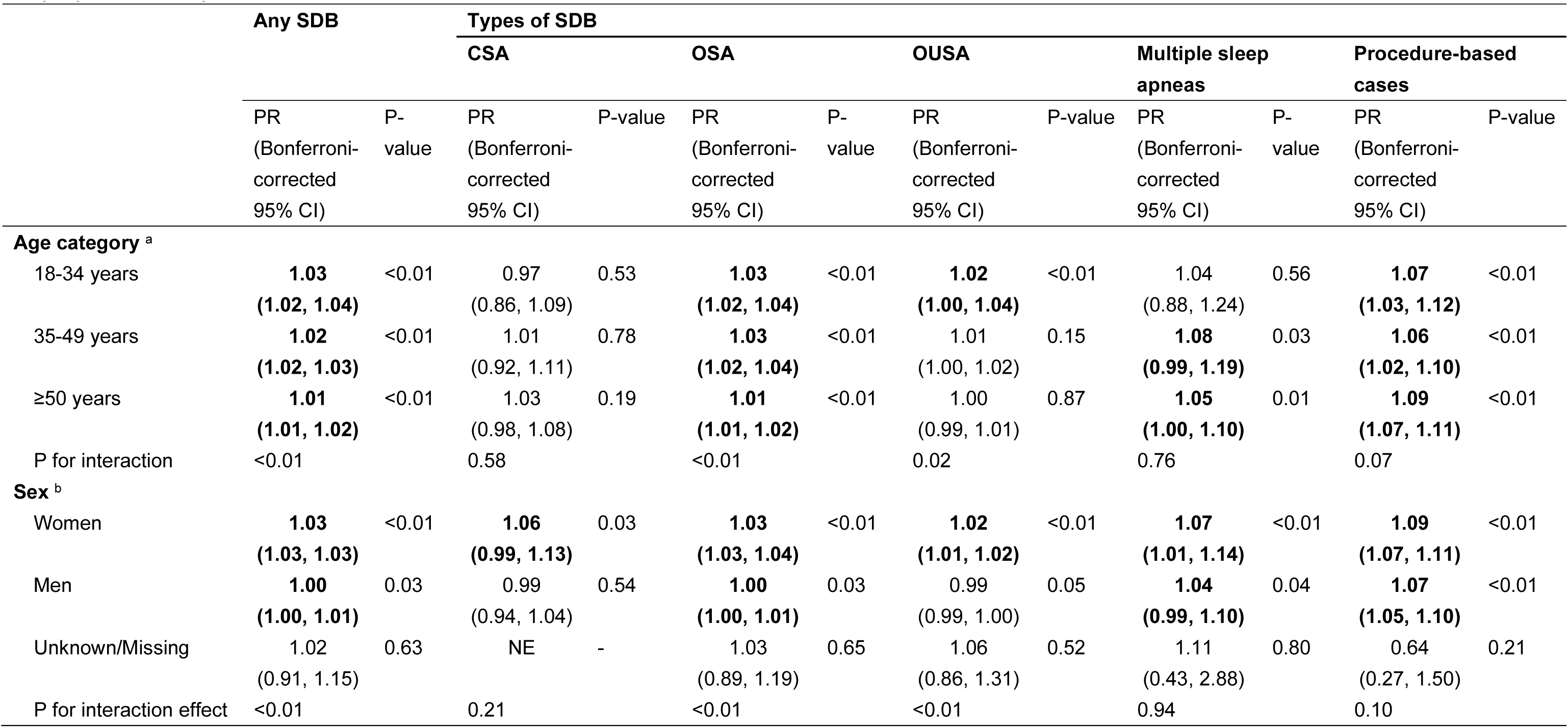
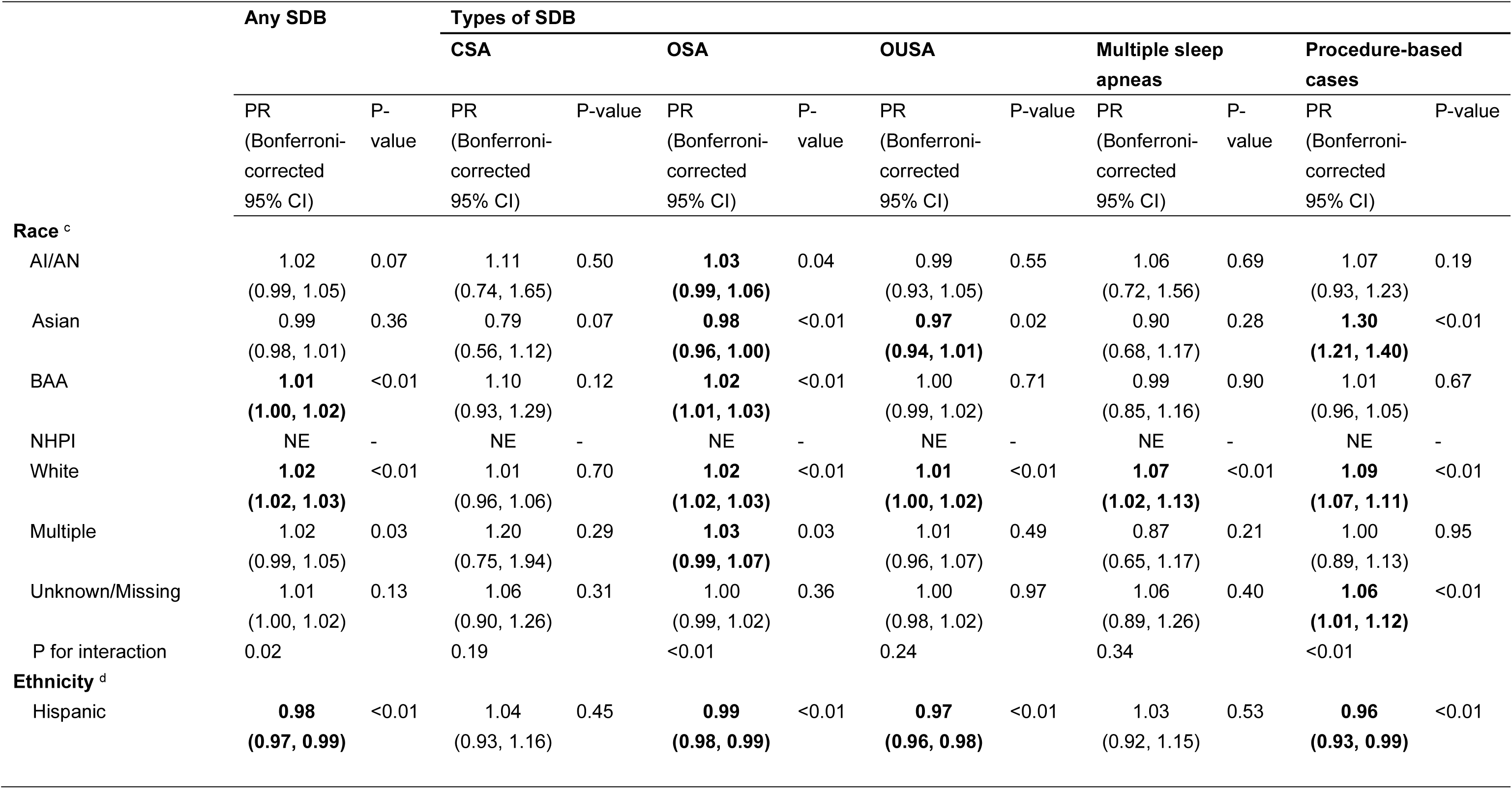
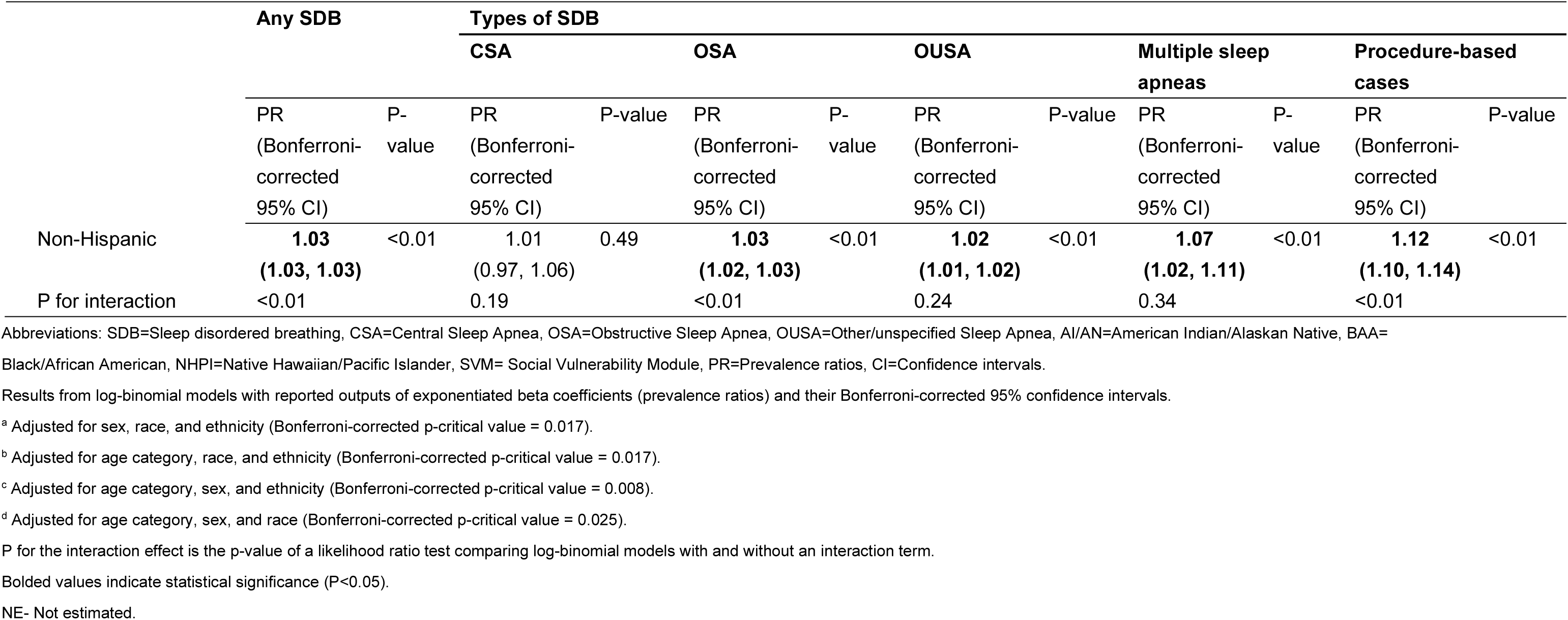
Sensitivity analysis for subgroup cross-sectional associations of per 0.1-unit increase in Social Vulnerability Module (SVM) ranks with sleep-disordered breathing (SDB), central sleep apnea (CSA), obstructive sleep apnea (OSA), and other/unspecified sleep apnea (OUSA), multiple sleep apneas, and procedure-based cases

**eTable 11.** Sensitivity analysis for subgroup cross-sectional associations of per 0.1-unit increase in Health Vulnerability Module (HVM) ranks with sleep-disordered breathing (SDB), central sleep apnea (CSA), obstructive sleep apnea (OSA), and other/unspecified sleep apnea (OUSA), multiple sleep apneas, and procedure-based cases

|  | Any SDB |  | Types of SDB |  |  |  |  |  |  |  |  |  |
| --- | --- | --- | --- | --- | --- | --- | --- | --- | --- | --- | --- | --- |
|  |  |  | CSA |  | OSA |  | OUSA |  | Multiple sleep apneas |  | Procedure-based cases |  |
|  | PR<br>(Bonferroni-corrected<br>95% CI) | P-value | PR<br>(Bonferroni-corrected<br>95% CI) | P-value | PR<br>(Bonferroni-corrected<br>95% CI) | P-value | PR<br>(Bonferroni-corrected<br>95% CI) | P-value | PR<br>(Bonferroni-corrected<br>95% CI) | P-value | PR<br>(Bonferroni-corrected<br>95% CI) | P-value |
| **Age category <sup>a</sup>** |  |  |  |  |  |  |  |  |  |  |  |  |
| 18-34 years | **1.04**<br>**(1.03, 1.05)** | <0.01 | 1.02<br>(0.92, 1.14) | 0.60 | **1.03**<br>**(1.02, 1.05)** | <0.01 | **1.04**<br>**(1.02, 1.05)** | <0.01 | 1.01<br>(0.87, 1.18) | 0.84 | **1.07**<br>**(1.03, 1.11)** | <0.01 |
| 35-49 years | **1.03**<br>**(1.03, 1.04)** | <0.01 | 1.03<br>(0.96, 1.12) | 0.31 | **1.03**<br>**(1.03, 1.04)** | <0.01 | **1.03**<br>**(1.02, 1.04)** | <0.01 | 1.05<br>(0.98, 1.13) | 0.1 | **1.05**<br>**(1.02, 1.08)** | <0.01 |
| ≥50 years | **1.02**<br>**(1.01, 1.02)** | <0.01 | 1.03<br>(0.99, 1.07) | 0.07 | **1.01**<br>**(1.01, 1.02)** | <0.01 | **1.02**<br>**(1.01, 1.02)** | <0.01 | **1.03**<br>**(1.00, 1.07)** | 0.03 | **1.03**<br>**(1.02, 1.05)** | <0.01 |
| P for interaction | <0.01 |  | 0.94 |  | <0.01 |  | <0.01 |  | 0.97 |  | 0.10 |  |
| **Sex <sup>b</sup>** |  |  |  |  |  |  |  |  |  |  |  |  |
| Women | **1.03**<br>**(1.02, 1.03)** | <0.01 | 1.04<br>(0.99, 1.10) | 0.05 | **1.02**<br>**(1.02, 1.03)** | <0.01 | **1.03**<br>**(1.02, 1.04)** | <0.01 | 1.03<br>(0.98, 1.08) | 0.14 | **1.04**<br>**(1.02, 1.06)** | <0.01 |
| Men | **1.02**<br>**(1.01, 1.02)** | <0.01 | 1.02<br>(0.97, 1.07) | 0.31 | **1.02**<br>**(1.01, 1.02)** | <0.01 | **1.02**<br>**(1.01, 1.02)** | <0.01 | **1.04**<br>**(1.00, 1.09)** | 0.02 | **1.04**<br>**(1.02, 1.06)** | <0.01 |
| Unknown/Missing | 0.95<br>(0.83, 1.09) | 0.38 | NE | - | 0.93<br>(0.78, 1.11) | 0.33 | 1.05<br>(0.84, 1.32) | 0.58 | 0.00<br>(0.00, Inf) | 1.00 | 0.70<br>(0.28, 1.73) | 0.35 |
| P for interaction effect | <0.01 |  | 0.66 |  | <0.01 |  | <0.01 |  | 0.39 |  | 0.66 |  |
|  |  |  | CSA |  | OSA |  | OUSA |  | Multiple sleep apneas |  | Procedure-based cases |  |
|  | PR<br>(Bonferroni-corrected<br>95% CI) | P-value | PR<br>(Bonferroni-corrected<br>95% CI) | P-value | PR<br>(Bonferroni-corrected<br>95% CI) | P-value | PR<br>(Bonferroni-corrected<br>95% CI) | P-value | PR<br>(Bonferroni-corrected<br>95% CI) | P-value | PR<br>(Bonferroni-corrected<br>95% CI) | P-value |
| **Race <sup>c</sup>** |  |  |  |  |  |  |  |  |  |  |  |  |
| AI/AN | **1.05**<br>**(1.03, 1.08)** | <0.01 | 1.14<br>(0.85, 1.53) | 0.23 | **1.07**<br>**(1.04, 1.10)** | <0.01 | 1.01<br>(0.97, 1.06) | 0.47 | 1.04<br>(0.77, 1.41) | 0.72 | 1.03<br>(0.92, 1.15) | 0.49 |
| Asian | 1.01<br>(0.99, 1.03) | 0.07 | 0.94<br>(0.60, 1.45) | 0.68 | 0.98<br>(0.96, 1.01) | 0.09 | 0.99<br>(0.96, 1.03) | 0.68 | 0.99<br>(0.71, 1.37) | 0.92 | **1.29**<br>**(1.21, 1.36)** | <0.01 |
| BAA | **1.01**<br>**(1.01, 1.02)** | <0.01 | 1.01<br>(0.90, 1.13) | 0.81 | **1.01**<br>**(1.00, 1.02)** | <0.01 | **1.02**<br>**(1.01, 1.03)** | <0.01 | 1.06<br>(0.94, 1.20) | 0.18 | 0.99<br>(0.96, 1.02) | 0.38 |
| NHPI | NE | - | NE | - | NE | - | NE | - | NE | - | NE | - |
| White | **1.02**<br>**(1.02, 1.03)** | <0.01 | 1.02<br>(0.98, 1.07) | 0.16 | **1.02**<br>**(1.02, 1.03)** | <0.01 | **1.03**<br>**(1.02, 1.03)** | <0.01 | **1.03**<br>**(0.99, 1.07)** | 0.04 | **1.04**<br>**(1.02, 1.05)** | <0.01 |
| Multiple | **1.04**<br>**(1.02, 1.07)** | <0.01 | 1.15<br>(0.81, 1.63) | 0.28 | **1.04**<br>**(1.00, 1.07)** | <0.01 | **1.05**<br>**(1.00, 1.10)** | <0.01 | 1.06<br>(0.81, 1.39) | 0.57 | 1.06<br>(0.95, 1.18) | 0.13 |
| Unknown/Missing | **1.02**<br>**(1.01, 1.03)** | <0.01 | 1.11<br>(0.96, 1.27) | 0.05 | **1.01**<br>**(1.00, 1.03)** | <0.01 | **1.02**<br>**(1.00, 1.04)** | <0.01 | 1.08<br>(0.93, 1.25) | 0.18 | **1.05**<br>**(1.01, 1.11)** | <0.01 |
| P for interaction | <0.01 |  | 0.60 |  | <0.01 |  | 0.20 |  | 0.99 |  | <0.01 |  |
| **Ethnicity <sup>d</sup>** |  |  |  |  |  |  |  |  |  |  |  |  |
| Hispanic | 1.00<br>(1.00, 1.01) | 0.14 | 1.07<br>(0.98, 1.18) | 0.09 | 1.00<br>(1.00, 1.01) | 0.14 | 1.00<br>(0.99, 1.01) | 1.00 | 1.04<br>(0.95, 1.15) | 0.29 | 1.01<br>(0.98, 1.04) | 0.58 |
|  |  |  | CSA |  | OSA |  | OUSA |  | Multiple sleep apneas |  | Procedure-based cases |  |
|  | PR<br>(Bonferroni-corrected<br>95% CI) | P-value | PR<br>(Bonferroni-corrected<br>95% CI) | P-value | PR<br>(Bonferroni-corrected<br>95% CI) | P-value | PR<br>(Bonferroni-corrected<br>95% CI) | P-value | PR<br>(Bonferroni-corrected<br>95% CI) | P-value | PR<br>(Bonferroni-corrected<br>95% CI) | P-value |
| Non-Hispanic | **1.03**<br>**(1.02, 1.03)** | <0.01 | 1.02<br>(0.98, 1.05) | 0.27 | **1.02**<br>**(1.02, 1.03)** | <0.01 | **1.03**<br>**(1.02, 1.03)** | <0.01 | **1.03**<br>**(1.00, 1.07)** | 0.03 | **1.05**<br>**(1.04, 1.07)** | <0.01 |
| P for interaction | <0.01 |  | 0.60 |  | <0.01 |  | 0.20 |  | 0.99 |  | <0.01 |  |

Abbreviations: SDB=Sleep disordered breathing, CSA=Central Sleep Apnea, OSA=Obstructive Sleep Apnea, OUSA=Other/unspecified Sleep Apnea, AI/AN=American Indian/Alaskan Native, BAA=
Black/African American, NHPI=Native Hawaiian/Pacific Islander, HVM=Health Vulnerability Module, PR=Prevalence ratios, CI=Confidence intervals.
Results from log-binomial models with reported outputs of exponentiated beta coefficients (prevalence ratios) and their Bonferroni-corrected 95% confidence intervals.
<sup>a</sup> Adjusted for sex, race, and ethnicity (Bonferroni-corrected p-critical value = 0.017).
<sup>b</sup> Adjusted for age category, race, and ethnicity (Bonferroni-corrected p-critical value = 0.017).
<sup>c</sup> Adjusted for age category, sex, and ethnicity (Bonferroni-corrected p-critical value = 0.008).
<sup>d</sup> Adjusted for age category, sex, and race (Bonferroni-corrected p-critical value = 0.025).
P for the interaction effect is the p-value of a likelihood ratio test comparing log-binomial models with and without an interaction term.
Bolded values indicate statistical significance (P<0.05).
NE- Not estimated.

**eTable 12.**
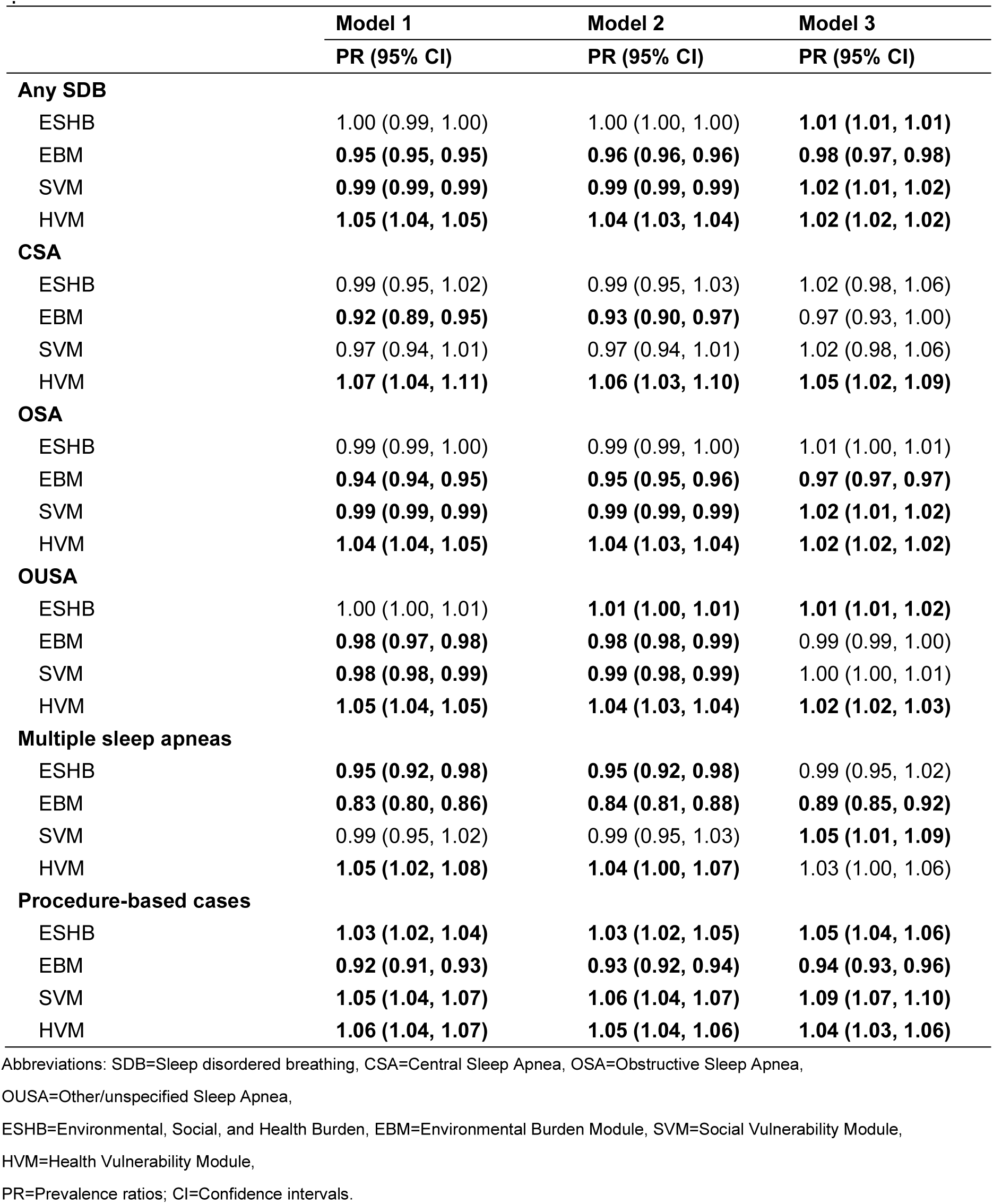
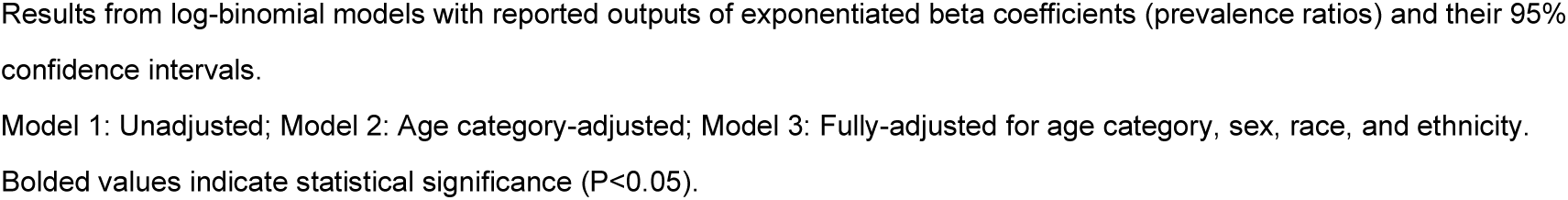
Sensitivity analysis for cross-sectional association of per 0.1-unit increase Environmental, Social, and Health Burden (ESHB), Environmental Burden Module (EBM), Social Vulnerability Module (SVM), and Health Vulnerability Module (HVM) ranks with sleep-disordered breathing (SDB), central sleep apnea (CSA), obstructive sleep apnea (OSA), and other/unspecified sleep apnea (OUSA), multiple sleep apneas, and procedure-based cases

|  | **Model 1** | **Model 2** | **Model 3** |
| --- | --- | --- | --- |
|  | **PR (95% CI)** | **PR (95% CI)** | **PR (95% CI)** |
| **Any SDB** |  |  |  |
| ESHB | 1.00 (0.99, 1.00) | 1.00 (1.00, 1.00) | **1.01 (1.01, 1.01)** |
| EBM | **0.95 (0.95, 0.95)** | **0.96 (0.96, 0.96)** | **0.98 (0.97, 0.98)** |
| SVM | **0.99 (0.99, 0.99)** | **0.99 (0.99, 0.99)** | **1.02 (1.01, 1.02)** |
| HVM | **1.05 (1.04, 1.05)** | **1.04 (1.03, 1.04)** | **1.02 (1.02, 1.02)** |
| **CSA** |  |  |  |
| ESHB | 0.99 (0.95, 1.02) | 0.99 (0.95, 1.03) | 1.02 (0.98, 1.06) |
| EBM | **0.92 (0.89, 0.95)** | **0.93 (0.90, 0.97)** | 0.97 (0.93, 1.00) |
| SVM | 0.97 (0.94, 1.01) | 0.97 (0.94, 1.01) | 1.02 (0.98, 1.06) |
| HVM | **1.07 (1.04, 1.11)** | **1.06 (1.03, 1.10)** | **1.05 (1.02, 1.09)** |
| **OSA** |  |  |  |
| ESHB | 0.99 (0.99, 1.00) | 0.99 (0.99, 1.00) | 1.01 (1.00, 1.01) |
| EBM | **0.94 (0.94, 0.95)** | **0.95 (0.95, 0.96)** | **0.97 (0.97, 0.97)** |
| SVM | **0.99 (0.99, 0.99)** | **0.99 (0.99, 0.99)** | **1.02 (1.01, 1.02)** |
| HVM | **1.04 (1.04, 1.05)** | **1.04 (1.03, 1.04)** | **1.02 (1.02, 1.02)** |
| **OUSA** |  |  |  |
| ESHB | 1.00 (1.00, 1.01) | **1.01 (1.00, 1.01)** | **1.01 (1.01, 1.02)** |
| EBM | **0.98 (0.97, 0.98)** | **0.98 (0.98, 0.99)** | 0.99 (0.99, 1.00) |
| SVM | **0.98 (0.98, 0.99)** | **0.99 (0.98, 0.99)** | 1.00 (1.00, 1.01) |
| HVM | **1.05 (1.04, 1.05)** | **1.04 (1.03, 1.04)** | **1.02 (1.02, 1.03)** |
| **Multiple sleep apneas** |  |  |  |
| ESHB | **0.95 (0.92, 0.98)** | **0.95 (0.92, 0.98)** | 0.99 (0.95, 1.02) |
| EBM | **0.83 (0.80, 0.86)** | **0.84 (0.81, 0.88)** | **0.89 (0.85, 0.92)** |
| SVM | 0.99 (0.95, 1.02) | 0.99 (0.95, 1.03) | **1.05 (1.01, 1.09)** |
| HVM | **1.05 (1.02, 1.08)** | **1.04 (1.00, 1.07)** | 1.03 (1.00, 1.06) |
| **Procedure-based cases** |  |  |  |
| ESHB | **1.03 (1.02, 1.04)** | **1.03 (1.02, 1.05)** | **1.05 (1.04, 1.06)** |
| EBM | **0.92 (0.91, 0.93)** | **0.93 (0.92, 0.94)** | **0.94 (0.93, 0.96)** |
| SVM | **1.05 (1.04, 1.07)** | **1.06 (1.04, 1.07)** | **1.09 (1.07, 1.10)** |
| HVM | **1.06 (1.04, 1.07)** | **1.05 (1.04, 1.06)** | **1.04 (1.03, 1.06)** |

Abbreviations: SDB=Sleep disordered breathing, CSA=Central Sleep Apnea, OSA=Obstructive Sleep Apnea,
OUSA=Other/unspecified Sleep Apnea,
ESHB=Environmental, Social, and Health Burden, EBM=Environmental Burden Module, SVM=Social Vulnerability Module,
HVM=Health Vulnerability Module,
PR=Prevalence ratios; CI=Confidence intervals.
Results from log-binomial models with reported outputs of exponentiated beta coefficients (prevalence ratios) and their 95% confidence intervals.
Model 1: Unadjusted; Model 2: Age category-adjusted; Model 3: Fully-adjusted for age category, sex, race, and ethnicity.
Bolded values indicate statistical significance ( $P < 0.05$ ).

